# Modelling the within-host dynamics of *Plasmodium vivax* hypnozoite activation: an analysis of the SPf66 vaccine trial

**DOI:** 10.1101/2024.01.03.24300707

**Authors:** Somya Mehra, Francois Nosten, Christine Luxemburger, Nicholas J White, James A Watson

## Abstract

*Plasmodium vivax* parasites can lie dormant in the liver as hypnozoites, activating weeks to months after sporozoite inoculation to cause relapsing malarial illness. It is not known what biological processes govern hypnozoite activation. We use longitudinal data from the most detailed cohort study ever conducted in an area where both *Plasmodium falciparum* and *P. vivax* were endemic to fit a simple within-host mathematical model of *P. vivax* hypnozoite activation. 1344 children living on the Thailand-Myanmar border were followed each day for 21 months. There were 2504 vivax and 1164 falciparum malaria symptomatic episodes recorded over 1988 person-years. The model assumes that hypnozoites activate independently at a constant rate (‘exponential clock model’). When this model was embedded in a stochastic framework for repeated infectious mosquito bites, with seasonality inferred from the incidence of clinical falciparum malaria episodes, it explained the observed temporal patterns of multiple (up to 13) recurrent vivax malaria episodes. Under this model we estimate the mean dormancy period for a single hypnozoite to be 6 months (i.e. a half-life of 4 months). We use the calibrated within-host model to characterise population level overdispersion in the risk of relapse, and assess the potential utility of a serological test for radical cure in low transmission settings. We show that mefloquine treatment of falciparum malaria eliminates early vivax relapses; and that there are substantially more *P. vivax* recurrences than expected under the model following artesunate monotherapy treatment for falciparum malaria. These results suggest that hypnozoites can be activated by symptomatic malarial illness.

## Introduction

*Plasmodium vivax* and *P. ovale* malaria parasites are characterised by their ability to lie dormant in liver cells for variable periods, with periodic activation leading to relapsing bloodstream infections. This has allowed *P. vivax* to extend into temperate regions and persist in people during periods which are unfavourable for the mosquito vectors [1]. Prevention of relapse requires treatment with 8-aminoquinoline antimalarials (“radical cure”), which cause haemolytic toxicity in patients with glucose-6-phosphate dehydrogenase (G6PD) deficiency [2], making elimination of this important parasite difficult [3].

The biology of relapse is not well understood. How hypnozoites activate in the liver remains largely unknown [4, 5]. A major challenge is the difficulty in developing appropriate *in vitro* platforms to study the fundamental biology of *P. vivax* [6–9]. A key question is how much variation in hypnozoite activation is driven by intrinsic versus extrinsic stimuli. There are at least two sub-species of *P. vivax* ; the prevalent short latency or “tropical” type which relapses at short intervals — although the frequency and intervals vary geographically — and a “temperate” long latency type which has an interval of approximately 8–9 months between either inoculation and the primary illness, or between the primary illness and the first relapse [10–12].

An intrinsic biological clock mechanism is likely to explain long latency in temperate strains of *P. vivax*. Several external triggers of activation have been proposed for both short and long latency strains. These range from seasonal changes [13] and the bites of potential malaria vectors [14] to the febrile inflammatory response [3, 15] and haemolysis [16]. One hypothesis is that each febrile malaria episode itself (regardless of the species) can trigger subsequent hypnozoite activation [3]. This hypothesis was based on the observation that the risk of vivax malaria after acute falciparum malaria in both adults and children enrolled in therapeutic studies in South East Asia was nearly as high as the risk after acute vivax malaria, and the intervals between acute illness and relapse were similar [17–20]: i.e. in therapeutic studies a falciparum malaria episode was highly predictive of a vivax relapse. Mosquito co-infections are relatively rare in low transmission settings [21]. This suggested that falciparum malaria could trigger hypnozoite activation [3]. It has been theorised that an external sensing mechanism may be coupled with spontaneous activation at a low baseline rate [3, 4].

Mathematical models of vivax relapse in combination with human data cannot prove or disprove a given biological theory. Further, mathematical and biological parsimony do not have the same heuristic values: biological parsimony can be considered from the viewpoint of ease of selection or evolution, but need not to have a simple mathematical formulation. However, if a mathematically simple model could predict patterns of relapse robustly within and across individuals, this would provide some evidence in favour of the corresponding biological theory, with the caveat that biological theories and mathematical models do not have a one-to-one equivalence. In addition, a mathematically simple model of relapse which could explain the majority of the inter- and intra-individual variability in relapse risk in endemic areas would have practical utility. For example, it would help understand where to target drug administration campaigns for radical cure. The mathematically simplest “base” model of vivax relapse was formulated by Michael White and colleagues: a constant activation rate per hypnozoite, with independent and identical dynamics for each hypnozoite and no external triggers [22]. This results in an exponentially distributed time to relapse, with a rate conditional on the hypnozoite burden. We refer to this as the “exponential clock” model.

In this work we derive analytical likelihoods for the exponential clock model allowing us to infer the model parameters from longitudinal data on recurrent infections. We fit the model to data from a very detailed prospective trial of a *P. falciparum* vaccine which proved ineffective [23]. In this trial, 1344 children aged 2 to 15 years of age living in a region of low seasonal malaria transmission situated on the North-Western border of Thailand were seen each day for 21 months. This is the most detailed cohort study of vivax malaria ever conducted. We show that in this epidemiological setting the exponential clock model can capture patterns of multiple recurrent infections over time across childhood age groups, with up to 13 symptomatic vivax episodes recorded per child. We found higher than expected rates of vivax after *P. falciparum* monoinfections treated with artesunate monotherapy, which is compatible with an external triggering mechanism. We also found that unlike chloroquine [24], mefloquine treatment provides sufficient drug exposure for one month to eliminate emerging relapses, a previously unappreciated public health benefit. This post-treatment prophylactic effect may have masked any triggering effects as most falciparum episodes were treated with artesunate-mefloquine. We use the fitted model to quantify overdispersion in relapse risk and characterise the potential utility of a serological test to guide radical cure.

## Results

### Modelling hypnozoite activation as an exponential clock

The exponential clock model is the simplest mathematical model describing hypnozoite activation. Under this model each hypnozoite activates independently with an exponentially distributed time to activation with constant rate *η*. To capture the observed variation in recurrence patterns, we add stochasticity in (i) the timing of infectious mosquito bites (non-homogeneous Poisson process); (ii) the inoculated sporozoite batch sizes (assumed geometric distribution); and (iii) sporozoite “destiny” (hypnozoite versus immediately developing form: binomial distribution, giving rise to correlation in the burden of primary infection and relapse associated with each bite). Finally, we model the acquisition of anti-disease immunity using age as a proxy for previous malaria exposure. Anti-disease immunity is defined as the probability that a blood-stream malaria infection results in symptomatic illness, and is modelled as a flexible decreasing deterministic function of age. An overview of the model parameters is given in Table 1.

**Table 1:** Summary of within-host model parameters and their interpretation.

| Parameter | Interpretation |
| --- | --- |
| $\eta$ | Rate of activation of each hypnozoite (the inverse is the expected duration of hypnozoite carriage) |
| $\nu$ | Mean size of successful sporozoite inoculum (i.e. number of sporozoites that undergo immediate development or form a viable hypnozoite that is destined to activate) |
| $p_{\text{rel}}$ | Probability that a sporozoite becomes a hypnozoite that will subsequently activate ($1 - p_{\text{rel}}$ undergo immediate development) |
| $\lambda(t)$ | Force of inoculation (i.e. anopheline vector infective bite rate per human) at time $t$ |
| $\rho$ | Proportional reduction in the probability of symptomatic bloodstream infection for each hypnozoite activation and immediate sporozoite development event between the ages of 2 and 15 years |
| $\gamma$ | Shape coefficient for the age-dependent anti-disease masking probability |

We show that the model likelihood is analytically tractable when formulated as an open network of infinite server queues (see Appendices B.1 and B.2, and [25]). Although very simple, this within-host model gives rise to complex temporal patterns of infection. There are three key structural features.

First, the exponential clock mechanism results in a linear relationship between the relapse rate in an individual and the size of their hypnozoite reservoir. Both the expected time to next relapse, and the variation in the time to next relapse, scale inversely with the hypnozoite burden. This results in short inter-relapse intervals initially following an infectious bite, with subsequent lengthening of the inter-relapse intervals for subsequent relapses as the hypnozoite reservoir is depleted. This pattern is consistent with observations from artificial infection studies [26, 27]. Under this model relapses occurring in quick succession can be explained by either a high activation rate or a large hypnozoite burden.

Second, the number of hypnozoites carried by individuals in low transmission areas (loosely defined as one *P. vivax* infectious bite per year, which approximates most vivax endemic areas outside of Oceania) is highly variable (overdispersed). This results from variation in the number and timing of infectious bites (infrequent inoculation), and variation in the number of sporozoites inoculated per bite. We expect zero inflation in the distribution of hypnozoite burdens in young children, persisting for a time scale determined by the force of inoculation (in particular, the rate at which infants are exposed to infectious mosquito bites). As the child grows under a constant force of inoculation, repeated mosquito sporozoite inoculation, and thus hypnozoite accumulation, is eventually balanced out by hypnozoite activation, resulting in a steady state distribution for the hypnozoite burden. The expected hypnozoite burden reaches approximately 90% of the mean steady state within four half-lives of hypnozoite carriage.

Third, there is a non-linear relationship between age and the number of symptomatic relapses. This non-linear relationship arises as a consequence of the competing time scales of hypnozoite accrual and acquisition of anti-disease immunity. Hypnozoite accrual is determined by the expected duration of hypnozoite carriage. The acquisition of anti-disease immunity is determined by the force of inoculation, and the access to treatment, but is modelled indirectly as a deterministic function of age. We note that age-dependent differences in mosquito inoculation rates will also result in age-dependent differences in hypnozoite burdens [28, 29].

### The SPf66 vaccine trial

The SPf66 vaccine trial (1993–1995) was the largest and most detailed cohort study in vivax malaria ever conducted. This was a randomised controlled double-blind trial assessing a synthetic protein vaccine candidate for falciparum malaria [23]. The vaccine was shown to have no clinical efficacy. 1344 children between 2 and 15 years of age, living in a camp for displaced persons situated on the Thailand-Myanmar border, were followed daily for symptoms over 21 months. Most of the children were of the Karen ethnic group. Malaria was diagnosed by microscopy and promptly treated under supervision. Typically, vivax monoinfections were treated with chloroquine monotherapy (25 mg base/kg over 3 days), while uncomplicated falciparum monoinfections and mixed infections were treated with artesunate (4 mg/kg for 3 days) and mefloquine (25 mg/kg on day 2 of treatment) combination therapy. Artesunate monotherapy for five days was given to some children with hyperparasitemic falciparum infections (see Appendix A.1 for details). Severe malaria cases were treated with artemether (intramuscular administration of 3.2 mg/kg on day 1, then 1.6 mg/kg per day), followed by oral artesunate-mefloquine combination therapy. Inter-consultation intervals show that chloroquine and mefloquine conferred little post-treatment prophylactic protection against falciparum malaria (Figure A.1), consistent with contemporaneous reports of widespread multidrug resistant *P. falciparum* [30]. Radical cure with primaquine was not standard of care at the time, and only a few children with very frequently relapsing vivax infections were given primaquine. Access to antimalarial drugs was tightly regulated [31], and self-treatment was very unlikely.

During the study the population of the camp remained relatively stable [31]. Malaria transmission occurred within the confines of the camp which was located within hill forest [31]. There were twice-yearly peaks in incidence during the rainy season (May to June) and cool season (November to December) respectively (Figure 1A) [32]. Around April 1994 artesunate-mefloquine combination therapy became the first line treatment for falciparum malaria in the camp, resulting in a substantial decline in falciparum transmission [31] (Figure 1A).

**Figure 1:**
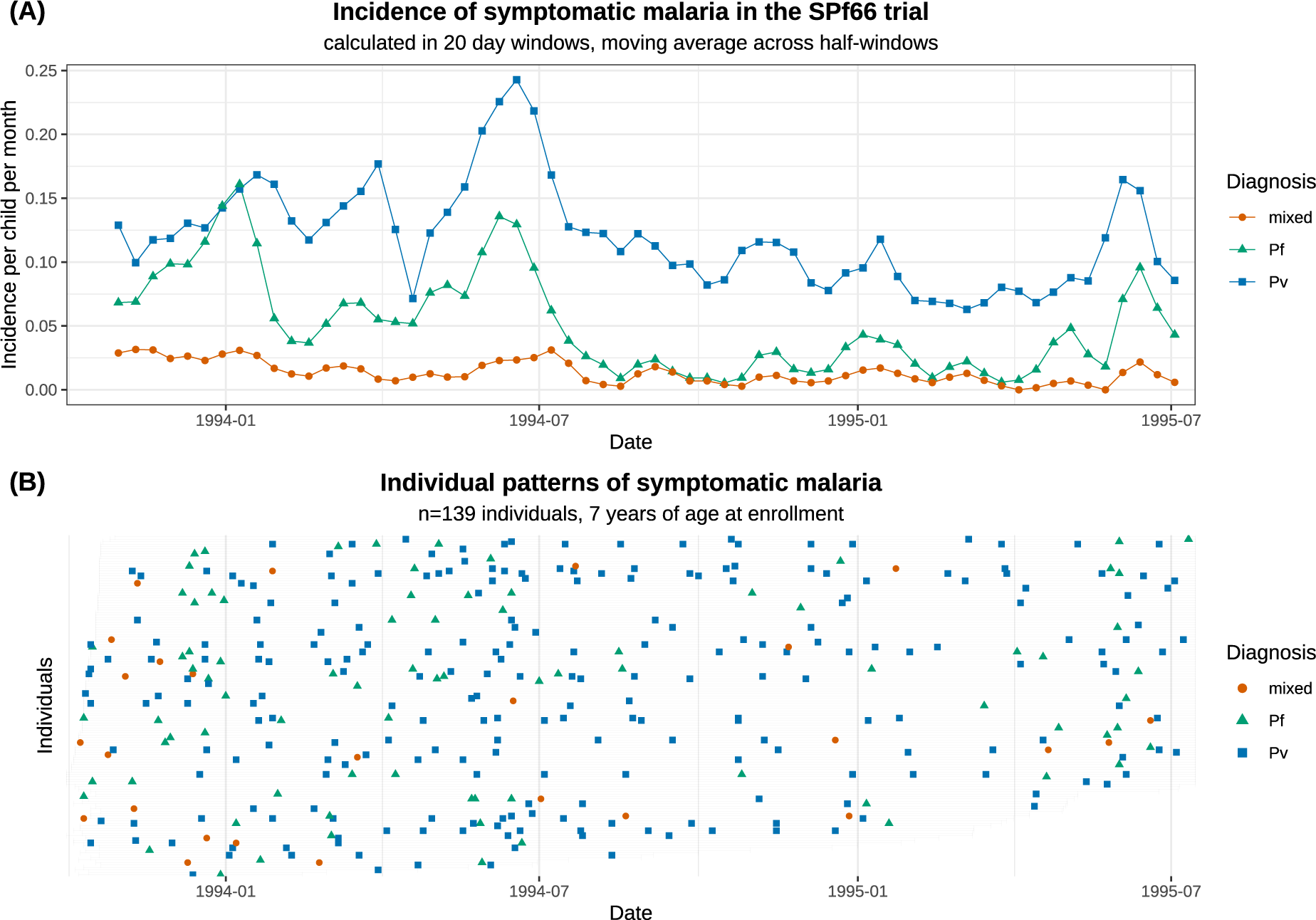
Symptomatic falciparum and vivax malaria in the SPf66 trial. Panel A shows the aggregate monthly incidence of symptomatic malaria (blue: *P. vivax* ; green: *P. falciparum*; orange: mixed infections). Mixed infections are double-counted as vivax and falciparum episodes. Panel B shows individual data from all participants aged 7 years at enrolment (grey bar shows the period of active detection; documented absences from the camp are not shown).

This study provides detailed data on the timing and frequency of symptomatic *P. vivax* and *P. falciparum* infections in a low-transmission co-endemic setting. After screening for treatment failures (*n* = 13 *P. vivax* monoinfections, *n* = 13 *P. falciparum* monoinfections and *n* = 1 mixed infection; see Appendix A.1), a total of 2504 symptomatic *P. vivax* infections and 1164 symptomatic *P. falciparum* infections were observed over 1988 person-years follow-up, of which 293 (8.7%) were mixed infections (Table 2). Up to 13 symptomatic vivax and 7 symptomatic falciparum episodes were recorded for each child (Figure 1B). Age structure in the incidence of symptomatic malaria is summarised in Appendix A.4.

**Table 2:** Summary of episodes detected through active clinical follow-up (daily surveillance) in the SPf66 trial (after removing likely treatment failures). Mixed infections are double-counted as both vivax and falciparum episodes. Age-stratified incidence rates are computed as the ratio of the total number of symptomatic episodes recorded within an age group, and the cumulative time at risk adjusted for left/right-censoring, documented absences from the camp and post-treatment prophylaxis.

| Age at enrolment (years) | 2 | 3 | 4 | 5 | 6 | 7 | 8 | 9 | 10 | 11 | 12 | 13 | 14 | 15 | Overall |
| --- | --- | --- | --- | --- | --- | --- | --- | --- | --- | --- | --- | --- | --- | --- | --- |
| Number of children | 73 | 87 | 76 | 121 | 114 | 139 | 125 | 116 | 147 | 96 | 101 | 82 | 49 | 18 | 1344 |
| Mean follow-up per child (years) | 1.50 | 1.57 | 1.53 | 1.51 | 1.50 | 1.53 | 1.47 | 1.53 | 1.45 | 1.42 | 1.46 | 1.43 | 1.30 | 1.01 | 1.48 |
| Number of Pv episodes | 141 | 171 | 187 | 263 | 255 | 272 | 275 | 188 | 265 | 142 | 172 | 123 | 39 | 11 | 2504 |
| Children with $\geq 1$ Pv episode | 44<br>(60%) | 57<br>(66%) | 50<br>(66%) | 88<br>(73%) | 74<br>(65%) | 80<br>(58%) | 81<br>(65%) | 65<br>(56%) | 82<br>(56%) | 51<br>(53%) | 51<br>(50%) | 43<br>(52%) | 21<br>(43%) | 5<br>(28%) | 792<br>(59%) |
| Mean Pv episodes per child | 1.93 | 1.97 | 2.46 | 2.17 | 2.24 | 1.96 | 2.20 | 1.62 | 1.80 | 1.48 | 1.70 | 1.50 | 0.80 | 0.61 | 1.86 |
| Pv incidence rate (per year) | 1.42 | 1.37 | 1.80 | 1.60 | 1.66 | 1.40 | 1.67 | 1.16 | 1.37 | 1.14 | 1.28 | 1.14 | 0.66 | 0.63 | 1.39 |
| Number of Pf episodes | 58 | 76 | 72 | 122 | 102 | 108 | 117 | 88 | 125 | 91 | 95 | 65 | 38 | 7 | 1164 |
| Children with $\geq 1$ Pf episode | 35<br>(48%) | 46<br>(53%) | 43<br>(57%) | 71<br>(59%) | 59<br>(52%) | 68<br>(49%) | 68<br>(54%) | 55<br>(47%) | 74<br>(50%) | 54<br>(56%) | 53<br>(52%) | 43<br>(52%) | 22<br>(45%) | 5<br>(28%) | 696<br>(52%) |
| Mean Pf episodes per child | 0.79 | 0.87 | 0.95 | 1.01 | 0.89 | 0.78 | 0.94 | 0.76 | 0.85 | 0.95 | 0.94 | 0.79 | 0.78 | 0.39 | 0.87 |
| Pf incidence rate (per year) | 0.54 | 0.57 | 0.63 | 0.69 | 0.61 | 0.52 | 0.65 | 0.51 | 0.60 | 0.68 | 0.66 | 0.57 | 0.61 | 0.39 | 0.60 |
| Number of mixed episodes | 23 | 26 | 25 | 38 | 29 | 26 | 34 | 19 | 28 | 15 | 14 | 12 | 3 | 1 | 293 |
| Children with $\geq 1$ mixed episode | 16<br>(22%) | 20<br>(23%) | 19<br>(25%) | 28<br>(23%) | 22<br>(19%) | 23<br>(17%) | 31<br>(25%) | 17<br>(15%) | 22<br>(15%) | 12<br>(13%) | 14<br>(14%) | 12<br>(15%) | 3<br>(6%) | 1<br>(6%) | 240<br>(18%) |
| Mean mixed episodes per child | 0.32 | 0.30 | 0.33 | 0.31 | 0.25 | 0.19 | 0.27 | 0.16 | 0.19 | 0.16 | 0.14 | 0.15 | 0.06 | 0.06 | 0.22 |
| Mixed incidence rate (per year) | 0.23 | 0.21 | 0.24 | 0.23 | 0.19 | 0.13 | 0.21 | 0.12 | 0.15 | 0.12 | 0.10 | 0.11 | 0.05 | 0.06 | 0.16 |

### Calibration to longitudinal recurrence data from the SPf66 trial

We fitted our within-host model to all symptomatic vivax episodes observed in the SPf66 trial. Falciparum infections were used to estimate seasonality. The model accounted for post-treatment prophylaxis, including an extended period of protection following mefloquine treatment during which bloodstream infection is eliminated; and a “bunching” effect following chloroquine treatment [33], whereby bloodstream infection is variably delayed resulting in aggregation of times to patent relapse (Appendix A.1). The assumptions of the model are summarised in Box 1, with detailed model fitting results provided in Appendix C.1.

### Duration of hypnozoite carriage

Patterns of recurrent symptomatic vivax malaria in the SPf66 trial were strongly informative for the hypnozoite activation rate *η*. Under the model we estimated a hypnozoite activation rate of 1*/*171 day*^−^*^1^ (95% credible interval [CrI] 1*/*206 to 1*/*144 day*^−^*^1^), amounting to a half-life of 118 days (95% CrI 100 to 143 days) for a population of hypnozoites (Table 2A). Detailed simulation studies showed that these estimates were robust to mis-specification of the sporozoite batch size (i.e. under- or over-dispersion of the size of the sporozoite inocula relative to the geometric distribution, Appendix D.1), in addition to mis-specification of the ratio of immediately-developing forms to hypnozoites (Appendix D.2). The estimated duration of hypnozoite carriage for the SPf66 trial (171 days) is similar to previous estimates for South East Asian strains (approximately 190 days), obtained by calibrating the exponential clock model with geometrically-distributed hypnozoite loads to data for the time to first relapse [22].

### Sporozoite inocula

The mean number of *successful* sporozoites inoculated per infective bite was estimated to be 6.9 (95% CrI 5.4 to 8.7) when assuming a ratio of immediately activating forms to hypnozoites of 6:4 (Table 2A). This estimate is a lower bound for the true mean sporozoite batch size, since hypnozoite death is unidentifiable based on longitudinal recurrence data (Appendix B.1). The corresponding estimate for the average number of hypnozoites per bite was 2.7 (95% CrI 2.2 to 3.5). However, this estimate was largely insensitive to the assumed ratio of immediately-developing forms to hypnozoites (Appendix D.2).

### Relative burden of relapse and primary infection

The ratio of relapsing to primary *P. vivax* infections within the cohort was estimated as 3.4 (95% CrI 2.8 to 4.2) (Table 2A), implying that *∼*75% of observed symptomatic *P. vivax* infections were relapses. This is lower than previous estimates of the relative relapse burden (*∼*95%) in patients seeking treatment for vivax malaria in the same transmission setting after treatment for their initial enrolment episode [35, 36]. Importantly, these previous estimates do not count the enrolment infection (which is less likely to be a relapse in low transmission setting). Both the ratio of relapsing to primary infection and the force of primary bloodstream infection (Figure 2B) — estimated to be in the vicinity of 0.4 (a)symptomatic primary infections per person per year, with declining transmission in the second half of the study — were relatively robust to mis-specification of the sporozoite batch size distribution (Appendix D.1).

#### Box 1

Exponential clock model assumptions

There are four structural assumptions in the model:

1. Each hypnozoite has an independent exponentially-distributed time to activation, with no external triggers.
2. The number of inoculated sporozoites is geometrically distributed.*^†^*
3. The ratio of immediately-developing sporozoites to hypnozoites is 6:4 (informed by recent *in vivo* and *in vitro* experiments on the Chesson strain of *P. vivax* [34]).*^†^*
4. The probability of clinical symptoms for each bloodstream infection decays monotonically as a function of age (i.e. age is considered a proxy for prior exposure and thus immunity), with all cases assumed to be symptomatic in 2 year olds.

There are four assumptions in the observational model (context dependent):

1. Each treatment is associated with a fixed period of subsequent prophylactic protection, during which all emerging bloodstream infections are eliminated. For *P. vivax* this is 15 days following chloroquine, 35 days following mefloquine, and 15 days following artesunate or quinine monotherapy.
2. Chloroquine provides an additional fixed period delaying (but not eliminating) *P. vivax* multiplication in the blood.
3. There is no age-stratification or heterogeneity in the force of inoculation for *P. vivax* between birth and 15 years of age.*^†^*
4. Seasonal fluctuations in the incidence of symptomatic falciparum malaria are a proxy for seasonal fluctuations in the force of inoculation for *P. vivax*.

† The impact of model mis-specification have been assessed for these assumptions

Based on inter-recurrence intervals, we can classify individual episodes probabilistically as relapses or reinfections (Appendix E.4).

### The relationship between age and symptomatic vivax malaria

If the average duration of dormancy for each hypnozoite is 6 months (half-life of 4 months), then the hypnozoite reservoir would reach steady state after approximately 2 years of life under a constant force of inoculation [25]. In the absence of age-stratification in the force of inoculation, this would imply that the number of hypnozoites was distributed identically across all age groups in the SPf66 trial: while there would have been variation in the hypnozoite burden across individuals, systematic differences across age groups would not be expected. Slowly eliminated antimalarial treatments affect relapse intervals, but age related differences in pharmacokinetics are considered negligible. As such, the model fit would imply that all age structure in the incidence of symptomatic vivax was determined by anti-disease immunity alone, leading to a monotonic decline in the incidence of symptomatic vivax across age groups (Figure 2C).

**Figure 2:**
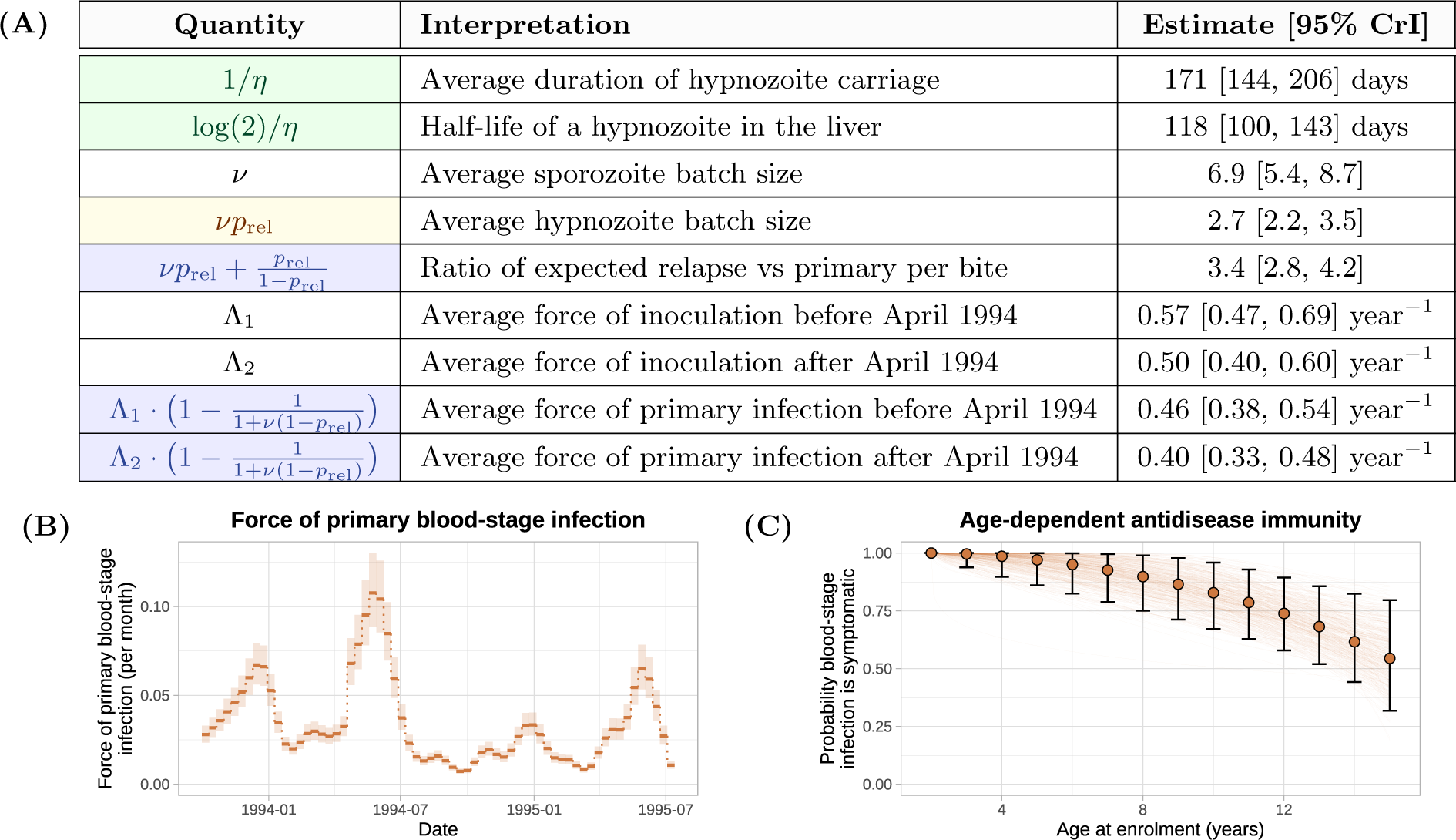
Summary of posterior model estimates. Panel A: posterior median estimates [95% credible intervals (CrI)] for quantities of epidemiological interest. Blue: estimates robust to mis-specification in the sporozite batch distribution; yellow: estimates robust to misspecification of the hypnozoite fating probability; green: estimates robust to both forms of mis-specification. Panel B: force of inoculation inferred from the incidence of symptomatic falciparum malaria over 10 day windows (shaded regions show 95% CrI). Scaling factors for the ratio of vivax to falciparum inoculation rates are estimated separately before and after April 1994 (day 200 of the study) when there was a camp-wide shift to artesunate-mefloquine treatment of falciparum malaria. Panel C: age-dependent probability of symptomatic bloodstream infection, relative to a 2 year old. Error bars indicate 95% CrI for each age group; continuous curves are plotted for 200 parameter combinations (*ρ, γ*) sampled from the posterior.

However, the data showed a non-monotonic relationship between age and symptomatic vivax malaria (Figure 3A). Peak incidence occurred around the age of four (*p* = 1 *×* 10*^−^*^4^ using an asymptotic bootstrap for the Mack-Wolfe test for umbrella alternatives with unknown peak, Appendix A.4). There were clear discrepancies between the model fit and the observed data for the age dependent cumulative *P. vivax* incidence rates (Figure 3B). This discrepancy may be explained by age-dependent differences in the frequency of mosquito inoculation in the first years of life. It is unlikely that there were substantial differences in the force of inoculation for children *>*2 years as there is no evidence for age structure in the incidence of symptomatic falciparum malaria (*p* = 0.8 for the Kruskall-Wallis test, Appendix A.4), a direct correlate of mosquito inoculation. However, it is plausible that infants under 2 years had lower exposure to infectious mosquito bites [37–39], leading to fewer hypnozoites accrued by age 2. As such, we re-fitted the model with the force of inoculation for infants under 2 years scaled down by a series of fixed factors. Estimates for the hypnozoite activation rate, the mean sporozoite batch size, the force of inoculation, and age-dependent anti disease masking characteristic were unchanged; however, the description of the non-monotonic age structure in the incidence of symptomatic vivax malaria was improved (Appendix C.2.3).

**Figure 3:**
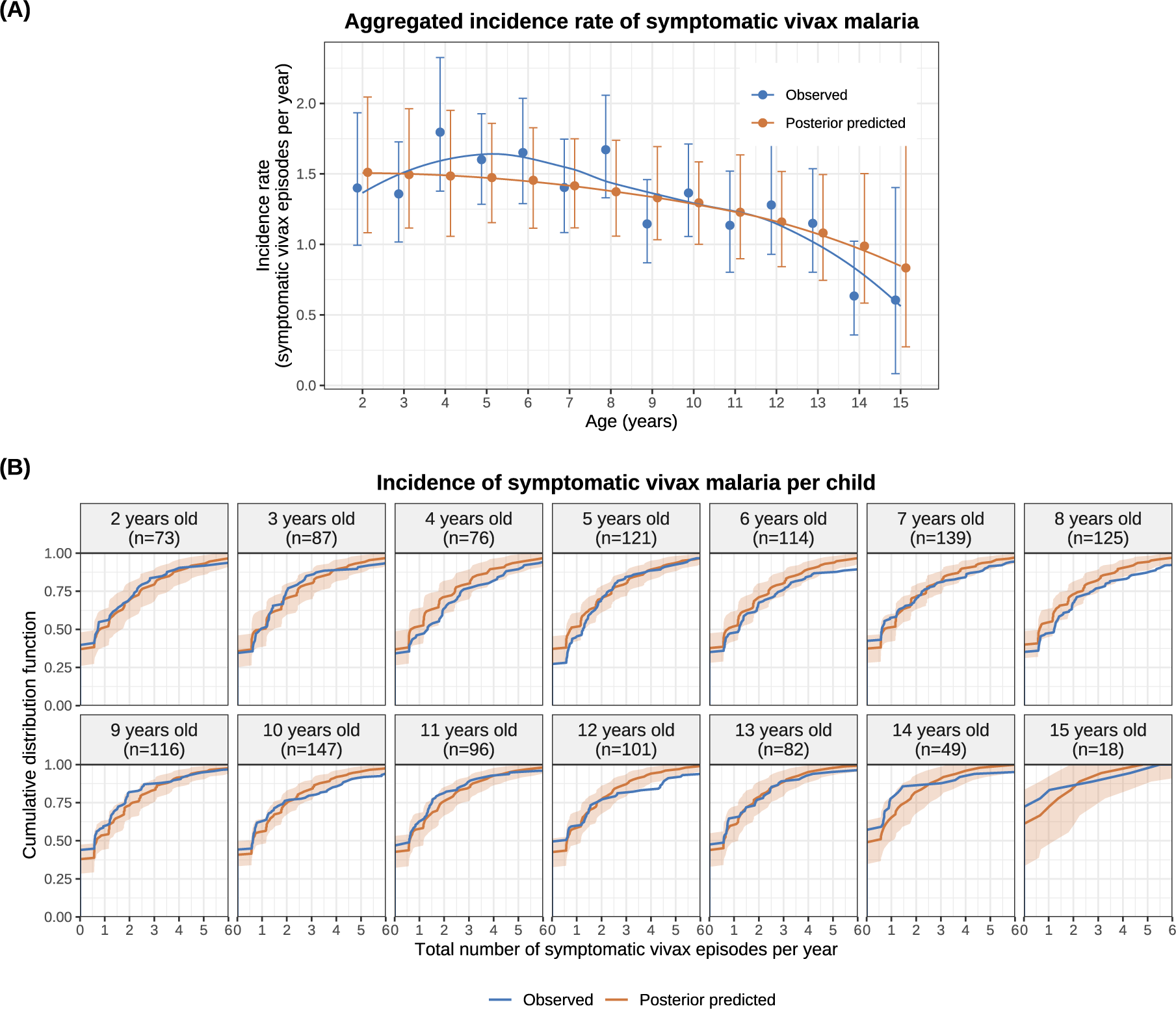
Characterising the model fit by age group. Panel A shows the observed versus posterior predicted incidence rate of symptomatic vivax malaria, stratified by age at enrolment; error bars show 95% confidence intervals for observed data (generated via bootstrapping), and 95% credible intervals for posterior predicted data. Panel B shows the cumulative distributions of the incidence of symptomatic vivax malaria per child. Shaded areas: 95% credible intervals. Simulation of data under the posterior predictive distribution is described in Appendix C.2.1.

### Vivax malaria following falciparum malaria

Several studies have described higher than expected rates of vivax relapse following symptomatic falciparum [17–20]. We assessed whether the exponential clock model without external triggering could explain the proportion of symptomatic *P. vivax* malaria episodes observed after symptomatic *P. falciparum* monoinfections in the SPf66 trial. We compared model predicted versus observed vivax recurrence rates following *P. falciparum* monoinfections, accounting for the documented history of vivax recurrence. Model predicted rates of vivax recurrence are dependent on correctly modelling seasonality which was a limitation in our model fit^1^. However, observed rates of vivax recurrence were largely compatible with model predictions at fixed time points (i.e. irrespective of *P. falciparum* infection) throughout the SPf66 trial (see Appendix, Figure C.4).

Falciparum monoinfections were stratified by schizonticidal treatment (artesunate monotherapy, *n* = 120 versus artesunate-mefloquine combination therapy, *n* = 607). The rates of vivax malaria 20 to 80 days after *P. falciparum* monoinfections treated with artesunate monotherapy were substantially higher than expected under the model (red, Figure 4). In contrast, the rates of vivax malaria in the same periods following artesunate-mefloquine were largely within the expected range (blue, Figure 4). These patterns persisted after matching treatment groups for the number of previous vivax episodes and the time of the baseline falciparum monoinfection^2^.

**Figure 4:**
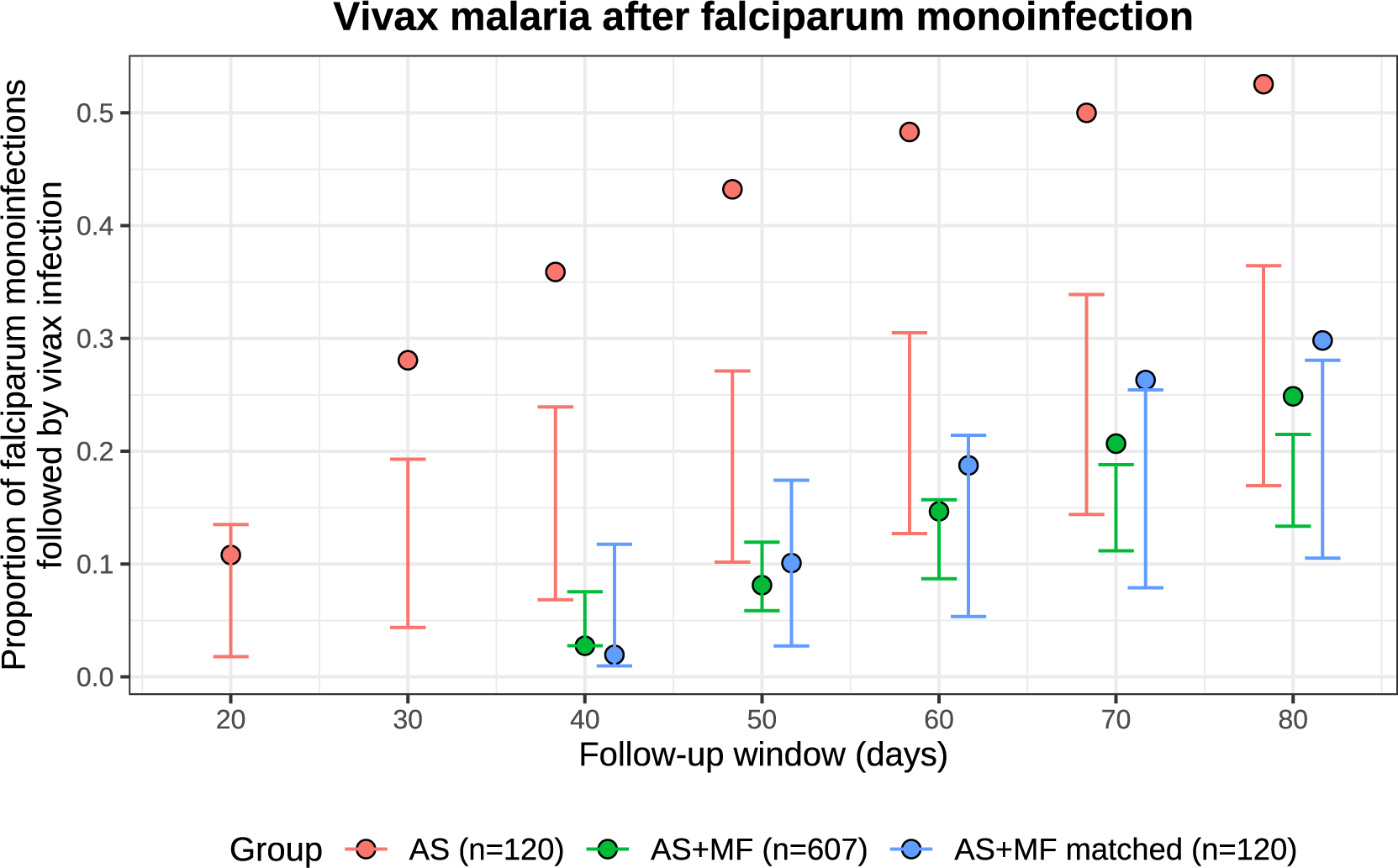
Vivax after falciparum monoinfection in the SPf66 trial. The model predicted conditional probabilities for each follow-up window are shown by the vertical bars (99% CrI). The observed proportions are shown by the filled circles. Red: artesunate (AS) monotherapy; blue: artesunate-mefloquine combination therapy (AS+MF); green: AS+MF episodes matched against AS episodes for the time of the baseline episode (to control for seasonality) and the history of vivax recurrence.

### Overdispersion and zero inflation of the hypnozoite burden

The model estimated considerable overdispersion in the hypnozoite burden across individuals, resulting in population heterogeneity in the risk of relapse. The steady state hypnozoite distribution is negative binomial [25] (see Appendix E.2). Under the assumption that our estimates for the mean (geometrically-distributed) sporozoite batch size (*ν* = 6.9) and the mean duration of hypnozoite carriage (1*/η* = 171 days) are generalisable across endemic settings with shortlatency *P. vivax*, we characterised hypnozoite burdens in individuals at steady state across a range of transmission settings.

Figure 5A shows the model predicted distribution in the number of hypnozoites carried by individuals as a function of different transmission intensities (for simplicity this assumes a constant force of inoculation). This substantial overdispersion has important consequences for *P. vivax* elimination and control. For example, suppose it was possible to interrupt human-to-mosquito transmission temporarily (for instance, by mass antimalarial drug treatment, or the widespread administration of ivermectin or an effective insecticide). The expected time until a threshold proportion of individuals no longer carry hypnozoites is determined by the overdispersion in hypnozoite carriage (Figure 5B). If the force of inoculation was 0.5 infectious bites per year before interruption of transmission, then only 27% of individuals are predicted to carry hypnozoites, but it would still take 1.6 years on average to eliminate hypnozoites with probability 98% in all individuals (Figure 5B).

**Figure 5:**
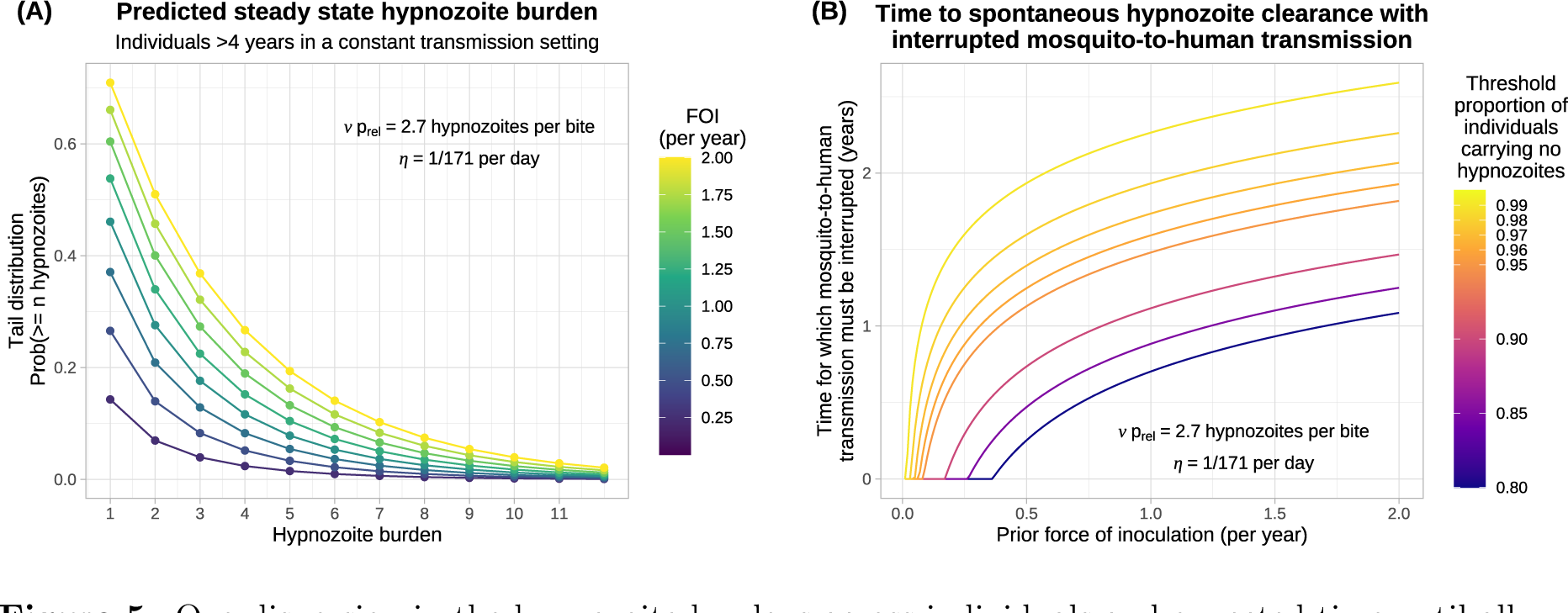
Overdispersion in the hypnozoite burdens across individuals and expected time until all hypnozoites spontaneously clear, as a function of the force of inoculation (FOI). Panel A shows the tail distribution for the size of the hypnozoite reservoir. Panel B shows the duration of time for which mosquito-to-human mosquito transmission must be interrupted for a threshold proportion of individuals to spontaneously clear all hypnozoites.

### Assessing the utility of a serological test for radical cure

There has been recent interest in using a multiplex serological assay to identify individuals who have had a recent bloodstream *P. vivax* infection [41, 42]. The serological test is based on a panel of 8 *P. vivax* specific antibodies and is estimated to have *>* 80% specificity and *>* 80% sensitivity in detecting bloodstream infections which occurred within the previous 9 months. This test was developed using data from low transmission settings (Thailand, Brazil and the Solomon Islands [41]), and included both symptomatic and asymptomatic infections, but was shown to exhibit lower accuracy under more intense transmission in Peru [43]. As recent bloodstream infection is predictive of current hypnozoite carriage, serological testing could allow for targeted administration of 8-aminoquinoline drugs. A cluster randomised trial is underway which will compare serological mass screening and provision of radical cure treatment (MSAT) of all eligible individuals against treatment of symptomatic cases only (https://www.pvstatem.eu/).

Using our calibrated model, we assessed the utility of such a test under different epidemiological scenarios. The positive predictive value of a recent bloodstream infection becomes comparable to the baseline probability of hypnozoite carriage for a randomly sampled individual as the force of inoculation increases (dashed grey vs green line, Figure 6). For transmission intensities lower than 0.5 infectious bites per year, individuals who have had recent bloodstream infections are more than twice as likely to harbour hypnozoites than randomly-sampled individuals; for a force of inoculation of 2 bites per year they are only 1.1 times more likely to harbour hypnozoites. The false omission rate (conditional probability of hypnozoite carriage given no recurrence occurred within the preceding 9 months) is generally low but rises to 18% under an inoculation rate of 2 infectious bites per year (maroon line, Figure 6). This demonstrates the limited utility of serological testing to identify hypnozoite carriage outside of near elimination settings [43].

**Figure 6:**
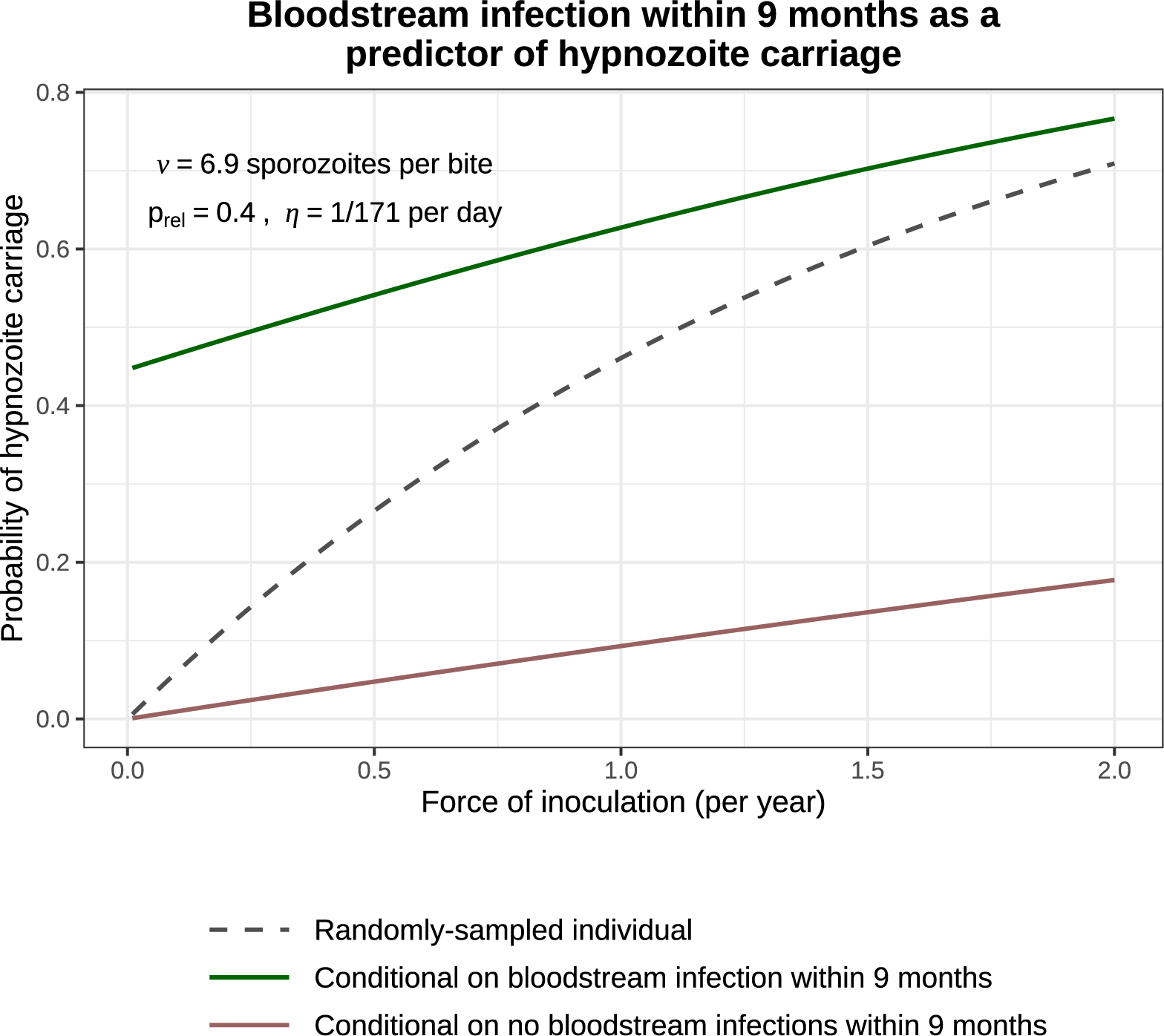
The probability of hypnozoite carriage. This compares the probability that a randomly-sampled individual carries hypnozoites (dashed grey line) with the conditional probability given a bloodstream infection within the previous 9 months (i.e. the positive predictive value, green) and the conditional probability given no bloodstream infection within the previous 9 months (i.e. the false omission rate, maroon).

Based on these observations, we used our model to assess the theoretical impact of serological MSAT, relative to MDA (both providing radical cure). To accommodate population heterogeneity in transmission, we allow the force of inoculation to be Gamma-distributed [44]. Given a serological test with 80% specificity and 80% sensitivity for recent recurrence, we predict that approximately 75% of hypnozoite carriers (in the eligible population) are correctly-targeted for treatment through MSAT, with this figure largely insensitive to both transmission intensity and heterogeneity (Figure 7A). The reduction in overtreatment (i.e. anti-hypnozoite treatment of individuals who do not carry hypnozoites) for MSAT versus MDA is most pronounced in low and heterogeneous transmission settings (Figure 7B). For example, in a population where individuals are bitten once a year on average, but 20% of individuals receive 80% of all infective bites (Gini coefficient 0.985), we predict serological MSAT to yield a 2.8 fold reduction in overtreatment relative to MDA (in the eligible population). This does not incorporate the small additional benefit from 8-aminoquinoline prophylaxis (i.e. preventing acquisition). It is important to stress that assessment of a complex intervention such as MSAT cannot be done only through the lens of a mathematical model. Our within-host model provides one piece in a complex cost-effectiveness framework. To characterise relative cost-effectiveness of MSAT versus MDA in near-elimination settings (i.e. where mass treatment could plausibly be cost-effective), it is necessary to take into account the operational costs and complexity (for example, serological screening without a point-of-care test requires two successive visits per individual) as well as the opportunity costs.

**Figure 7:**
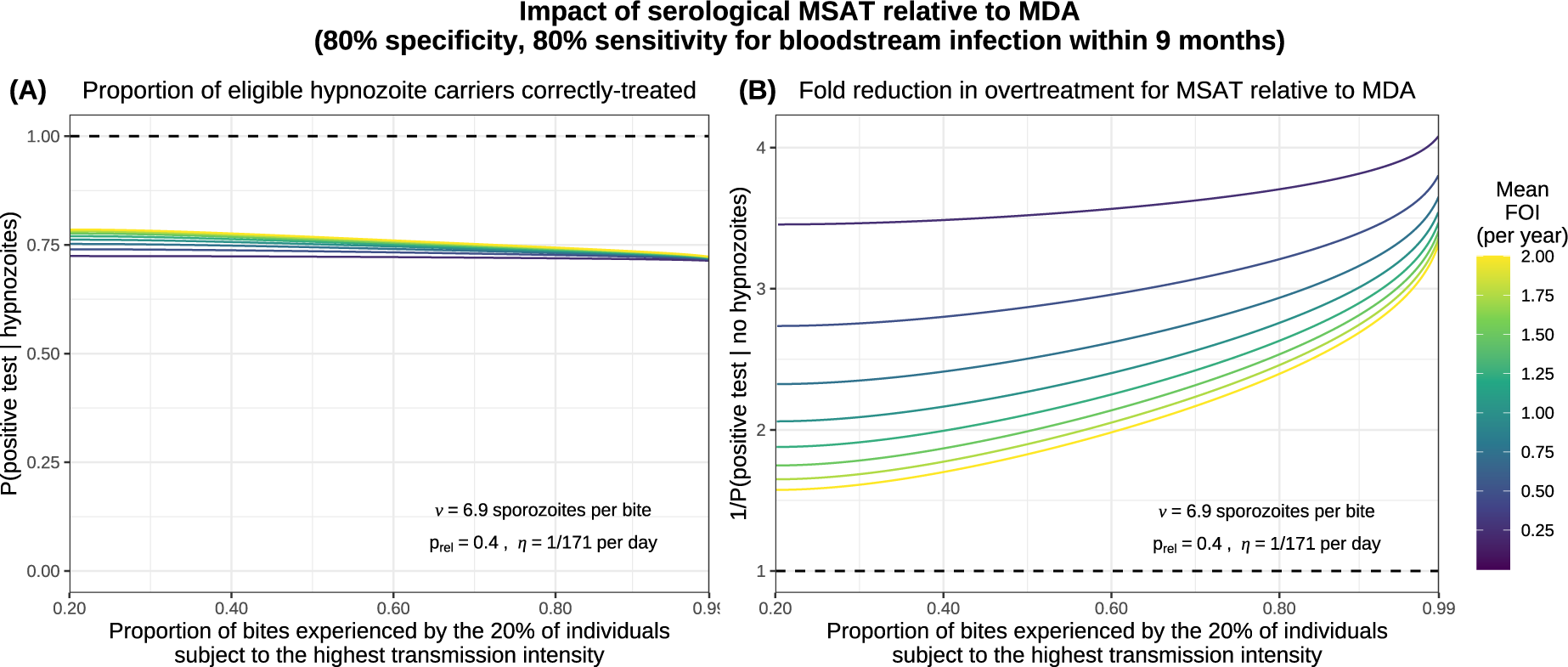
The modelled impact of serological MSAT (assuming 80% sensitivity and 80% specificity for bloodstream infection within 9 months) relative to MDA, as a function of transmission intensity and population heterogeneity. We model each individual to be subject to a constant force of inoculation (FOI), sampled from a Gamma distribution [44] parameterised by the mean biting rate in the population and the proportion of bites experienced by the 20% of individuals subject to the highest transmission intensity. Panel A shows the proportion of eligible hypnozoite carriers who are correctly treated in the MSAT setting, while panel B shows the fold reduction in overtreatment for MSAT versus MDA.

## Discussion

*Plasmodium vivax* is now the predominant malaria parasite species in most of endemic Asia and Latin America. It is an important cause of morbidity and developmental delay in children, and in high transmission settings it contributes to childhood mortality via severe anaemia [45]. The main barrier to *P. vivax* elimination is relapse. Unless an effective radical cure regimen with an 8-aminoquinoline antimalarial is given, treatment of the bloodstream infection does not prevent subsequent hypnozoite activation and consequent recurrent malaria. Characterising the within-host dynamics of relapse is important for understanding risk factors for recurrent malaria and temporal patterns of illness. It informs strategies for malaria control and elimination and methods of therapeutic assessment. We have used serial data from an intensively-followed large cohort of children studied on the North-Western border of Thailand, a region of low seasonal malaria transmission. Although the average *P. vivax* inoculation rate in this setting was estimated at approximately 0.5 infectious bites per person per year, this was sufficient to generate a high frequency of infection in younger children with consequent substantial morbidity.

We showed that a simple model of relapse based on independent constant rate activation dynamics for each hypnozoite was sufficient to capture most of the inter- and intra-individual variability in recurrence patterns. Under this simple model, the extremely detailed SPf66 vaccine trial data allowed confident estimation that the mean duration of carriage for a single hypnozoite is approximately 6 months. This model-dependent estimate was strongly informed by the patterns of successive recurrences observed in each individual, and was robust to mis-specification of the sporozoite batch size, age-stratification in the force of inoculation, and the ratio of immediately-developing sporozoites to hypnozoites.

A uniform force of inoculation did not explain the overall pattern of infections by age in the SPf66 trial. On the Thai-Myanmar border, anopheline vectors largely feed outdoors around dawn and dusk [46] so behavioural differences (e.g. greater mobility as children get older, or age-related outdoor activities like play, domestic labour and farming [47]) may have lead to differential exposures across age groups. Variation in body surface area [28] and the regularity of bed-net usage across age groups [48–50] may have been additional contributing factors to the conjectured lower force of inoculation among the youngest children.

The exponential clock model coupled with chloroquine suppression (but not elimination) of subsequent asexual parasite growth [33] creates a cyclical pattern of recurrent malaria. Although it is intrinsically “wasteful”, in that hypnozoites may activate at a time when subsequent asexual growth is suppressed by active infection (or antimalarial treatment) and so the parasite progeny do not reach transmissible densities, the random activation model requires sporozoite inocula which are still within the range considered likely in the natural setting. The model yields testable hypotheses about *P. vivax* biology (for example the mean duration of carriage) and provides a simple framework with which to assess the utility of public health interventions (for example interrupting transmission or MSAT).

In the SPf66 vaccine trial, both vivax and falciparum malaria were frequent in children, and mixed infections were recorded in 8.7% of cases. Most of the falciparum malaria cases were treated with artesunate-mefloquine which provided a long post-treatment suppression of subsequent *P. vivax* infections. However, by comparison with chloroquine, which suppresses but usually does not eliminate the relapse bloodstream infection [33], these data suggested that mefloquine does eliminate a proportion of the expected *P. vivax* relapses. This may be explained by the elimination profile of mefloquine which results in relatively high blood concentrations in the weeks following drug administration [51], whereas the chloroquine (and piperaquine) levels decline rapidly before plateauing at a relatively low level [33, 52]. This is a significant public health advantage for mefloquine which has not been appreciated previously.

In malaria endemic areas *P. falciparum* infections are commonly followed by *P. vivax* infections with an interval that is similar to the interval between a primary *P. vivax* infection and its subsequent relapse [17–20]. In this study mefloquine appears to have eliminated the blood stages *P. vivax* recurrence occurring within the first month so vivax malaria recurrences after falciparum malaria were less frequent. However in the minority of symptomatic falciparum episodes (*n* = 137) treated with artesunate monotherapy (limited post-treatment prophylaxis), we observed higher rates, consistent with previous studies indicating significantly different rates following slowly-vs rapidly-eliminated antimalarial treatment [17, 19, 20]. In particular, the rate of early *P. vivax* recurrence following artesunate monotherapy was much higher than could be explained by random hypnozoite activation. While these results are partially confounded by seasonality and do not provide definitive proof that hypnozoites can be activated from external triggering events such as febrile illness, this observation is compatible with external triggering. The model provides a general framework for characterising the importance of external triggering.

There are several factors which may have influenced the observed vivax malaria patterns in the SPf66 trial, and thus affect the interpretation of some inferred model parameters. The intensity of follow-up with daily participant visits and prompt antimalarial treatment would have attenuated any disease modification in hypnozoite activation risk. High quality microscopy ensured that all symptomatic malaria was identified, but presumably also captured incidental parasitaemias with other febrile illnesses. A key correlate of acute febrile malaria is thrombocytopemia (platelet count *<* 150, 000/mm^3^) [53]. In the SPf66 cohort, thrombocytopenia was only present in 34% of clinical consultations with a diagnosis of vivax monoinfection; 41% of consultations with a diagnosis of falciparum monoinfection and 42% of consultations with a diagnosis of mixed infection (Appendix A.3). This suggests that some recorded episodes may have indeed been asymptomatic parasitemias with coincident febrile illness. In addition, the daily assessment means that the duration of illness was nearly always less than 24 hours, which is unusually brief for malaria and shorter than in clinical trials of antimalarial treatment. This artificial situation, necessary for the vaccine trial, would have substantially attenuated any disease modification in hypnozoite activation risk. In therapeutic studies approximately half the relapses of *P. vivax* are not accompanied by fever, and recent studies in areas of similar transmission intensity in this region indicate high rates of asymptomatic parasitaemia — even in young children [54, 55]. Some of these are detectable by good microscopy. Primaquine was not given to prevent relapse, and children were visited each day. As such, a detected vivax parasitaemia in a febrile child could be a new episode of symptomatic vivax malaria, or it could represent asymptomatic parasitaemia and a coincidental viral infection. Even some falciparum malaria cases could have been asymptomatic parasitaemias and coincident viral infection. Although the population was aggregated together, there were no physical barriers. As such, movement, individual behaviours, geographic location and use of malaria preventive measures may have given rise to population heterogeneity in inoculation rates.

In conclusion, a simple model of *P. vivax* relapse assuming constant rate activation of liver hypnozoites was sufficient to explain the majority of variation in relapse patterns in a low transmission setting co-endemic for falciparum and vivax malaria. This does not exclude other processes such as disease activation, which would explain satisfactorily the elevated rate of *P. vivax* recurrence following symptomatic *P. falciparum* monoinfection, but for the most part does not require them. This model can be used to help assess the utility of public health interventions for the control and elimination of vivax malaria, and provides a methodology for the analysis of detailed prospective malariometric studies.

## Methods

### Data from the SPf66 vaccine trial

The peptide polymer SPf66 was amongst the earliest synthetic vaccine candidates for falciparum malaria [56]. Early field studies gave rise to inconsistent efficacy estimates [57–59]. Putative evidence of the ineffectiveness of the SPf66 vaccine against symptomatic malaria was provided by a randomised double-blind trial conducted between October 1993 and July 1995 on the Thailand-Myanmar border, a region co-endemic for *P. falciparum* and *P. vivax* [23]. In retrospect, this vaccine trial constitutes the most detailed cohort study of vivax malaria ever conducted.

### Study design

Over a study period spanning 21 months, daily home or school visits were conducted for 1344 children aged 2 to 15 years of age living in Shoklo camp — a community of refugees of Karen ethnic minority. Children exhibiting signs of illness were referred to outpatient clinics, where malaria was subsequently diagnosed using light microscopy. Absences from the camp were duly documented. Additional passive surveillance data were also collected, in thorough cross-sectional surveys conducted at two to three month intervals. We did not use data from the cross-sectional surveys in this analysis as the interpretation of an asymptomatic microscopic infection was limited by the proximity of the pyrogenic threshold for vivax infection (estimated to be *∼*150/*µ*L in a preceding study conducted in the camp between November 1991 and 1992 [46]) to the limit of detection of light microscopy. We focused instead on the sequence of symptomatic malaria episodes recorded for each child, assuming perfect detection of symptomatic episodes during periods of active clinical follow-up.

### Antimalarial treatment

Confirmed symptomatic cases of malaria were promptly treated with supervision; asymptomatic episodes detected through cross-sectional surveys were not treated. Typically, vivax monoinfections were treated with chloroquine (25 mg base/kg over 3 days), while uncomplicated falciparum monoinfections or mixed infections were treated with artesunate-mefloquine combination therapy (artesunate 4 mg/kg per day for 3 days and a single dose of mefloquine 25 mg base/kg on day 2 of treatment), with some exceptions; most notably, the administration of artesunate monotherapy to some children with hyperparasitemic falciparum malaria (see Appendix A.1 for details). Neither chloroquine nor mefloquine appear to have conferred any post-treatment protection against falciparum malaria, consistent with contemporaneous reports of widespread multidrug resistant *P. falciparum* [30]. Differential patterns of extended prophylactic protection against *P. vivax* were apparent following the administration of the slowly-eliminated antimalarials chloroquine and mefloquine: while chloroquine appeared to have given rise to a “prophylactic bunching” phenomenon, whereby residual drug levels (below the minimum inhibitory concentration) lead to the delayed manifestation rather than suppression of bloodstream infection [33], mefloquine appeared to have eliminated *P. vivax* bloodstream infections for an extended duration of time (Figure A.1, Appendix A.1). We adjusted for post-treatment prophylaxis by defining a fixed period of prophylactic protection for each drug regimen, during which all emerging bloodstream infections are eliminated. This was assumed to span 15 days, with two exceptions: chloroquine monotherapy was assumed to provide no protection against falciparum, while mefloquine was assumed to eliminate all emerging *P. vivax* bloodstream infections for an extended period of 35 days. Any consultation that overlapped with the prophylactic protection period of a previous episode was flagged as a possible treatment failure (see Appendix A.1.4 for details).

### Incidence of symptomatic malaria

We computed the incidence of symptomatic malaria after removing likely treatment failures and adjusting the person-time at risk for periods of prophylactic protection (in addition to left/right-censoring and documented absences from the camp). To assess age structure, we calculated age-stratified incidence rates (i.e. total number of symptomatic episodes divided by the cumulative person-years at risk for each age group) with bootstrap resampling. We applied the Kruskall-Wallis test and the non-parametric Mack-Wolfe test for umbrella alternatives (with unknown peak) [60] to the incidence per individual (i.e. the number of symptomatic episodes divided by the time at risk) within each age group (Appendix A.4).

### A theoretical within-host framework for hypnozoite accrual

We embedded the exponential clock model of White et al. [22] in a stochastic framework of sporozoite inoculation from repeated mosquito bites. This model was formulated as an open network of infinite server queues, generalising the construction in [25] to capture sporozoite “destiny” (immediate development vs hypnozoite formation). The original model in [22] accommodated a constant death rate for each hypnozoite to capture hepatocyte turnover. Since hypnozoite death is unidentifiable on the basis of temporal recurrence data, we formulated a reparameterised system restricted to successful sporozoites — that is, sporozoites that either undergo immediate development or form hypnozoites that eventually activate to give rise relapses (see Appendix B.1 for details of this reparameterisation, and an explicit link to [25]).

### Model structure

The construction of the within-host framework, illustrated in Figure 8, is as follows:

- The timing of infective bites is modelled with a non-homogeneous Poisson process characterised by the time-dependent rate *λ*(*t*).
- The number of *successful* sporozoites *S* established by each infective bite is modelled to follow a geometric distribution, with mean *ν* and state space ℤ*_≥_*_0_, that is, probability mass function (PMF)

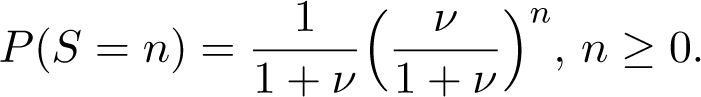
- With probability *p*_rel_, each (successful) sporozoite forms a hypnozoite that is destined to activate (state *H*); otherwise, it undergoes immediate development.
- The immediate development of one or more sporozoites within a batch gives rise to a primary infection (state *P*).
- The time to activation for each hypnozoite — or equivalently, the duration of hypnozoite carriage — is modelled to be exponentially-distributed with mean 1*/η* (with hypnozoite activation represented by a transition from state *H* to *A*).

**Figure 8:**
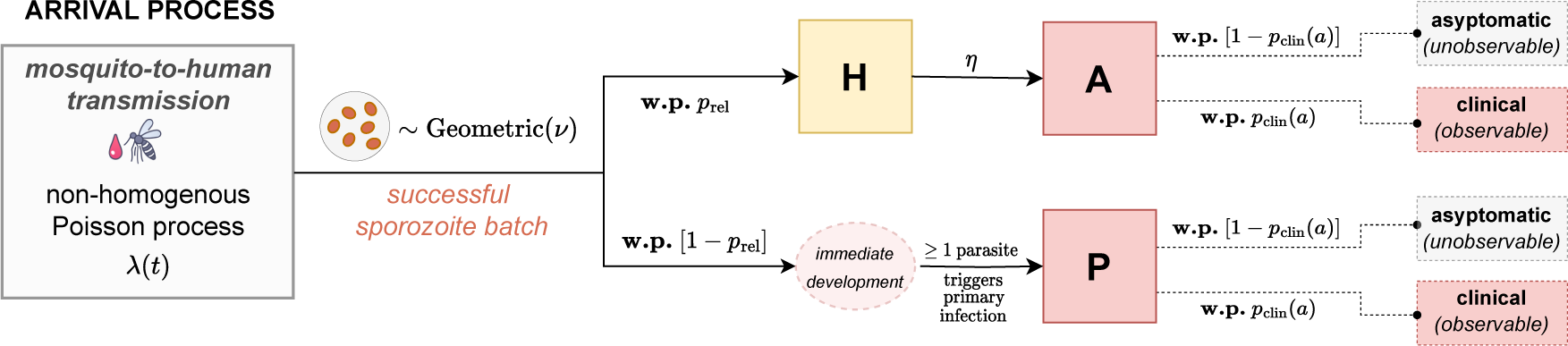
Schematic of the open network of infinite server queues used to model the burden of blood-stream infection (see Appendix B.1 for a link to [25]). Adapted from Figure 1 of Mehra et al. (2022), *arXiv:2208.10403*. **w.p.**: with probability.

Treating age as a proxy for previous malaria exposure, anti-disease immunity is modelled as an age-dependent masking mechanism. For an individual of age *a* at enrolment, each immediately-developing sporozoite batch (i.e. arrival into node *P*) and hypnozoite activation event (i.e. arrival into node *A*) in the study period is assumed to give rise to clinical symptoms with some probability *p*_clin_(*a*); the time to manifestation of clinical symptoms is not modelled. We assume the functional form

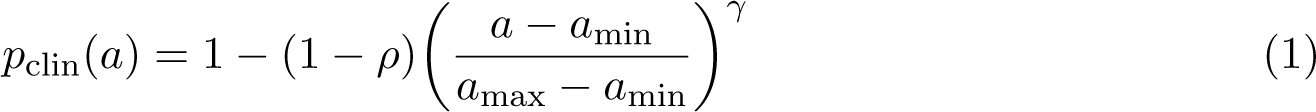

whereby

1. *p*_clin_(*a*_min_) = 1 for the youngest age group in the study (*a*_min_ = 2 year olds);
2. the parameter *ρ* corresponds to the proportional reduction in the probability that a blood-stream infection will be symptomatic across the age range of the study (i.e. between *a*_min_ = 2 and *a*_max_ = 15 years of age); and

- the exponent *γ* modulates the steepness/shape of the (monotonically decreasing) curve, which is concave up in the case *γ <* 1, linear in the case *γ* = 1 and concave down if *γ >* 1.

The proportion of symptomatic infections in the 2 year old group is not identifiable^3^ but we assume it is 1. We therefore interpret *p*_clin_ is the probability that a bloodstream infection is symptomatic *relative* to the average 2 year old (the reference group). If a large proportion of cases in 2 year olds were in fact asymptomatic, then we would underestimate the sporozoite batch size.

### Analytical derivation of metrics of epidemiological interest

Metrics of epidemiological interest were derived analytically under this stochastic within-host framework (Appendix E). For simplicity, these results assume that the hypnozoite reservoir has reached stationarity under a constant force of inoculation *λ* (in the time period before the study started; this has very little impact on the parameter inference). In this setting, the hypnozoite burden can be shown to follow a negative binomial distribution with mean *λνp*_rel_*/η* and variance *λνp*_rel_(1 + *νp*_rel_)*/η* [25]. To achieve spontaneous hypnozoite clearance with probability *c* in each individual, mosquito-to-human transmission would therefore need to be interrupted for time

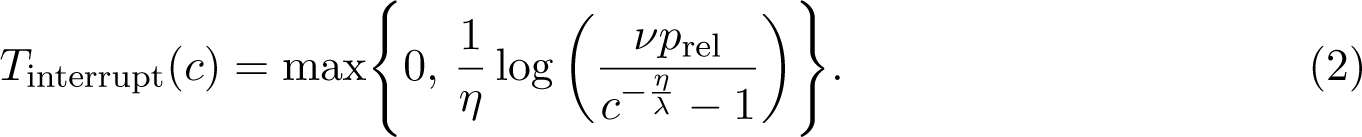

The probability of recurrence (i.e. at least one hypnozoite activation and/or immediate sporozoite development event) in any window of length *W* takes the form

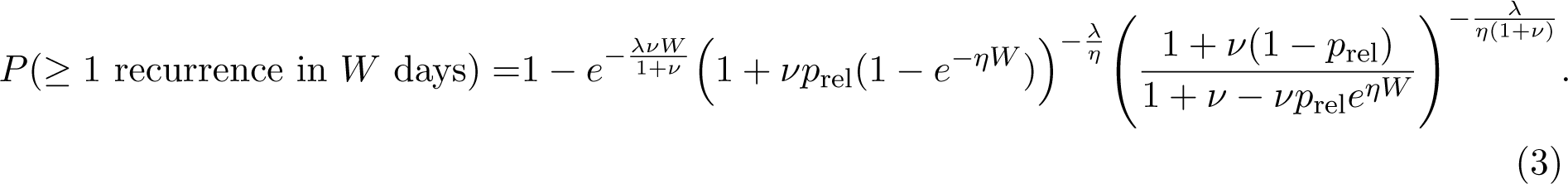

In addition, the specificity of bloodstream infection within the previous *W* days as a predictor of hypnozoite carriage is:

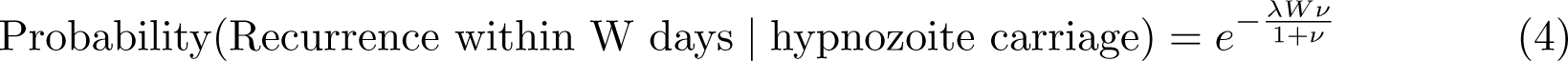

This decays exponentially with the force of inoculation *λ*. Given an imperfect serological test with sensitivity *r*_sens_ and specificity *r*_spec_ for detecting bloodstream infection in the previous *W* days, the fold reduction in over-treatment for MSAT versus MDA (in the eligible population) can be written

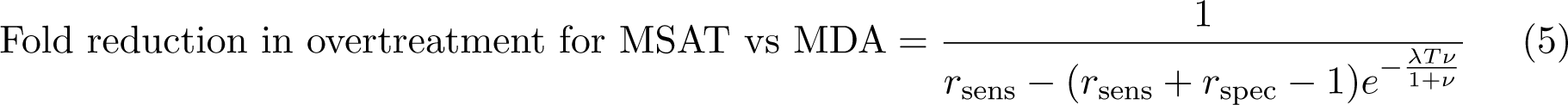

assuming the outcome of the serological test given recent recurrence is conditionally independent to hypnozoite carriage. Accommodating population heterogeneity by randomly sampling the force of inoculation for each individual from a Gamma distribution [44] is also analytically tractable.

### An inferential framework tailored to the SPf66 vaccine trial

We constructed an inferential framework, tailored to the SPf66 dataset, that allowed us to compute the exponential clock model likelihood for multiple observed recurrent vivax episodes. For this to be tractable we discretised the study period into *n*_obs_ = 65 windows, each of length *T* = 10 days. The force of inoculation for *P. vivax* was assumed to be piecewise constant over each window, enabling the derivation of an analytic likelihood and substantially simplifying inference. Age is the key covariate in the model, serving as a determinant of both the baseline hypnozoite burden (since it determines the period of time over which the reservoir is permitted to accrue before the start of the study) and the degree of anti-disease masking.

### Context-dependent observational model

#### Discretised vivax infection states adjusted for post-treatment prophylaxis

In addition to anti-disease immunity (which is a structural feature of the within-host model), hypnozoite activation and/or immediate sporozoite development events may have been masked (i.e. unobserved) due to either censoring or post-treatment prophylaxis. For each child, we constructed a binary sequence of clinical infection states, indicating whether or not a symptomatic vivax episode was recorded for each 10 day window. To account for censoring, we masked any windows in which a child was actively followed-up for 5 days or less (due to either left- or right-censoring, or a documented absence from the camp). We additionally modelled post-treatment prophylaxis as follows (with some exceptions in the vicinity of possible treatment failures, see Appendix A.5.1 for details). For every window *i* for which antimalarial treatment was administered, we masked window (*i* + 1); if mefloquine was administered, we additionally masked windows (*i* + 2) and (*i* + 3) to allow for an extended period of prophylactic protection (i.e. we assumed that merozoites released from the liver during this period were unable to successfully establish bloodstream infections because drug levels were high enough to eliminate the asexual parasites resulting from hepatic schizogony [between 10,000 to 100,000 parasites]). If chloroquine was administered in window *i* and a symptomatic vivax episode was recorded in window (*i* + 3), then we grouped together windows (*i* + 2) and (*i* + 3) (i.e. we allowed for the potentially delayed manifestation of clinical symptoms following any hypnozoite activation or immediate sporozoite development events during this period, due to an asexual parasite multiplication rate that was temporarily dampened by residual drug levels but still exceeded one [33]). Discretised vivax infection states for children aged 7 at enrolment are visualised in Figure A.5, Appendix A.5.1.

#### Seasonality in the force of inoculation

We used the incidence of symptomatic falciparum malaria as a direct proxy of the *P. vivax* mosquito inoculation rate. To infer relative seasonal fluctuations in the force of inoculation for *P. vivax* over the study period, we fitted a smoothing spline to the incidence of symptomatic falciparum malaria calculated in 20 day windows, with a moving average across half-windows (see Appendix A.5.2 for details); in the main model fit we then estimated a scaling factor governing the ratio of vivax to falciparum inoculation rates (this was done as a two-stage process without uncertainty propagation). The force of inoculation for vivax relative to falciparum was estimated separately before and after April 1994 (day 200 of the study period), to account for variation in the relative burden of vivax vs falciparum malaria following the camp-wide shift in treatment regimen for falciparum malaria [31, 32]. We assumed a constant force of inoculation prior to the study period i.e. seasonality and systematic changes in vivax transmission in the years preceding the study were not modelled. We did not account for heterogeneity or age-stratification in the force of inoculation (the impact of age-stratification was assessed in a sensitivity analysis, Appendix C.2.3).

#### Analytic model likelihood

Under the theoretical framework, for an individual of age *a* at enrolment, we modelled hypnozoite accrual for duration *a*, before considering the number of hypnozoite activation and immediate development events *N_i_*(*a*) in each window *i* = 1*, . . ., n*_obs_ of the study period. We characterised the joint distribution of **N**(*a*) from first principles, by deriving a multivariate probability generating function (PGF) (Appendix B.2). Our argument mimics the approach of [25, 61–63] and others: we condition first on the time course of each sporozoite, followed by the size of an incoming sporozoite batch and finally, the sequence of bite times. For an individual of age *a*, the likelihood of clinical symptoms in a set of windows *I* (which may be aggregated due to prophylactic bunching) takes the form

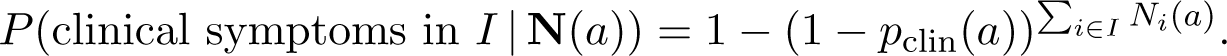

Application of the inclusion-exclusion principle to the multivariate PGF of **N**(*a*) allows the likelihood of a given sequence of symptomatic episodes to be recovered analytically (see Appendix B.2 for details). The time complexity of evaluating the likelihood scales exponentially with the number of symptomatic episodes experienced by each child; computational constraints, coupled with numerical instability arising from catastrophic cancellation, yield a practical upper bound of 13 symptomatic episodes per child for our implementation, which is the maximum observed in the SPf66 trial.

#### Bayesian inference using the Metropolis-Hastings algorithm

We performed Bayesian inference using the Metropolis-Hastings algorithm. The proportion of hypnozoites (vs immediately developing forms) was set to *p*_rel_ = 0.4, informed by recent *in vivo* and *in vitro* experiments on the Chesson strain of *P. vivax* [34]. We thus generated estimates for 6 parameters: the hypnozoite activation rate *η*; the mean sporozoite batch size *ν*; the average force of inoculation Λ_1_ and Λ_2_ before and after day 200 of the study period respectively; and the parameters *ρ* and *γ* governing the age-dependent anti-disease masking probability (Equation (1)). We used flat improper priors on the non-negative real line for the parameters *{*Λ_1_, Λ_2_, ν, η} but informative normal priors

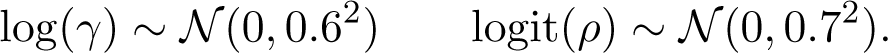

The prior distribution for the probability of symptomatic infection *ρ* at age 15 relative to age 2 had median value 0.5, with 95% mass in the interval [0.2, 0.8]. The prior distribution for the shape parameter log(*γ*) of the anti-disease masking curve was symmetric around zero (corresponding to linear decline), with 95% mass in the interval [-1.18, 1.18]; applying the transformation log(*γ*) *1→ −* log(*γ*) reflects the age-dependent anti-disease masking probability along the line of identity. We implemented a symmetric (rectified) normal proposal distribution. Details are provided in Appendix C.1.

#### Posterior predictive checks

To perform posterior predictive checking, we simulated symptomatic vivax episodes under 2000 parameter combinations sampled from the posterior uniformly at random (without replacement). Simulated cohorts had the same age structure as the SPf66 trial. The sequence of hypnozoite activation and immediate sporozoite development events under the exponential clock model was obtained through direct stochastic simulation (Appendix C.2.1). We adjusted for anti-disease masking to recover a discretised sequence of binary clinical infection states over *T* = 10 day windows. We then imposed an observational model, accounting for lapses in clinical follow-up (informed by patterns of left- and right-censoring, and documented camp absences in the SPf66 cohort) and post-treatment prophylaxis (with identical parameters to chloroquine). *P. falciparum* episodes were not simulated. We compared age structure and seasonal fluctuations in the incidence of symptomatic vivax malaria for these simulated cohorts against observed data.

#### Vivax after falciparum monoinfection

To interrogate whether the exponential clock model could explain observed rates of vivax recurrence following falciparum monoinfection, we derived the probability of observing a symptomatic vivax recurrence in 20 to 80 day windows following each falciparum monoinfection (due to either spontaneous hypnozoite activation and/or immediate sporozoite development). In doing so, we conditioned on the recorded history of vivax recurrence prior to the baseline falciparum monoinfection. We additionally accounted for censoring due to lapses in clinical follow-up and post-treatment prophylaxis following the treatment of falciparum malaria in each relevant follow-up window. Given the resultant set of probabilities, we modelled the number of falciparum monoinfections followed by a vivax recurrence within the window using a Poisson binomial distribution (i.e. as a sum of independent, but non-identically distributed Bernoulli random variables). We computed 99% credible intervals for the proportion of falciparum monoinfections followed by vivax recurrence based on 2000 parameter combinations sampled uniformly at random from the posterior. Technical details are provided in Appendix C.3.

We stratified baseline episodes based on treatment with artesunate monotherapy vs artesunate-mefloquine combination therapy, given empirical evidence that early emerging bloodstream infections were suppressed following the administration of mefloquine (see Kaplan-Meier curves for time to vivax recurrence following falciparum monoinfection, Figure A.2). We removed falciparum monoinfections in the vicinity of treatment failure (Appendix A.1.4) and considered baseline episodes up until April 1995 (day 570 of the study, to yield adequate clinical follow-up) leaving *n* = 120 falciparum monoinfections treated with artesunate monotherapy and *n* = 607 treated with artesunate-mefloquine combination therapy. For doubly-robust analysis, we additionally matched the treatment groups by the number of previous vivax episodes and the time of the baseline falciparum monoinfection (to control for seasonality) using the nearest neighbour method implemented in the R function MatchIt::match.data [64].

#### Sensitivity to model misspecification

To assess the sensitivity of our parameter estimates to model misspecification, we conducted several supplementary analyses. Sporozoite inocula may be over- or under-dispersed relative to the geometric distribution. We thus performed a series of simulation studies, whereby data were simulated with negative binomial sporozoite batches, but inference was performed under the assumption of geometric sporozoite batches (Appendix D.1). Additionally, the ratio of hypnozoites to immediately-developing forms for the Chesson strain [34] may not be representative of South East Asian strains. As such, we conducted a similar sensitivity analysis for the hypnozoite fating probability *p*_rel_ (Appendix D.2). In the absence of clear age structure in the incidence of symptomatic falciparum infection (Figure A.4, Appendix A.4), it is reasonable to assume there was no age stratification in the mosquito inoculation rate for 2 to 15 year olds; however, infants under the age of 2 may have been subject to lower exposure [37–39]. To accommodate potential age-stratification in mosquito inoculation for infants *<* 2 years vs children *>* 2 years, we re-calibrated the model with the force of inoculation for infants scaled down by a series of fixed factors (Appendix C.2.3).

### Data and code availability

All data and code are openly available on a GitHub repository: https://github.com/somyamehra/SPf66.

## Data Availability

All data relevant to the study are openly available on a GitHub repository.

https://github.com/somyamehra/SPf66

## Acknowledgements

NJW is a Principal Research Fellow funded by the Wellcome Trust (093956/Z/10/C). JAW is a Sir Henry Dale Fellow funded by the Wellcome Trust (223253/Z/21/Z). SM acknowledges funding from the Australian Research Council through Laureate Fellowship FL130100039.

This research was funded by Wellcome. A CC BY or equivalent licence is applied to the author accepted manuscript arising from this submission, in accordance with the grant’s open access conditions.

## Footnotes

1 In individuals not enrolled in the SPf66 trial, uncomplicated falciparum malaria in the camp was largely treated with mefloquine monotherapy until early 1994, with associated high rates of treatment failure [31, 40]: this biases our estimates of seasonality in mosquito inoculation, and seasonal fluctuations in the incidence of symptomatic vivax malaria are poorly captured, particularly before April 1994 (see Figure C.3, Appendix C.2.2).

2 Since artesunate monotherapy was typically administered to hyperparasitemic children, there may be systematic variation in prior exposure and seasonal confounding between these groups. Matching can correct for this confounding.

3 If we restrict our attention to the subset of the hypnozoite reservoir that is destined to activate, then an individual of age *a* can be thought to accrue hypnozoite batches of mean size *νp*_rel_*p*_clin_(*a*) over a period of time *a*. As such, the proportion of symptomatic infections in the baseline age group is not structurally identifiable on the basis of data for symptomatic *relapse* alone. Since we expect relapse to constitute the majority of the bloodstream infection burden, practical identifiability issues would plausibly persist despite the additional contribution of primary infection.

