## Supporting Information for "Modelling the within-host dynamics of *Plasmodium vivax* hypnozoite activation: an analysis of the SPf66 vaccine trial"

**Summary**

In this supplement, we provide technical details of our theoretical and statistical analysis. Supplementary details of the SPf66 vaccine trial, including an overview of antimalarial treatment, seasonality and age structure in symptomatic episodes, are presented in Appendix A. Appendix B concerns the stochastic within-host framework in which we embed the exponential clock model, including the derivation of an analytic likelihood for multiple recurrent infections. In Appendix C, we address the calibration of the model to clinical recurrence data from the SPf66 vaccine trial, encompassing parameter estimation using the Metropolis-Hastings algorithm and posterior predictive checks for seasonality and age structure. The sensitivity of parameter estimates to model misspecification — namely, parametric misspecification of the sporozoite batch size distribution and a hypnozoite fating probability $p_{\text{rel}}$ diverging from the Chesson strain — is discussed in Appendix D. Analytic expressions for various quantities of epidemiological interest, with accompanying derivations, are provided in Appendix E. Appendix F is devoted to a discussion a of the calibrated model, centred on the 8 theses of vivax relapse biology posited by White [1].

**F Theses of vivax relapse biology** **64**

### Appendix A

### Data from the SPf66 vaccine trial

#### A.1 Antimalarial treatment

##### A.1.1 Treatment regimens

Standard treatment regimens for the SPf66 cohort were as follows [3]:

- *Vivax monoinfection*: chloroquine 25 mg base/kg total dose over 3 days.
- *Uncomplicated falciparum moninfection or mixed infection*: artesunate 4 mg/kg per day for 3 days and a single dose of mefloquine 25 mg base/kg on day 2 of treatment.
- *Severe and complicated malaria*: artemether, commencing with an intramuscular dose 3.2 mg/kg on day one and then 1.6 mg/kg daily, followed by artesunate-mefloquine combination treatment, as for uncomplicated falciparum malaria.

However, antimalarial treatment records diverge from these standard regimens on occasion and are summarised in Table A.1 for completeness. Notably, artesunate monotherapy was administered in 15.5% ($n = 137$ of $n = 884$) of consultations with a falciparum monoinfection diagnosis, typically to hyperparasitaemic patients.

##### A.1.2 Post-treatment prophylaxis

We formulate a model of post-treatment prophylaxis informed by observed inter-consultation intervals, stratified by diagnosis and treatment at baseline.

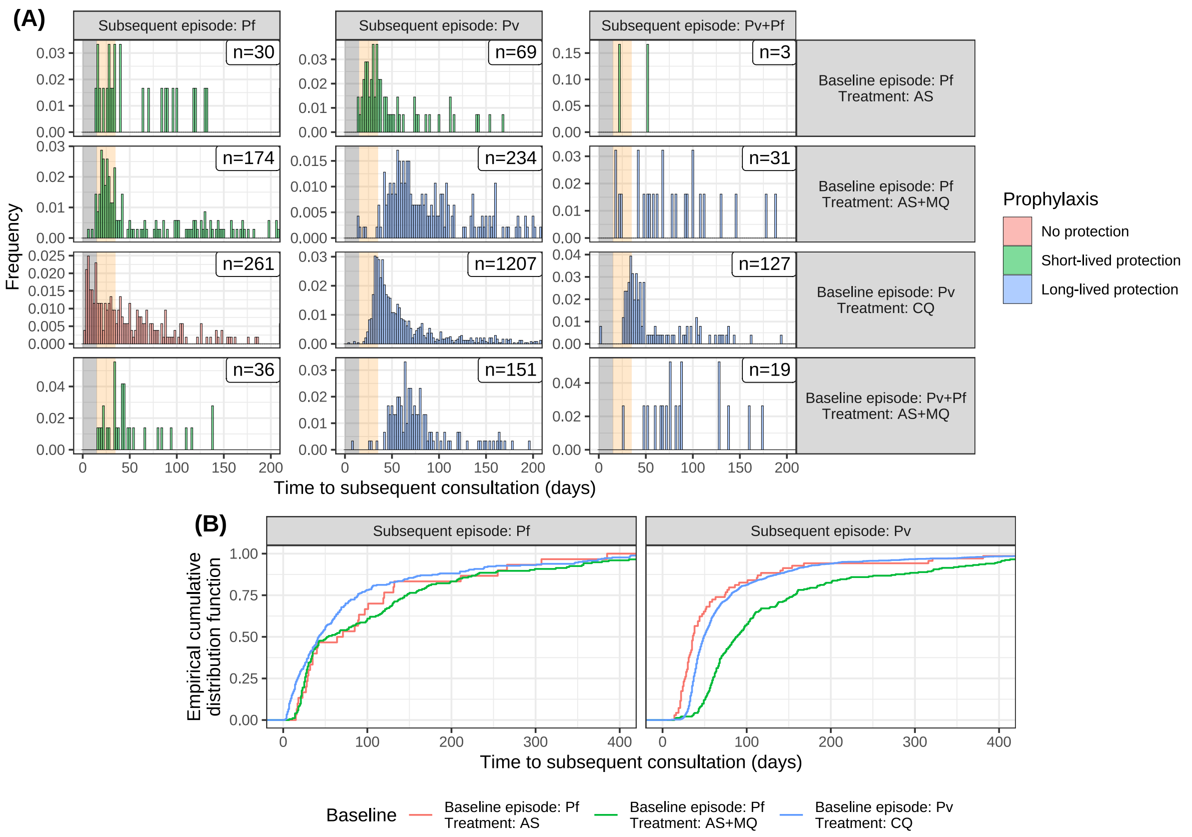

**Figure A.1:** Inter-consultation intervals (prior to screening treatment failure) following standard antimalarial treatment i.e. chloroquine monotherapy (CQ) for vivax monoinfection; artesunate monotherapy (AS) for falciparum monoinfection or artesunate-mefloquine combination therapy (AS+MQ) for either falciparum monoinfection or mixed infections). For a child with $\ell$ clinical consultation records, we report $(\ell - 1)$ inter-consultation intervals, stratified by baseline treatment and diagnosis. Frequency distributions (truncated at 200 days but annotated with total counts) are shown in Panel A, while empirical cumulative distribution functions are shown in Panel B.

| Diagnosis | Recorded treatment | No. consultations |
|---|---|---|
| *Pv monoinfection* | Chloroquine monotherapy | 2180 |
| | Artesunate-mefloquine | 35 |
| | Artesunate monotherapy | 9 |
| *Pf monoinfection* | Artesunate-mefloquine | 720 |
| | Artesunate monotherapy | 137 |
| | Quinine-tetracycline | 9 |
| | Artemether | 7 |
| | Artesunate-tetracycline | 5 |
| | Mefloquine monotherapy | 3 |
| | Quinine monotherapy | 2 |
| | Chloroquine monotherapy | 1 |
| *Pv+Pf mixed infection* | Artesunate-mefloquine | 266 |
| | Artesunate monotherapy | 18 |
| | Chloroquine monotherapy | 5 |
| | Quinine-tetracycline | 2 |
| | Artesunate-tetracycline | 2 |
| | Artemether | 1 |

**Table A.1:** Number of recorded clinical consultations (prior to screening treatment failure), stratified by diagnosis (via light microscopy) and antimalarial treatment regimen.

###### A.1.2.1 Protection against *P. falciparum*

Chloroquine does not appear to confer any protection against falciparum monoinfection, with an immediate risk of falciparum monoinfection following chloroquine monotherapy (Figure A.1A). Additionally, comparison of inter-consultation intervals following artesunate monotherapy versus artesunate-mefloquine combination therapy (Figure A.1B) suggest that mefloquine did not confer extended protection against *P. falciparum*. These observations are concordant with contemporaneous reports of widespread multidrug resistant *P. falciparum* [4].

###### A.1.2.2 Protection against *P. vivax*

We see differential patterns of extended prophylactic protection against *P. vivax* following the administration of chloroquine versus mefloquine, both of which are slowly-eliminated. Chloroquine is known to give rise to a 'prophylactic bunching' phenomenon, whereby residual drug levels (below the minimum inhibitory concentration) lead to the delayed manifestation, rather than supression, of bloodstream infection [5]. Intervals between chloroquine-treated vivax monoinfec-

| Diagnosis | Treatment | Masking | Bunching |
|---|---|---|---|
| *P. vivax* | Chloroquine monotherapy | 15 days | 20 days |
| | Artesunate-mefloquine | 35 days | 0 days |
| | Artesunate monotherapy | 15 days | 0 days |
| | Other (no mefloquine) | 15 days | 0 days |
| *P. falciparum* | Chloroquine monotherapy | 0 days | 0 days |
| | Artesunate-mefloquine | 15 days | 0 days |
| | Artesunate monotherapy | 15 days | 0 days |
| | Other | 15 days | 0 days |

**Table A.2:** Prophylactic masking and bunching periods for each drug regimen.

tions exhibit a unimodal characteristic, with a mode of approximately 35 days and a comparatively steep rise from 25 to 35 days (Figure A.1A). While this characteristic is influenced by temporal auto-correlation in vivax recurrences (recent recurrence is a predictor of hypnozoite carriage and consequently the risk of subsequent relapse), it is qualitatively consistent with prophylactic bunching. In contrast, comparison of inter-consultation intervals following artesunate monotherapy versus artesunate-mefloquine monotherapy (Figure A.1B) suggests that mefloquine leads to the suppression, rather than just the delay, of vivax recurrence for an extended duration of time.

##### A.1.3 Modelling post-treatment prophylaxis

For each drug regimen, we model post-treatment prophylaxis in two stages, each of fixed duration:

- An initial prophylactic 'masking' period, during which merozoites emerging from the liver are unable to establish bloodstream infection i.e. any hypnozoite activation and/or immediate development events are masked;
- A subsequent prophylactic "bunching" period, during which the manifestation of bloodstream infection is potentially delayed i.e. any hypnozoite activation and/or immediate development events within this window have a potentially delayed manifestation at the end of the bunching window.

Bunching and masking periods, stratified by drug regimen and diagnosis, are detailed in Table A.2.

##### A.1.4 Screening treatment failure

Any consultation falling within the masking period (Table A.2) of a previous consultation is flagged as a possible treatment failure; 33 such consultations are shortlisted. We handle these cases as follows:

- If the malaria diagnosis is consistent across a flagged sequence of treatment failures and the preceding episode (there are $n = 14$ such sequences, implicating $n = 15$ flagged consultations), then we retain only the first episode but extend out the prophylactic masking period from the time of the initial consultation, to the end of the prophylactic protection period following the final consultation.
- If a diagnosis of vivax monoinfection is followed by a diagnosis of mixed infection within 3 days or less (there are $n = 2$ such cases), then we remove the initial vivax episode.
- If the initial diagnosis is a falciparum monoinfection that is either
  - followed by a diagnosis of vivax monoinfection ($n = 7$ flagged consultations)
  - followed by a diagnosis of mixed infection ($n = 5$ flagged consultations), but after the falciparum prophylactic protection period for the previous consultation has lapsed

  then we retain both episodes but set the period of prophylactic protection against vivax malaria to be zero following treatment of the initial episode.
- If the initial diagnosis is a mixed followed by a diagnosis of vivax monoinfection ($n = 4$ flagged consultations), then we retain only the initial mixed infection but extend out the prophylaxis masking period for vivax malaria from the time of the initial consultation, to the end of the prophylactic protection period following the final consultation.

After masking treatment failures, we retain 2211 of the 2224 vivax monoinfections; 871 of 884 falciparum monoinfections; and 293 of 294 mixed infections. The time at risk for each child is adjusted for prophylactic masking, in addition to left- and right-censoring, and documented absences from the camp.

#### A.2 Vivax after falciparum monoinfection

Of particular interest is the rate of vivax malaria following febrile falciparum malaria [6–9]. Kaplan-Meier survival curves from the time from each recorded falciparum monoinfection to the

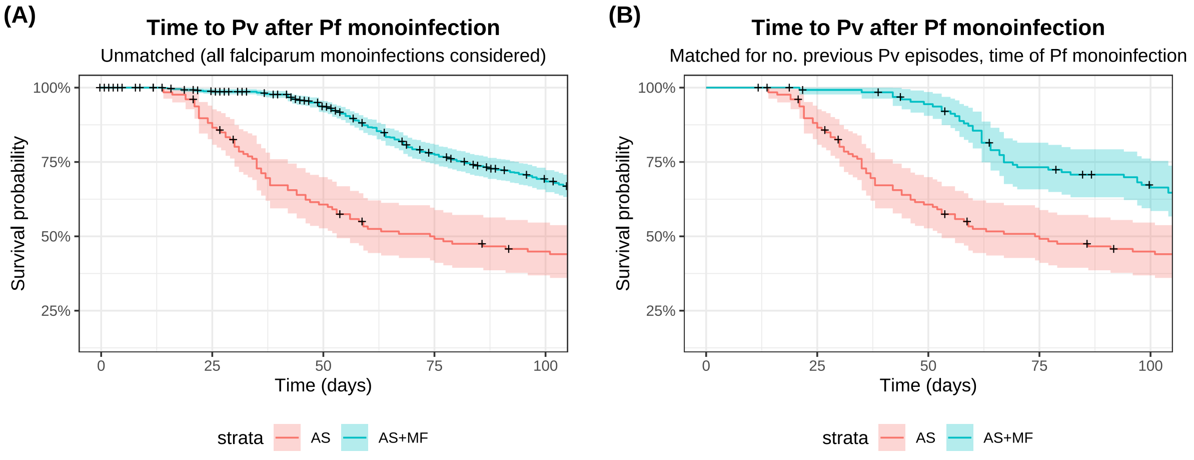

**Figure A.2:** Kaplan-Meier survival curves for the time to vivax infection following falciparum monoinfection, stratified by treatment with artesunate monotherapy (AS) vs artesunate-mefloquine combination therapy (AS+MQ) *prior* to screening treatment failure. Panel A shows results for all AS+MQ-treated falciparum monoinfections. Panel B shows results for a subset of AS+MQ-treated falciparum monoinfections, that have been matched to AS-treated falciparum monoinfections for the number of previously recorded vivax episodes and the time of the baseline falciparum monoinfection.

subsequent symptomatic vivax episode are shown in Figure A.2. We adjust for right-censoring, but not interruptions in clinical follow-up due to documented absences from the camp. We stratify baseline episodes by treatment with either artesunate monotherapy, or artesunate-mefloquine combination therapy. Since artesunate monotherapy was typically administered to children with repeated falciparum episodes (to treat hyperparasitaemic patients), we match the treatment groups by the number of previous vivax episodes and the time of the baseline falciparum monoinfection (to control for seasonality) using the nearest neighbour method implemented in the R function `MatchIt::match.data` [10]. This should remove confounding by seasonality and prior exposure. Survival analysis has been performed using the R function `survival::survfit` [11].

Comparison of survival curves following artesunate monotherapy vs artesunate-mefloquine combination therapy, after matching for the history of vivax malaria and seasonality (Figure A.2B), suggests that mefloquine eliminates bloodstream infections which emerge in the first month after treatment.

| Diagnosis | $< 150,000/\text{mm}^3$ | $150,000 - 200,000/\text{mm}^3$ | $> 200,000/\text{mm}^3$ |
|---|---|---|---|
| *Pf monoinfection* | 40.9% | 27.6% | 31.5% |
| *Pv monoinfection* | 34.0% | 31.0% | 35.0% |
| *Pv+Pf monoinfection* | 41.6% | 29.5% | 28.8% |

**Table A.3:** Summary of platelet counts recorded at each consultation (prior to screening treatment failure).

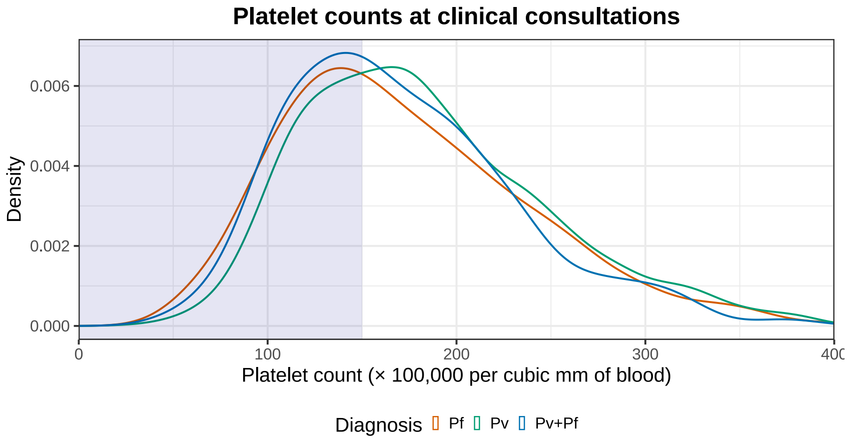

**Figure A.3:** Platelet counts recorded at each consultation (prior to screening treatment failure), stratified by diagnosis. The threshold for thrombocytopenia (platelet count below 150,000/mm$^3$) is highlighted in blue.

#### A.3 Platelet counts

Thrombocytopenia is a feature of all human clinical malaria infections (platelet count < 150,000/mm$^3$) [12]. Platelet counts exceeding 200,000/mm$^3$ are unusual for symptomatic malaria. Density plots of platelet counts recorded at each clinical consultation are shown in Figure A.3, and summarised in Table A.3. The relatively large proportion of consultations with normal platelet counts suggests that some symptomatic malaria episodes may have been asymptomatic parasitemias with coincident febrile viral infections.

#### A.4 Observed age structure in the incidence of symptomatic malaria

Here, we summarise observed age structure in the incidence of symptomatic malaria in the SPf66 cohort. Mixed infections are double-counted as both vivax and falciparum episodes. We define age groups based on the age at enrolment (2 to 15 years).

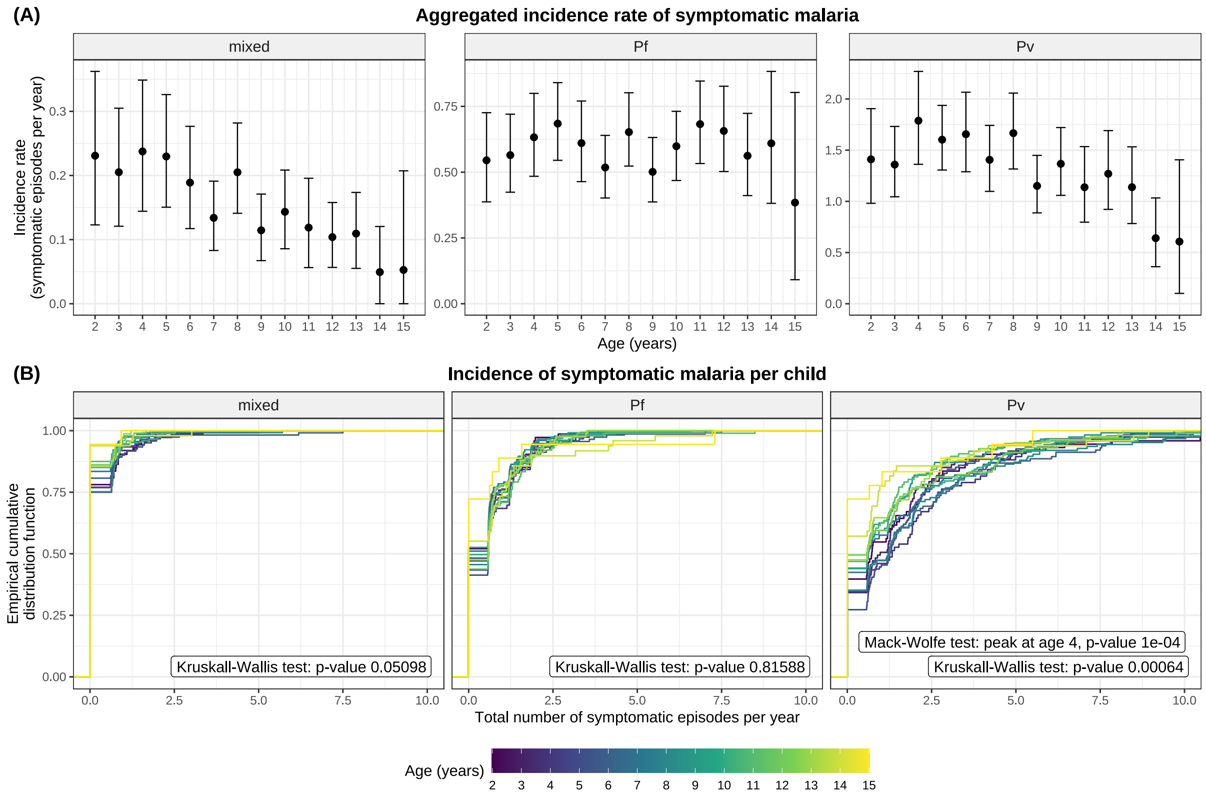

**Figure A.4:** Overview of age structure in the incidence of symptomatic malaria in the SPf66 cohort.

(A) The aggregated incidence rate stratified by age group. Error bars depict 95% confidence intervals generated through bootstrap resampling with 2000 replicates.

(B) Age-stratified empirical cumulative distribution functions (CDFs) for the incidence per individual $Y_q^{(\ell)}/T_q^{(\ell)}$, annotated with $p$-values for the Kruskall-Wallis rank sum test and the Mack-Wolfe test (with unknown peak) for umbrella alternatives [13].

For each child $\ell$, of age $a^{(\ell)}$, and each possible diagnosis $q \in \{\text{Pf, Pv, mixed}\}$, we compute the total number of symptomatic episodes $Y_q^{(\ell)}$ with diagnosis $q$ (after screening for treatment failure) and the cumulative time at risk $T_q^{(\ell)}$ (adjusting for left- and right-censoring; any documented absences from the camp and prophylactic masking).

The aggregated incidence rate for a particular age group $a$ is computed as the quotient

$$I_q(a) = \frac{\sum_\ell \mathbb{1}_{\{a^{(\ell)}=a\}} Y_q^{(\ell)}}{\sum_\ell \mathbb{1}_{\{a^{(\ell)}=a\}} T_q^{(\ell)}}, \tag{A.1}$$

and visualised in Figure A.4A. We generate 95% confidence intervals through bootstrap resampling with 2000 replicates.

The incidence per individual is $Y_q^{(\ell)}/T_q^{(\ell)}$, and stratified by age group (Figure A.4B). We conduct non-parametric rank sum tests to screen for age structure in the incidence per individual: the Kruskall-Wallis test [13] is performed using the R function `stats::kruskal.test` [14], while the Mack-Wolfe test for umbrella alternatives (with unknown peak) [13, 15] is performed using the R function `PMCMRplus::mackWolfeTest` [16], with $p$-values estimated from bootstrap permutations with 10000 iterations.

#### A.5 Pre-processing for model calibration

##### A.5.1 Discretised vivax infection states

For the purpose of model calibration, we discretise the study period into $n_{\text{obs}} = 65$ windows, each of length $T = 10$ days. Prior to applying the prophylactic masking and bunching periods detailed in Table A.2, we shift each recorded clinical consultation to the midpoint of the relevant window. Each window is assigned one of four possible states:

- *Masking* (state $M$): Hypnozoite activation and/or immediate sporozoite development events can go unobserved due to lapses in clinical follow-up (due to left/right-censoring or a documented absence from a camp) or prophylactic protection (whereby merozoites that emerge from the liver are unable to establish bloodstream infection). We calculate the total number of days within each window that are censored either due to interruptions in active clinical follow-up or prophylaxis (i.e. overlap with the prophylactic masking period of a previous consultation). If more than 5 days of a given window are censored in this manner, then we mask observations in the relevant window. In most cases, if antimalarial treatment is administered in window $i$, then to adjust for post-treatment prophylaxis, we mask observations in window $(i+1)$; and additionally windows $(i+2)$ and $(i+3)$ in the event of mefloquine treatment (Table A.2). However, prophylactic masking periods are adjusted for treatment failure in a small number of cases (Appendix A.1.4).
- *Clinical consultation* (state $C$): This indicates a window in which a clinical consultation was recorded, and which does not overlap for more than 5 days with the prophylactic

bunching period of a previous consultation; in effect, a hypnozoite activation and/or immediate sporozoite development event is assumed to give rise to a symptomatic bloodstream infection without delay in this window.

- *Bunching* (state $B$): If a clinical consultation is recorded within a window that overlaps for more than 5 days with the prophylactic bunching period of a previous consultation, then we group together the set of windows overlapping with the said prophylactic bunching period. We say that the hypnozoite activation and/or immediate sporozoite development event giving rise to the relevant consultation may have occurred in any window within this group. If chloroquine is administered in window $i$, then the corresponding bunching period generally spans windows $(i + 2)$ and $(i + 3)$ (Table A.2) with some adjustments for treatment failure (Appendix A.1.4); that is, given a hypnozoite activation and/or immediate sporozoite development event in window $(i + 2)$, we allow for the potentially delayed manifestation of bloodstream infection in window $(i + 3)$.

- *No clinical consultation* (state $H$): This indicates a window during which no clinical consultation is recorded; does not belong to the bunching group of a recorded consultation; and for which less than 5 days are censored (due to either a lapse of clinical follow-up or prophylactic masking).

For each child $\ell$, we recover a vector $\mathbf{C}^{(\ell)} \in \{M, C, B, H\}^{n_{\text{obs}}}$. To show how the discretised data are represented, Figure A.5 shows the discretised vivax infection states for children aged 7 at enrolment.

##### A.5.2 Seasonality

Hypnozoite latency periods obscure seasonality in the force of inoculation for *P. vivax*. The incidence of falciparum infection is a more direct correlate of mosquito inoculation rates. We estimate relative seasonal fluctuations in the force of inoculation for *P. vivax* from the incidence of symptomatic falciparum infection (computed after screening for treatment failure, and correcting the time at risk for left/right censoring, post-treatment prophylaxis and documented camp absences). We assume that the force of inoculation for *P. vivax* is piecewise constant over $T = 10$ day windows, and encode relative seasonal fluctuations in a seasonality vector $\mathbf{S}$, discretised over $T = 10$ day windows. To estimate $\mathbf{S}$ over the study period, we fit a cubic smoothing spline (24 degrees of freedom, using the R function `stats::smooth.spline` [14]) to the incidence of symptomatic falciparum infection averaged over the SPf66 cohort, calculated in 20 day windows with a moving average across half-windows (Figure A.6). We normalise $\mathbf{S}$ to

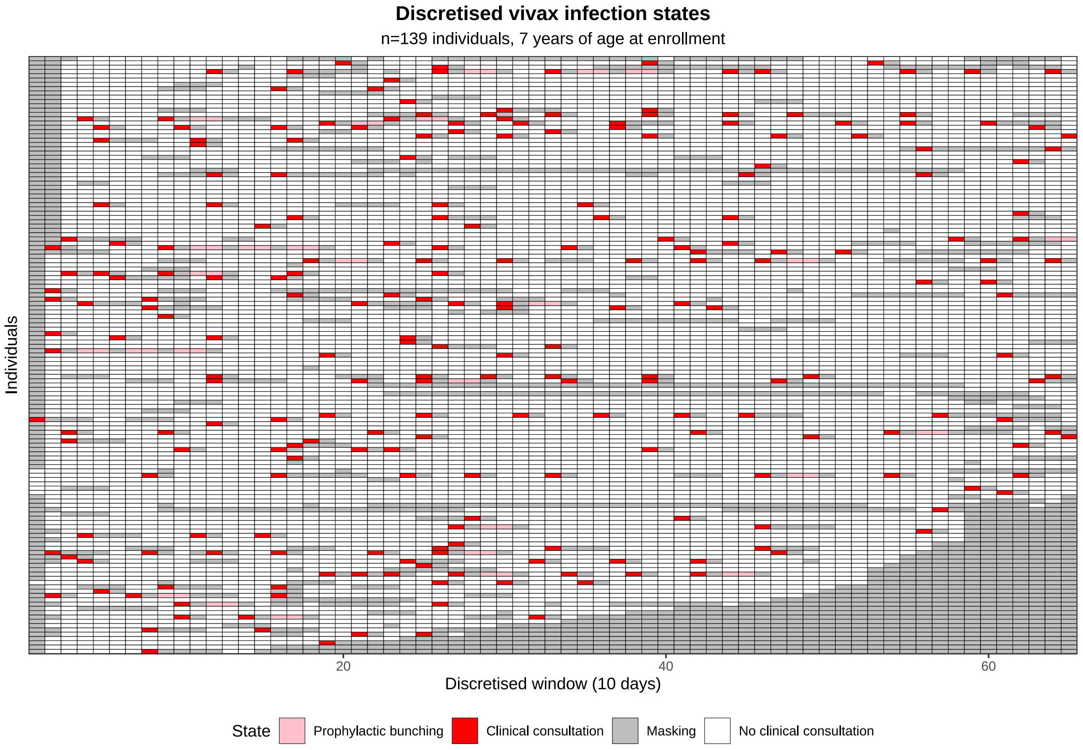

**Figure A.5:** Discretised vivax infection states, adjusted for post-treatment prophylaxis, for children aged 7 at enrolment.

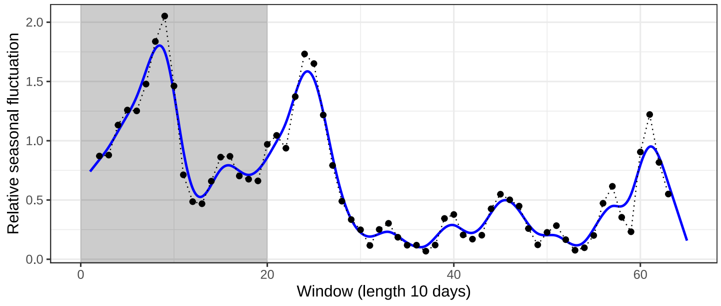

**Figure A.6:** Seasonality estimate. Relative seasonal fluctuations in the incidence of symptomatic falciparum infection (calculated in 20 day windows with a moving average across half windows) are shown in black. The smoothing spline fit is shown in blue.

yield a mean of one in the first 200 days of the study. We do not account for seasonal fluctuations and systematic changes in transmission prior to the study period, whereby we set $S_i = 1$ for all windows $i$ preceding the study.

We then parameterise the force of inoculation for *P. vivax* with a set of scaling factors, that is, the force of inoculation in window $i$ is given by the product of $S_i$ and a scaling factor $\Lambda$. Given the concerted shift towards artemisinin-based combination therapies (ACT) for *P. falciparum* across the camp in mid-1994 — and the consequent reduction in the transmission of *P. falciparum* infection [17] — we accommodate a shift in the relative inoculation rates of *P. falciparum* vs *P. vivax* by adopting the scaling factor $\Lambda_1$ until day 200 (or window 20, this corresponds to April 1994) of the study period, and the scaling factor $\Lambda_2$ thereafter. $\Lambda_1$ can be interpreted as the "historical" force of inoculation preceding the study, and is an important determinant of the initial hypnozoite burden. We calculate the ratio of the respective inoculation rates for vivax and falciparum based on the unnormalised smoothing spline; that is, we interpret the daily incidence rate, calculated over 20 day windows with a moving average over half-windows, as the daily force of inoculation for *P. falciparum*.

### Appendix B

### Theoretical framework

#### B.1 An open network of infinite server queues

##### B.1.1 The within-host framework of Mehra et al. [2]

We begin by recapitulating the short-latency within-host model of Mehra et al. [2], in which each hypnozoite is susceptible to activation directly upon establishment in the host liver [18]. In short, we construct an open network of infinite server queues, with nodes $\{H, A, D, P\}$ (Figure B.1) such that:

- node $H$ corresponds to latent (but activatable) hypnozoites within the liver;
- node $A$ corresponds to hypnozoites that have activated;
- node $D$ corresponds to hypnozoites that have died before activating; and
- node $P$ corresponds to primary infections.

The arrival process for the queueing network — corresponding to the sequence of infective mosquito bites — is assumed to follow a non-homogeneous Poisson process with rate $\lambda(t)$, referred to hereafter as the force of inoculation. Associated with each arrival event is:

- With probability $p_{\text{prim}}$, the initiation of a primary infection, whereby the occupancy of node $P$ increases by one.
- The addition of a batch of hypnozoites in the liver (geometrically-distributed with mean size $\beta$ and state space $\mathbb{Z}_{\geq 0}$), whereby the occupancy of node $H$ increases by $n$ with prob-

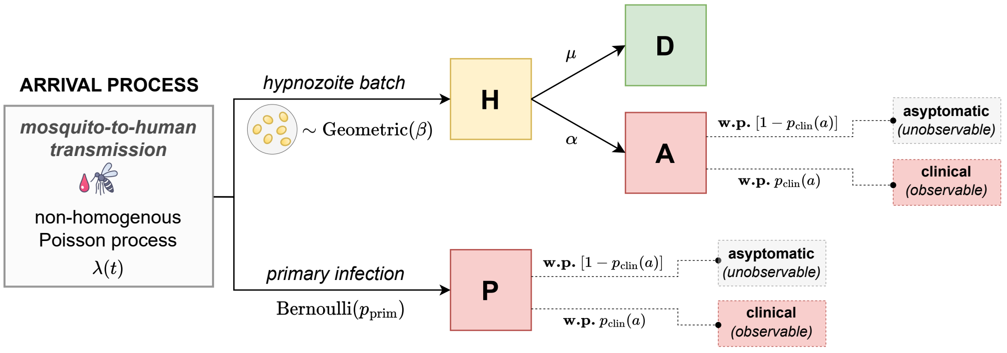

**Figure B.1:** Schematic of the open network of infinite server queues used to model within-host liver- and bloodstream infection. Adapted from Figure 1 of Mehra et al. (2022), *arXiv:2208.10403* and Figure 3 of [2]. **w.p.**: with probability.

ability:

$$p_n = \frac{1}{1+\beta}\Big(\frac{\beta}{1+\beta}\Big)^n.$$

Independent stochastic processes govern the dynamics of each hypnozoite in node $H$. Each hypnozoite remains in node $H$ for an exponentially-distributed period of time, of mean duration $1/(\alpha+\mu)$, where-after it enters either node $A$ with probability $\alpha/(\alpha+\mu)$; or node $D$ with probability $\mu/(\alpha+\mu)$. Here, we additionally impose a simple model of age-dependent anti-disease immunity, whereby each primary infection (that is, arrival into node $P$) and relapse (that is, arrival into node $A$) is independently marked to be clinical with an age-dependent probability $p_{\text{clin}}(a)$, and asymptomatic otherwise.

##### B.1.2 An extension allowing for sporozoite destiny

We modify the arrival process of the queueing network constructed in Mehra et al. [2] by allowing for stochastic sporozoite fating. Each sporozoite is independently assigned one of two fates: with probability $(1-p_{\text{hyp}})$, it undergoes immediate development, whereby it is routed into queue $P$; and with probability $p_{\text{hyp}}$, it forms a hypnozoite whereby it is routed into queue $H$. As such, the probability of an infectious bite giving rise to a primary infection is dependent on the size of the sporozoite inocolum, rather than being held fixed. The immediate development of one or more sporozoites is assumed to give rise to a primary infection. Arrivals into queues $H$ and $P$, however, behave identically to the model detailed above.

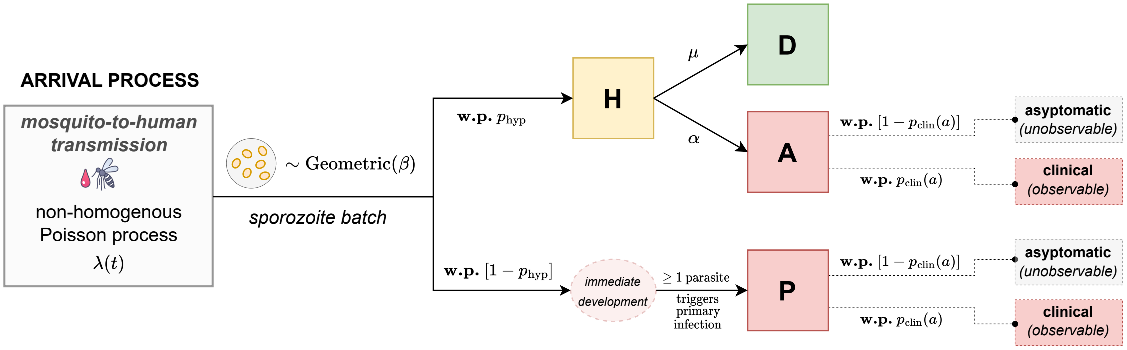

**Figure B.2:** Schematic of the extended within-host model, allowing for stochastic fating of each sporozoite. Adapted from Figure 1 of Mehra et al. (2022), *arXiv:2208.10403*. **w.p.**: with probability.

###### B.1.2.1 A re-parameterised model

Each arrival into the queuing network depicted in Figure B.2 can take one of three routes:

- With probability $(1 - p_{\text{hyp}})$, it enters node $P$ (that is, a sporozoite undergoes immediate development).
- With probability $p_{\text{hyp}}\alpha/(\alpha + \mu)$, it enters node $H$ but is subsequently routed to node $A$ (that is, a sporozoite develops into a hypnozoite that subsequently activates).
- With probability $p_{\text{hyp}}\mu/(\alpha + \mu)$, it enters node $H$ but is subsequently routed to node $D$ (that is, a sporozoite develops into a hypnozoite that dies prior to activation).

Here, we are interested in the temporal distribution of hypnozoite activation events and immediate sporozoite development. This is equivalent to the temporal distribution of arrivals into nodes $A$ and $P$ respectively. The occupancy of node $D$ is completely unobservable using data from bloodstream infections only. By preemptively marking each incoming sporozoite with its eventual fate or destination, we can restrict ourselves to the "successful" subset of the sporozoite inoculum that gives rise to either primary infection or relapse. Denote by $S_{\text{successful}}$ the size of the successful sporozoite inoculum, and $S$ the size of the total sporozoite inoculum, which has PGF

$$\mathbb{E}\left[z^S\right] = \big(1 + \beta(1 - z)\big)^{-1}. \tag{B.1}$$

Using the law of total probability, it is straightforward to show that

$$\mathbb{E}\left[z^{S_{\text{successful}}}\right] = \mathbb{E}\left[\left(\frac{p_{\text{hyp}}\mu}{\alpha+\mu} + \left(1 - \frac{p_{\text{hyp}}\mu}{\alpha+\mu}\right)z\right)^S\right] = \left(1 + \beta\left(1 - \frac{p_{\text{hyp}}\mu}{\alpha+\mu}\right)(1 - z)\right)^{-1},$$

or equivalently,

$$S_{\text{successful}} \sim \text{Geometric}\Big(\frac{1}{\nu+1}\Big)$$

where we denote

$$\nu := \mathbb{E}[S_{\text{successful}}] = \beta\Big(1 - \frac{p_{\text{hyp}}\mu}{\alpha+\mu}\Big).$$

Given a sporozoite is successful, it gives rise to a relapse with probability

$$p_{\text{rel}} = \frac{p_{\text{hyp}}\frac{\alpha}{\alpha+\mu}}{(1-p_{\text{hyp}}) + p_{\text{hyp}}\frac{\alpha}{\alpha+\mu}}.$$

Further, given a hypnozoite eventually activates, we can readily show that the time to activation is exponentially-distributed with rate

$$\eta := \alpha + \mu.$$

Omitting the node $D$ corresponding to (unobservable) hypnozoite death, we can reparameterise the queueing network in Figure B.2 to recover the system depicted in the main text. Interpretation of the parameters $\{\nu, p_{\text{rel}}\}$ is slightly different however.

#### B.2 Derivation of model likelihood: binary clinical infection states in discretised windows

Here, we characterise the temporal patterns of clinical recurrence that emerge under the theoretical framework. For simplicity, rather than treating consultation times as continuous random variables, we discretise observations into uniform windows and assign each window a binarised clinical infection state. Sporozoite inocula are assumed to be geometrically-distributed [18].

In Section B.2.1, we derive a multivariate PGF for the number of immediate sporozoite development and hypnozoite activation events $N_i$ in each window $i$. The assumption of a piecewise-constant force of inoculation yields an analytic, closed-form expression for the multivariate PGF. To account for anti-disease immunity, we impose an age-dependent masking effect that acts independently on each immediate sporozoite development and hypnozoite activation event and governs the absence/presence of clinical symptoms (Section B.2.2). We then formulate a binary infection state $c_i \in \{0, 1\}$, whereby $C_i = 1$ if at least one hypnozoite activation or reinfection event gives rise to clinical symptoms in window $i$, and $C_i = 0$ otherwise. A procedure for recovering the likelihood of a sequence of binary infection states $\mathbf{C}$, drawing on the inclusion-exclusion principle, is delineated in Section B.2.3. Given the use of slowly-eliminated antimalarials (namely, chloroquine and mefloquine) over the course of the SPf66 trial, we also propose a simple observation model to account for extended periods of prohylaxis and the delayed manifestation of subsequent bloodstream infection ('prophylactic bunching') following antimalarial treatment [5] (Section B.2.4). An extension to allow for population heterogeneity in the form of a Gamma-distributed force of inoculation [19] is detailed in Section B.2.5.

##### B.2.1 A multivariate PGF for the temporal distribution of immediate sporozoite development and hypnozoite activation events

Consider a discrete set of times $0 < t_1 < \cdots < t_n$, with $t_0 = 0$. Denote by $N_i$ the number of initiated bloodstream infections — that is, hypnozoite activation events and immediately-developing sporozoite batches, or equivalently, arrivals into nodes $A$ and $P$ — in the interval $(t_{i-1}, t_i]$. Here, we derive the multivariate probability generating function (PGF)

$$\mathbb{E}\left[\prod_{i=1}^{n} z_i^{N_i}\right] = \sum_{\ell_1=0}^{\infty} \cdots \sum_{\ell_n=0}^{\infty} P(N_1 = \ell_1, \dots, N_n = \ell_n) \prod_{i=1}^{n} z_i^{\ell_i},$$

which uniquely characterises the distribution of $\mathbf{N} = (N_1, \dots, N_n)$. To do so, we condition first on the size of an incoming sporozoite batch, and then the sequence of arrival/bite times, following an analogous approach to [2, 20–22] and others. We denote by

$$B(u) = 1 - e^{-\eta u} \tag{B.2}$$

the cdf of the time to activation for each hypnozoite.

Suppose a successful sporozoite is inoculated at time $t_{j-1} < \tau < t_j$. Then the multivariate PGF for the number of hypnozoite activation $H_i$ and immediate sporozoite development $F_i$ events in each interval $(t_{i-1}, t_i]$ can be expressed

$$\begin{aligned}
&\mathbb{E}\left[\prod_{i=1}^{n} x_i^{H_i} y_i^{F_i} \,\middle|\, \text{sporozoite established at time } t_{j-1} < \tau < t_j\right] \\
&= \underbrace{(1 - p_{\text{rel}}) y_j}_{\text{immediate development}} + \underbrace{p_{\text{rel}}[1 - B(t_n - \tau)]}_{\text{hypnozoite activates after } t_n} + \underbrace{p_{\text{rel}} B(t_j - \tau) x_j}_{\text{hypnozoite activates in } (\tau, t_j]} \\
&\quad + \sum_{k=j+1}^{n} \underbrace{p_{\text{rel}} \big[B(t_k - \tau) - B(t_{k-1} - \tau)\big] x_k}_{\text{hypnozoite activates in } (t_{k-1}, t_k]}.
\end{aligned}$$

Under the assumption that each sporozoite is governed by an independent stochastic process, a batch arrival of $M$ sporozoites at time $t_{j-1} < \tau < t_j$ yields the PGF

$$\mathbb{E}\left[\prod_{i=1}^{n} x_i^{H_i} y^{F_i} \,\Big|\, M \text{ sporozites established at time } t_{j-1} < \tau < t_j\right]$$
$$= \Big\{(1-p_{\text{rel}})y_j + p_{\text{rel}}\Big[1 - B(t_n - \tau) + x_j B(t_j - \tau) + \sum_{k=j+1}^{n} x_k\big[B(t_k - \tau) - B(t_{k-1} - \tau)\big]\Big]\Big\}^M.$$

A routine application of the law of total expectation — accommodating a geometrically-distributed sporozoite batch of mean size $\nu$ at time $t_{j-1} < \tau < t_j$ — yields

$$\mathbb{E}\left[\prod_{i=1}^{n} x_i^{H_i} y^{F_i} \,\Big|\, \text{bite at time } t_{j-1} < \tau < t_j\right]$$
$$= \bigg(1 + \nu\Big\{1 - (1-p_{\text{rel}})y_j - p_{\text{rel}}\Big[1 - B(t_n - \tau) + x_j B(t_j - \tau) + \sum_{k=j+1}^{n} x_k\big[B(t_k - \tau) - B(t_{k-1} - \tau)\big]\Big]\Big\}\bigg)^{-1}. \tag{B.3}$$

Noting that $N_i = H_i + \mathbb{1}\{F_i \geq 1\}$ for a single inoculation, we use Equation (B.3) to recover the joint PGF of $\mathbf{N}$ given a single inoculation at time $t_{j-1} < \tau < t_j$:

$$\mathbb{E}\left[\prod_{i=1}^{n} z_i^{N_i} \,\Big|\, \text{bite at time } t_{j-1} < \tau < t_j\right]$$
$$= z_j\bigg(1 + \nu p_{\text{rel}}\Big[B(t_n - \tau) - z_j B(t_j - \tau) - \sum_{k=j+1}^{n} z_k\big[B(t_k - \tau) - B(t_{k-1} - \tau)\big]\Big]\bigg)^{-1}$$
$$+ (1-z_j)\bigg(1 + \nu(1-p_{\text{rel}}) + \nu p_{\text{rel}}\Big[B(t_n - \tau) - z_j B(t_j - \tau) - \sum_{k=j+1}^{n} z_k\big[B(t_k - \tau) - B(t_{k-1} - \tau)\big]\Big]\Big\}\bigg)^{-1}. \tag{B.4}$$

Drawing on the independent increment property of non-homogeneous Poisson processes, we note that the number of bites $S_j$ in each disjoint interval $(t_{j-1}, t_j]$ are independent random variables, with

$$S_j \sim \text{Poisson}\Big(\int_{t_{j-1}}^{t_j} \lambda(\tau) d\tau\Big).$$

Further, conditional on $S_j = n_j$ bites in the interval $(t_{j-1}, t_j]$, the (unordered) bite times $T_1^{(j)}, \dots, T_{n_j}^{(j)}$ are i.i.d. with pdf

$$f_j(\tau) = \frac{\lambda(\tau)}{\int_{t_{j-1}}^{t_j} \lambda(\tau) d\tau} \cdot \mathbb{1}_{\{t_{j-1} < \tau < t_j\}}.$$

Through a standard conditioning argument, in which we consider the number of bites $S_j$ in each disjoint interval $(t_{j-1}, t_j]$ followed by the conditional sequence of bite times $T_1^{(j)}, \dots, T_{n_j}^{(j)}$

— with the critical assumption of mutual independence between hypnozoite activation times — we obtain the multivariate PGF

$$\mathbb{E}\left[\prod_{i=1}^{n} z_i^{N_i}\right] = \exp\left\{ -\sum_{j=1}^{n} \int_{t_{j-1}}^{t_j} \lambda(\tau)\left(1 - \mathbb{E}\left[\prod_{i=1}^{n} z_i^{N_i} \,\middle|\, \text{bite at time } t_{j-1} < \tau < t_j\right]\right) d\tau \right\}. \tag{B.5}$$

Substituting Equation (B.3) into (B.5) yields the multivariate PGF for $\mathbf{N}$:

$$\begin{aligned}\mathbb{E}\left[\prod_{i=1}^{n} z_i^{N_i}\right] = \exp\Bigg\{ \sum_{j=1}^{n} \int_{t_{j-1}}^{t_j} \lambda(\tau)\Bigg[ &-1 + z_j\Big(1 + \nu p_{\text{rel}}\Big[B(t_n - \tau) - z_j B(t_j - \tau) - \sum_{k=j+1}^{n} z_k\big[B(t_k - \tau) - B(t_{k-1} - \tau)\big]\Big]\Big)^{-1} \\ &+ (1 - z_j)\Big(1 + \nu(1 - p_{\text{rel}}) + \nu p_{\text{rel}}\Big[B(t_n - \tau) - z_j B(t_j - \tau) - \sum_{k=j+1}^{n} z_k\big[B(t_k - \tau) - B(t_{k-1} - \tau)\big]\Big]\Big\}\Big)^{-1}\Bigg] d\tau \Bigg\}\end{aligned} \tag{B.6}$$

For the service time distribution $B(u) = 1 - e^{-\eta u}$, the assumption of a piecewise-constant force of inoculation on each interval $(t_{j-1}, t_j]$, that is,

$$\lambda(t) = \lambda_j \text{ for all } t \in (t_{j-1}, t_j]$$

allows us to evaluate Equation (B.6) analytically. For convenience, we restrict ourselves hereafter to uniformly separated intervals $t_j = jT$. In this setting, we obtain

$$\begin{aligned}\mathbb{E}\left[\prod_{i=1}^{n} z_i^{N_i}\right] = \exp\Bigg\{ \sum_{j=1}^{n} \lambda_j \int_{t_{j-1}}^{t_j} \Bigg[ &-1 + z_j\Big(1 + \nu p_{\text{rel}}(1 - z_j) - \nu p_{\text{rel}} h_j(\mathbf{z}, \mathbf{t}) e^{\eta\tau}\Big)^{-1} \\ &+ (1 - z_j)\Big(1 + \nu(1 - p_{\text{rel}} z_j) - \nu p_{\text{rel}} h_j(\mathbf{z}, \mathbf{t}) e^{\eta\tau}\Big)^{-1}\Bigg] d\tau \Bigg\},\end{aligned} \tag{B.7}$$

where we define

$$h_j(\mathbf{z}, \mathbf{t}) = e^{-\eta n T} - z_j e^{-\eta j T} + (1 - e^{-\eta T}) \sum_{k=j+1}^{n} z_k e^{-\eta(k-1)T}.$$

Using standard integral 2.313.1 of [23], we compute

$$\begin{aligned}\mathbb{E}\left[\prod_{i=1}^{n} z_i^{N_i}\right] = \prod_{j=1}^{n} e^{-\lambda_j T} &\left(1 - \frac{\big(1 + \nu p_{\text{rel}}(1 - z_j)\big)\big(1 - e^{-\eta T}\big)}{1 + \nu p_{\text{rel}} - \nu p_{\text{rel}}\big[e^{-\eta(n-j+1)T} + (1 - e^{-\eta T})\sum_{k=j}^{n} z_k e^{-\eta(k-j)T}\big]}\right)^{-\frac{\lambda_j z_j}{\eta(1+\nu p_{\text{rel}}(1-z_j))}} \\ &\left(1 - \frac{\big(1 + \nu(1 - p_{\text{rel}} z_j)\big)\big(1 - e^{-\eta T}\big)}{1 + \nu - \nu p_{\text{rel}}\big[e^{-\eta(n-j+1)T} + (1 - e^{-\eta T})\sum_{k=j}^{n} z_k e^{-\eta(k-j)T}\big]}\right)^{-\frac{\lambda_j (1-z_j)}{\eta(1+\nu(1-p_{\text{rel}} z_j).)}}\end{aligned} \tag{B.8}$$

Now, consider a study period of duration $n_{\text{obs}} T$. For an individual of age $n_{\text{age}} T$ at the onset of the study, we account for a period of hypnozoite accrual $\big(0, n_{\text{age}} \cdot T\big]$ during which initiated bloodstream infections are unobserved/masked; but seek to recover the complete distribution of infection during the study period $\big(n_{\text{age}}T,\, (n_{\text{age}} + n_{\text{obs}})T\big]$. Reindexing

$$V_i := N_{i+n_{\text{age}}} \text{ for } i = \{1, \dots, n_{\text{obs}}\},$$

we recover the multivariate PGF for $\mathbf{V}$ by plugging the vector

$$\mathbf{z} = \big(z_1 = 1, \dots, z_{n_{\text{age}}} = 1, x_1, \dots, x_{n_{\text{obs}}}\big)$$

into Equation (B.8). This yields the expression

$$\begin{aligned}
\mathbb{E}\left[\prod_{i=1}^{n_{\text{obs}}} x_i^{V_i}\right] = \prod_{j=1}^{n_{\text{age}}} e^{-\lambda_j T} \left(1 - \frac{1 - e^{-\eta T}}{1 + \nu p_{\text{rel}} e^{-\eta T(n_{\text{age}}-j)} \Big[e^{-\eta T} - e^{-\eta T(n_{\text{obs}}+1)} - (1 - e^{-\eta T}) \sum_{k=1}^{n_{\text{obs}}} x_k \cdot e^{-\eta T k}\Big]}\right)^{-\frac{\lambda_j}{\eta}} \\
\prod_{j=1}^{n_{\text{obs}}} e^{-\lambda_{j+n_{\text{age}}} T} \left(1 - \frac{\big(1 + \nu p_{\text{rel}}(1 - x_j)\big)\big(1 - e^{-\eta T}\big)}{1 + \nu p_{\text{rel}} - \nu p_{\text{rel}} \big[e^{-\eta(n_{\text{obs}}-j+1)T} + (1 - e^{-\eta T}) \sum_{k=j}^{n_{\text{obs}}} x_k e^{-\eta(k-j)T}\big]}\right)^{-\frac{\lambda_{j+n_{\text{age}}} x_j}{\eta(1+\nu p_{\text{rel}}(1-x_j))}} \\
\left(1 - \frac{\big(1 + \nu(1 - p_{\text{rel}} x_j)\big)\big(1 - e^{-\eta T}\big)}{1 + \nu - \nu p_{\text{rel}} \big[e^{-\eta(n_{\text{obs}}-j+1)T} + (1 - e^{-\eta T}) \sum_{k=j}^{n_{\text{obs}}} x_k e^{-\eta(k-j)T}\big]}\right)^{-\frac{\lambda_{j+n_{\text{age}}}(1-x_j)}{\eta(1+\nu(1-p_{\text{rel}} x_j))}}.
\end{aligned} \tag{B.9}$$

Suppose the hypnozoite reservoir has reached stationarity under a constant force of inoculation $\lambda$ prior to the study period period. Then from [2], the hypnozoite burden $H$ at time zero has PGF

$$\mathbb{E}\left[z^H\right] = \big(1 + \nu p_{\text{rel}}(1 - z)\big)^{-\frac{\lambda}{\eta}}.$$

Using the law of total expectation, the number of hypnozoite activation events $W_i$ in window $i$ of the study period attributable to hypnozoites established prior to the study period takes the form

$$\mathbb{E}\left[\prod_{i=1}^{n} z_i^{W_i}\right] = \left(1 + \nu p_{\text{rel}} \left[1 - e^{-n_{\text{obs}} T} - \sum_{j=1}^{n} e^{-\eta(i-1)T}(1 - e^{-\eta T}) z_j\right]\right)^{-\frac{\lambda}{\eta}}.$$

Therefore, in the limit $n_{\text{age}} \to \infty$ with $\lambda_j = \lambda$ for all $j \leq n_{\text{age}}$, we obtain the expression

$$\begin{aligned}
\mathbb{E}\left[\prod_{i=1}^{n_{\text{obs}}} x_i^{V_i}\right] = \left(1 + \nu p_{\text{rel}} \left[1 - e^{-\eta n_{\text{obs}} T} - \sum_{i=1}^{n} e^{-\eta(i-1)T}(1 - e^{-\eta T}) x_j\right]\right)^{-\frac{\lambda}{\eta}} \\
\prod_{j=1}^{n_{\text{obs}}} e^{-\lambda_{j+n_{\text{age}}} T} \left(1 - \frac{\big(1 + \nu p_{\text{rel}}(1 - x_j)\big)\big(1 - e^{-\eta T}\big)}{1 + \nu p_{\text{rel}} - \nu p_{\text{rel}} \big[e^{-\eta(n_{\text{obs}}-j+1)T} + (1 - e^{-\eta T}) \sum_{k=j}^{n_{\text{obs}}} x_k e^{-\eta(k-j)T}\big]}\right)^{-\frac{\lambda_{j+n_{\text{age}}} x_j}{\eta(1+\nu p_{\text{rel}}(1-x_j))}} \\
\left(1 - \frac{\big(1 + \nu(1 - p_{\text{rel}} x_j)\big)\big(1 - e^{-\eta T}\big)}{1 + \nu - \nu p_{\text{rel}} \big[e^{-\eta(n_{\text{obs}}-j+1)T} + (1 - e^{-\eta T}) \sum_{k=j}^{n_{\text{obs}}} x_k e^{-\eta(k-j)T}\big]}\right)^{-\frac{\lambda_{j+n_{\text{age}}}(1-x_j)}{\eta(1+\nu(1-p_{\text{rel}} x_j))}}.
\end{aligned} \tag{B.10}$$

##### B.2.2 Clinical infection states

Suppose an individual is of age $a$ at the onset of the study period. To account for anti-disease immunity, we assume that *each* initiated bloodstream infection (that is, a hypnozoite or reinfection event) during the study period gives rise to clinical symptoms with probability $p_{\text{clin}}(a)$. Here, we treat age $a$ as a proxy for prior exposure. For notational convenience, we drop the argument $a$ hereafter.

Denote by $U_i$ the number of clinical bloodstream infections initiated in each interval $(t_{i+n_{\text{age}}-1}, t_{i+n_{\text{age}}}]$ of the study period. It follows that

$$U_i | V_i \sim \text{Binomial}\big(V_i, p_{\text{clin}}\big).$$

A routine application of the law of total expectation allows us to recover the PGF for $U_i$ from that of $V_i$:

$$\mathbb{E}\left[\prod_{i=1}^{n_{\text{obs}}} x_i^{U_i}\right] = \mathbb{E}\left[\prod_{i=1}^{n_{\text{obs}}} \Big(1 - p_{\text{clin}} + p_{\text{clin}} x_i\Big)^{V_i}\right]. \tag{B.11}$$

From Equation (B.9), it follows that

$$\begin{aligned}\mathbb{E}\left[\prod_{i=1}^{n_{\text{obs}}} x_i^{U_i}\right] = &\prod_{j=1}^{n_{\text{age}}} e^{-\lambda_j T}\left(1 - \frac{1 - e^{-\eta T}}{1 + \nu p_{\text{rel}} p_{\text{clin}} e^{-\eta T(n_{\text{age}}-j)}\left[e^{-\eta T} - e^{-\eta T(n_{\text{obs}}+1)} - (1 - e^{-\eta T})\sum_{k=1}^{n_{\text{obs}}} x_k \cdot e^{-\eta T k}\right]}\right)^{-\frac{\lambda_j}{\eta}} \\ &\prod_{j=1}^{n_{\text{obs}}} e^{-\lambda_{j+n_{\text{age}}} T}\left(1 - \frac{\big(1 + \nu p_{\text{rel}} p_{\text{clin}}(1 - x_j)\big)\big(1 - e^{-\eta T}\big)}{1 + \nu p_{\text{rel}} p_{\text{clin}} - \nu p_{\text{rel}} p_{\text{clin}}\left[e^{-\eta(n_{\text{obs}}-j+1)T} + (1 - e^{-\eta T})\sum_{k=j}^{n_{\text{obs}}} x_k e^{-\eta(k-j)T}\right]}\right)^{-\frac{\lambda_{j+n_{\text{age}}}(1 - p_{\text{clin}} + p_{\text{clin}} x_j)}{\eta(1 + \nu p_{\text{rel}} p_{\text{clin}}(1 - x_j))}} \\ &\left(1 - \frac{\big(1 + \nu(1 - p_{\text{rel}}(1 - p_{\text{clin}}) - p_{\text{rel}} p_{\text{clin}} x_j)\big)\big(1 - e^{-\eta T}\big)}{1 + \nu\big(1 - p_{\text{rel}}(1 - p_{\text{clin}})\big) - \nu p_{\text{rel}} p_{\text{clin}}\left[e^{-\eta(n_{\text{obs}}-j+1)T} + (1 - e^{-\eta T})\sum_{k=j}^{n_{\text{obs}}} x_k e^{-\eta(k-j)T}\right]}\right)^{-\frac{\lambda_{j+n_{\text{age}}} p_{\text{clin}}(1 - x_j)}{\eta(1 + \nu(1 - p_{\text{rel}}(1 - p_{\text{clin}}) - p_{\text{rel}} p_{\text{clin}} x_j))}}.\end{aligned} \tag{B.12}$$

##### B.2.3 Binarised clinical infection states

For the purposes of inference, we shift our attention to the binary variable

$$C_i = \mathbb{1}\{U_i > 0\}$$

describing the absence/presence of clinical symptoms in each interval $(t_{i+n_{\text{age}}-1}, t_{i+n_{\text{age}}}]$ of the study period. Under our heavily-simplified model of anti-disease immunity, $C_i = 1$ with probability $1 - (1 - p_{\text{clin}})^n$ given $V_i = n$ hypnozoite activation events or reinfection events with at at least one immediately-developing sporozoite occur in window $i$.

The marginal likelihood of observing no clinical symptoms in window $w$ can be computed by plugging $x_w = 0$, $x_k = 1$ for all $k \neq w$ into Equation (B.8) to yield

$$\begin{aligned}P(C_w = 0) = &\prod_{j=1}^{n_{\text{age}}+w-1} e^{-\lambda_j T}\left(1 - \frac{1 - e^{-\eta T}}{1 + \nu p_{\text{rel}} p_{\text{clin}}(1 - e^{\eta T}) e^{-\eta T(n_{\text{age}}+w-j)}}\right)^{\frac{-\lambda_j}{\eta}} \\ &e^{-\lambda_{w+n_{\text{age}}} T\left[1 - \frac{1 - p_{\text{clin}}}{1 + \nu p_{\text{rel}} p_{\text{clin}}} - \frac{p_{\text{clin}}}{1 + \nu(1 - p_{\text{rel}}(1 - p_{\text{clin}}))}\right]}\Big(1 + \nu p_{\text{rel}} p_{\text{clin}}(1 - e^{-\eta T})\Big)^{\frac{\lambda_{w+n_{\text{age}}}(1 - p_{\text{clin}})}{\eta(1 + \nu p_{\text{rel}} p_{\text{clin}})}}\end{aligned}$$

$$\left(\frac{1+\nu(1-p_{\text{rel}})}{1+\nu(1-p_{\text{rel}})+\nu p_{\text{rel}}p_{\text{clin}}(1-e^{\eta T})}\right)^{\frac{-\lambda_{w+n_{\text{age}}}p_{\text{clin}}}{\eta(1+\nu(1-p_{\text{rel}}(1-p_{\text{clin}}))}}. \tag{B.13}$$

In the limit $n_{\text{age}} \to \infty$ with a constant force of infection $\lambda_j = \lambda$, from Equation (B.10), we obtain the likelihood of no clinical recurrence in a follow-up period of length $T$:

$$\begin{aligned}P(C_1 = 0) =& e^{-\lambda T\left[1-\frac{1-p_{\text{clin}}}{1+\nu p_{\text{rel}}p_{\text{clin}}}-\frac{p_{\text{clin}}}{1+\nu(1-p_{\text{rel}}(1-p_{\text{clin}}))}\right]}\left(1+\nu p_{\text{rel}}p_{\text{clin}}(1-e^{-\eta T})\right)^{-\frac{\lambda}{\eta}\left(1-\frac{1-p_{\text{clin}}}{1+\nu p_{\text{rel}}p_{\text{clin}}}\right)} \\ &\left(\frac{1+\nu(1-p_{\text{rel}})}{1+\nu(1-p_{\text{rel}})+\nu p_{\text{rel}}p_{\text{clin}}(1-e^{\eta T})}\right)^{-\frac{\lambda p_{\text{clin}}}{\eta(1+\nu(1-p_{\text{rel}}(1-p_{\text{clin}})))}}.\end{aligned} \tag{B.14}$$

We can also recover the joint likelihood of the $\mathbf{C}$ analytically from the joint PGF of $\mathbf{U}$, given by Equation (B.8). For notational convenience, given a set of parameters $\{\lambda, \eta, \nu, p_{\text{clin}}\}$, denote the multivariate PGF for $\mathbf{U}$ by

$$f(x_1, \ldots, x_{n_{\text{obs}}}) = \mathbb{E}\left[\prod_{i=1}^{n} x_i^{U_i} \,\middle|\, \lambda, \eta, \nu, p_{\text{clin}}\right].$$

As a base case, we observe that the conditional PGF of $\mathbf{U}$ given $U_{n_{\text{obs}}} = C_{n_{\text{obs}}} = 0$ can be written

$$\begin{aligned}&\mathbb{E}\left[\prod_{i=1}^{n_{\text{obs}}-1} x_i^{U_i} \,\middle|\, C_{n_{\text{obs}}} = 0\right] \cdot P(C_{n_{\text{obs}}} = 0) \\ &\quad= \sum_{\ell_1=0}^{\infty} \cdots \sum_{\ell_{n_{\text{obs}}-1}=0}^{\infty} P(U_1 = \ell_1, \ldots, U_{n_{\text{obs}}-1} = \ell_{n_{\text{obs}}-1}, U_{n_{\text{obs}}} = 0) \prod_{i=1}^{n_{\text{obs}}-1} x_i^{\ell_i} \\ &\quad= f(x_1, \ldots, x_{(n_{\text{obs}}-1)}, 0),\end{aligned}$$

while, given $C_{n_{\text{obs}}} = 1$, or equivalently, $U_{n_{\text{obs}}} > 0$,

$$\begin{aligned}&\mathbb{E}\left[\prod_{i=1}^{n_{\text{obs}}-1} x_i^{U_i} \,\middle|\, C_{n_{\text{obs}}} = 1\right] \cdot P(C_{n_{\text{obs}}} = 1) \\ &\quad= \sum_{\ell_1=0}^{\infty} \cdots \sum_{\ell_{n_{\text{obs}}-1}=0}^{\infty} \sum_{\ell_{n_{\text{obs}}}=1}^{\infty} P(U_1 = \ell_1, \ldots, U_{n_{\text{obs}}-1} = \ell_{n_{\text{obs}}-1}, U_{n_{\text{obs}}} = \ell_{n_{\text{obs}}}) \prod_{i=1}^{n_{\text{obs}}-1} x_i^{\ell_i} \\ &\quad= f(x_1, \ldots, x_{(n_{\text{obs}}-1)}, 1) - f(x_1, \ldots, x_{(n_{\text{obs}}-1)}, 0).\end{aligned}$$

If the observation $C_{n_{\text{obs}}}$ is missing, then we marginalise the multivariate PGF over $U_{n_{\text{obs}}}$ to obtain

$$\mathbb{E}\left[\prod_{i=1}^{n_{\text{obs}}-1} x_i^{U_i}\right] = \sum_{\ell_1=0}^{\infty} \cdots \sum_{\ell_{n_{\text{obs}}-1}=0}^{\infty} P(U_1 = \ell_1, \ldots, U_{n_{\text{obs}}-1} = \ell_{n_{\text{obs}}-1}) \prod_{i=1}^{n_{\text{obs}}-1} x_i^{\ell_i}$$

$$= f(x_1, \dots, x_{(n_{\text{obs}}-1)}, 1).$$

By the inclusion-exclusion principle, it thus follows that

$$P\big(\mathbf{C} = \mathbf{c}\big) = \sum_{\mathbf{y}_+ \in S_+(\mathbf{c})} f(\mathbf{y}_+) - \sum_{\mathbf{y}_- \in S_-(\mathbf{c})} f(\mathbf{y}_-) \tag{B.15}$$

where

$$j(\mathbf{c}) := \{i \in \{1, \dots, n_{\text{obs}}\} : c_i \text{ non-missing}\} \tag{B.16}$$

$$S_+(\mathbf{c}) := \Big\{\mathbf{y} \in \{0,1\}^{n_{\text{obs}}} : y_i = 1 \,\forall\, i \in j(\mathbf{c})^{\complement},\ y_i \leq c_i \,\forall\, i \in j(\mathbf{c}),\ \sum_{i \in j(\mathbf{c})} (c_i - y_i) \text{ even}\Big\} \tag{B.17}$$

$$S_-(\mathbf{c}) := \Big\{\mathbf{y} \in \{0,1\}^{n_{\text{obs}}} : y_i = 1 \,\forall\, i \in j(\mathbf{c})^{\complement},\ y_i \leq c_i \,\forall\, i \in j(\mathbf{c}),\ \sum_{i \in j(\mathbf{c})} (c_i - y_i) \text{ odd}\Big\}. \tag{B.18}$$

The time complexity of evaluating the likelihood $P(\mathbf{C} = \mathbf{c})$ scales exponentially with the number of windows $|\mathbf{c}|$ for which clinical symptoms are observed: the joint PGF $f$ is called $2^{|\mathbf{c}|}$ times to compute the likelihood of $P(\mathbf{C} = \mathbf{c})$. As such, this approach is computationally viable only for comparatively infrequent infection data, as seen in the SPf66 cohort (with at most 13 recorded infections per individual over the course of the study period, after screening treatment failures).

While we can theoretically disentangle (a)symptomatic infection under this framework, exponential time complexity constrains inference on the basis of (a)symptomatic infection: to compute the likelihood of an infection sequence with $n_c$ clinical windows; $n_a$ confirmed asymptomatic windows, and $n_u$ windows with no clinical infection but potentially unobserved asymptomatic infection, the joint PGF $f$ would need to be called $2^{n_c} \cdot 3^{n_u}$ times. Given the temporal sparsity of active detection and the low incidence of clinical infection in the SPf66 cohort, the majority of discretised windows yield no clinical, but potentially unobserved asymptomatic infection. As such, accounting for asymptomatic infection is computationally viable only for coarse windows $T$.

##### B.2.4 Accounting for prophylactic protection and "bunching"

We adopt a simple model of prophylactic protection (spanning $I_{\text{mask}}$ windows) and bunching (spanning $I_{\text{bunch}}$ windows) following each treated recurrence. Specifically, given a clinical recurrence is treated in window $i$, we assume that:

- No immediately-developing sporozoites or hypnozoite activation events in windows $(i + 1), \dots, (i + I_{\text{mask}})$ are able to successfully-establish bloodstream infection, whereby the observations $C_{i+1}, \dots, C_{i+I_{\text{mask}}}$ are masked/set to be missing.

- Any immediately-developing sporozoites or hypnozoite activation events in windows $(i+1+I_{\text{mask}}), \dots, (i+I_{\text{mask}}+I_{\text{bunch}})$ will bunch together and manifest in window $(i+I_{\text{mask}}+I_{\text{bunch}})$; that is, we define a new binary infection state at window $(i + I_{\text{mask}} + I_{\text{bunch}})$

$$B_{i+I_{\text{mask}}+I_{\text{bunch}}} := \mathbb{1}\left\{ \sum_{j=1}^{I_{\text{bunch}}} U_{i+I_{\text{mask}}+j} > 0 \right\},$$

  while masking the observations $C_{i+1+I_{\text{mask}}}, \dots, C_{i+I_{\text{mask}}+I_{\text{bunch}}-1}$.

Application of the inclusion-exclusion principle to the multivariate PGF (B.8), as in Section B.2.3 allows us to recover the likelihood of a sequence of binarised clinical recurrence states, whilst accounting for periods of prophylactic protection and bunching associated with each bout of antimalarial treatment.

##### B.2.5 Accounting for population heterogeneity in the force of inoculation

Suppose seasonality in the force of inoculation is encoded in the vector $\mathbf{S} = (S_1, \dots, S_n)$, such that $\lambda_j = \lambda S_j$ for some scalar $\lambda$. To allow for population heterogeneity in the force of inoculation, following the approach of [19], we model $\lambda$ to be Gamma-distributed

$$\lambda \sim \Gamma(\kappa, \theta) \implies p(\lambda \,|\, \kappa, \theta) = \frac{1}{\Gamma(\kappa)\theta^{\kappa}} \lambda^{\kappa-1} e^{-\frac{\lambda}{\theta}}$$

with the shape-scale parametrisation.

We note from Equation (B.12) that we can write the joint PGF for $\mathbf{U}$ in the form

$$\mathbb{E}\left[ \prod_{i=1}^{n_{\text{obs}}} x_i^{U_i} \,\middle|\, \lambda, \nu, \eta, p_{\text{clin}} \right] = h(\mathbf{x}, \mathbf{S}, \nu, \eta, p_{\text{clin}})^{\lambda} \tag{B.19}$$

where

$$\begin{aligned} h(\mathbf{x}, \mathbf{S}, \nu, \eta, p_{\text{clin}}) = e^{-T \sum_{j=1}^{n} S_j} \prod_{j=1}^{n_{\text{age}}} \left( 1 - \frac{1 - e^{-\eta T}}{1 + \nu p_{\text{rel}} p_{\text{clin}} e^{-\eta T (n_{\text{age}} - j)} \left[ e^{-\eta T} - e^{-\eta T (n_{\text{obs}}+1)} - \sum_{k=1}^{n_{\text{obs}}} x_k \cdot e^{-\eta T k} \right]} \right)^{-\frac{S_j}{\eta}} \\ \prod_{j=1}^{n_{\text{obs}}} \left( 1 - \frac{\left(1 + \nu p_{\text{rel}} p_{\text{clin}} (1 - x_j)\right)\left(1 - e^{-\eta T}\right)}{1 + \nu p_{\text{rel}} p_{\text{clin}} - \nu p_{\text{rel}} p_{\text{clin}} \left[ e^{-\eta (n_{\text{obs}} - j + 1) T} + (1 - e^{-\eta T}) \sum_{k=j}^{n_{\text{obs}}} x_k e^{-\eta (k-j) T} \right]} \right)^{-\frac{S_j (1 - p_{\text{clin}} + p_{\text{clin}} x_j)}{\eta (1 + \nu p_{\text{rel}} p_{\text{clin}} (1 - x_j))}} \\ \left( 1 - \frac{\left(1 + \nu (1 - p_{\text{rel}} (1 - p_{\text{clin}}) - p_{\text{rel}} p_{\text{clin}} x_j)\right)\left(1 - e^{-\eta T}\right)}{1 + \nu \left(1 - p_{\text{rel}} (1 - p_{\text{clin}})\right) - \nu p_{\text{rel}} p_{\text{clin}} \left[ e^{-\eta (n_{\text{obs}} - j + 1) T} + (1 - e^{-\eta T}) \sum_{k=j}^{n_{\text{obs}}} x_k e^{-\eta (k-j) T} \right]} \right)^{-\frac{S_j p_{\text{clin}} (1 - x_j)}{\eta (1 + \nu (1 - p_{\text{rel}} (1 - p_{\text{clin}}) - p_{\text{rel}} p_{\text{clin}} x_j))}}. \end{aligned} \tag{B.20}$$

Using the law of total expectation, we can marginalise Equation (B.19) with respect to $\lambda \sim \Gamma(\kappa, \theta)$

$$\mathbb{E}\left[ \prod_{i=1}^{n} x_i^{U_i} \,\middle|\, \nu, \eta, p_{\text{clin}}, \kappa, \theta \right] = \int_0^{\infty} \mathbb{E}\left[ \prod_{i=1}^{n} x_i^{U_i} \,\middle|\, \lambda, \nu, \eta, p_{\text{clin}} \right] \cdot p(\lambda \,|\, \kappa, \theta) \, d\lambda$$

$$
\begin{aligned}
&= \mathbb{E}_\lambda \left[ h(\mathbf{x}, \mathbf{S}, \nu, \eta, p_{\text{clin}})^\lambda \right] \\
&= \Big(1 - \theta \log h(\mathbf{x}, \mathbf{S}, \nu, \eta, p_{\text{clin}})\Big)^{-\kappa} \qquad \text{(B.21)}
\end{aligned}
$$

where we have recognised the moment generating function (MGF) for the Gamma distribution.

Substituting Equation (B.20) into (B.21) yields the PGF of $\mathbf{U}$, conditional on the parameter set $(\mathbf{S}, \nu, \eta, p_{\text{clin}})$ but marginalised with respect to $\lambda \sim \text{Gamma}(\theta, \kappa)$:

$$
\begin{aligned}
&\mathbb{E}\left[ \prod_{i=1}^{n} x_i^{U_i} \,\Big|\, \mathbf{S}, \nu, \eta, p_{\text{clin}}, \kappa, \theta \right] \\
&= \Bigg(1 + \theta \Bigg\{ \sum_{j=1}^{n} S_j T + \sum_{j=1}^{n_{\text{age}}} \frac{S_j}{\eta(1 + \nu p_{\text{rel}}(1 - z_j))} \log\Bigg(1 - \frac{(1 + \nu p_{\text{rel}}(1 - z_j))(1 - e^{-\eta T})}{1 + \nu p_{\text{rel}} - \nu p_{\text{rel}}\left[e^{-\eta(n-j+1)T} + (1 - e^{-\eta T}) \sum_{k=j}^{n} z_k e^{-\eta(k-j)T}\right]}\Bigg) \\
&\qquad \sum_{j=1}^{n} \frac{S_j(1 - x_j)}{} \log\Bigg(1 - \frac{(1 + \nu p_{\text{clin}} - \nu p_{\text{clin}} x_j)(1 - e^{-\eta T})}{1 + \nu p_{\text{clin}}(1 - e^{-\eta T(n_{\text{obs}} - j + 1)}) - \nu p_{\text{clin}}(1 - e^{-\eta T}) \sum_{k=j}^{n_{\text{obs}}} x_k \cdot e^{-\eta T(k-j)}}\Bigg) \Bigg\} \Bigg)^{-\kappa}. \qquad \text{(B.22)}
\end{aligned}
$$

Application of the inclusion-exclusion principle to the PGF (B.22), as detailed in Section B.2.3, allows us to recover the likelihood binary infection state $C_i = \mathbb{1}\{U_i > 0\}$ for a given set of parameters $\{\mathbf{S}, \nu, \eta, p_{\text{clin}}\}$ whilst marginalising over $\lambda \sim \Gamma(\kappa, \theta)$; the model of propyhlactic protection and bunching proposed in Section B.2.4 also applies.

### Appendix C

### Calibration to the SPf66 vaccine trial

#### C.1 Parameter estimation

##### C.1.1 Metropolis-Hastings algorithm

For the cohort of $n_{\text{cohort}} = 1344$ children, indexed $\ell = 1, \ldots, n_{\text{cohort}}$, we compute the likelihood

$$L\big(\mathbf{C}^{(1)}, \ldots, \mathbf{C}^{(n_{\text{cohort}})} \,\big|\, \Lambda_1, \Lambda_2, \nu, \eta, \rho, \gamma\big) = \prod_{\ell=1}^{n_{\text{cohort}}} L\big(\mathbf{C}^{(\ell)} \,\big|\, \Lambda_1, \Lambda_2, \nu, \eta, \rho, \gamma\big)$$

using the analytic expressions derived in Appendix B.2, where known values/covariates (specifically, the age $n_{\text{age}}^{(\ell)}$ of each child $\ell$ in units of $T$ day windows; the seasonality vector $\mathbf{S}$ estimated from the incidence of clinical falciparum episodes; and the ratio $p_{\text{rel}} = 0.4$ of sporozoites that form hypnozoites, informed by *in vivo* and *in vitro* estimates for the Chesson strain of *P. vivax*) have been dropped for notational convenience.

We take flat improper priors on $(0, \infty)$ for the parameters $\Lambda_1$, $\Lambda_2$, $\nu$ and $\eta$, but informative priors for the parameters governing the age-dependent anti-disease masking curve

$$\text{logit}(\rho) \sim \mathcal{N}(0, 0.7^2) \qquad \log(\gamma) \sim \mathcal{N}(0, 0.6^2).$$

To generate a candidate parameter set $(\Lambda_1^*, \Lambda_2^*, \nu^*, \eta^*, \rho^*, \gamma^*)$ given $(\Lambda_1', \Lambda_2', \nu', \eta', \rho', \gamma')$, we adopt the symmetric proposal distribution

$$\begin{aligned}
\Lambda_1^* &\sim \mathcal{N}^R(\Lambda_1',\, (0.02/365)^2) \\
\Lambda_2^* &\sim \mathcal{N}^R(\Lambda_2',\, (0.02/365)^2) \\
\nu^* &\sim \mathcal{N}^R(\nu',\, 0.2^2) \\
\eta^* &\sim \mathcal{N}^R(\eta',\, 1/2000^2)
\end{aligned}$$

$$\text{logit}(\rho^*) \sim \mathcal{N}\big(\text{logit}(\rho'),\, 0.05^2\big)$$
$$\log(\gamma^*) \sim \mathcal{N}\big(\log(\gamma'),\, 0.05^2\big)$$

where $\mathcal{N}^R(\mu, \sigma^2)$ denotes the rectified normal distribution, equivalent to the normal distribution $\mathcal{N}(\mu, \sigma^2)$ with all negative values mapped to zero.

Initial values $(\Lambda_1', \Lambda_2', \nu', \eta', \rho', \gamma')$ are sampled independently for each parameter from

$$\begin{aligned} \Lambda_1', \Lambda_2' &\sim U[0.1/365,\, 1/365] \\ \nu' &\sim U[0.5,\, 8] \\ \eta' &\sim U[1/500,\, 1/50] \\ \text{logit}(\rho') &\sim \mathcal{N}\big(0,\, 0.7^2\big) \\ \log(\gamma') &\sim \mathcal{N}\big(0,\, 0.6^2\big). \end{aligned}$$

We aggregate results over 4 chains, spanning 100,000 iterations each, and discard the initial 20,000 iterations for each chain as the burn-in period. To assess convergence, we report Gelman-Rubin diagnostic, calculated using Equation (1.1) of [24] after discarding the burn-in period.

##### C.1.2 Trace plots

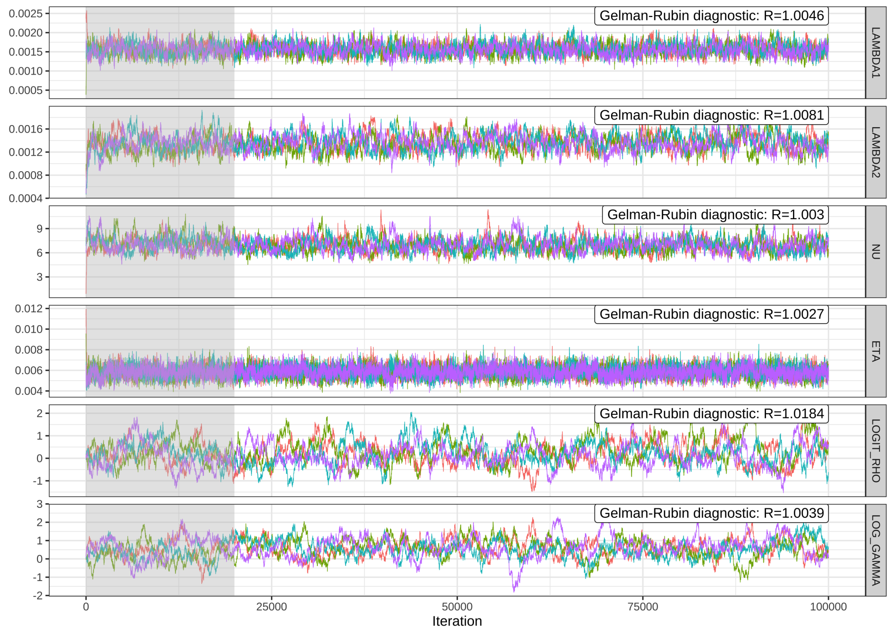

**Figure C.1:** Trace plots for the Metropolis-Hastings fitting regime. The burn-in period is shaded in grey. Averaging over seasonal fluctuations, $\Lambda_1$ represents the mean force of inoculation up to day 200 of the study period, while $\Lambda_2$ represents the mean force of inoculation thereafter.

##### C.1.3 Pairwise posterior distributions

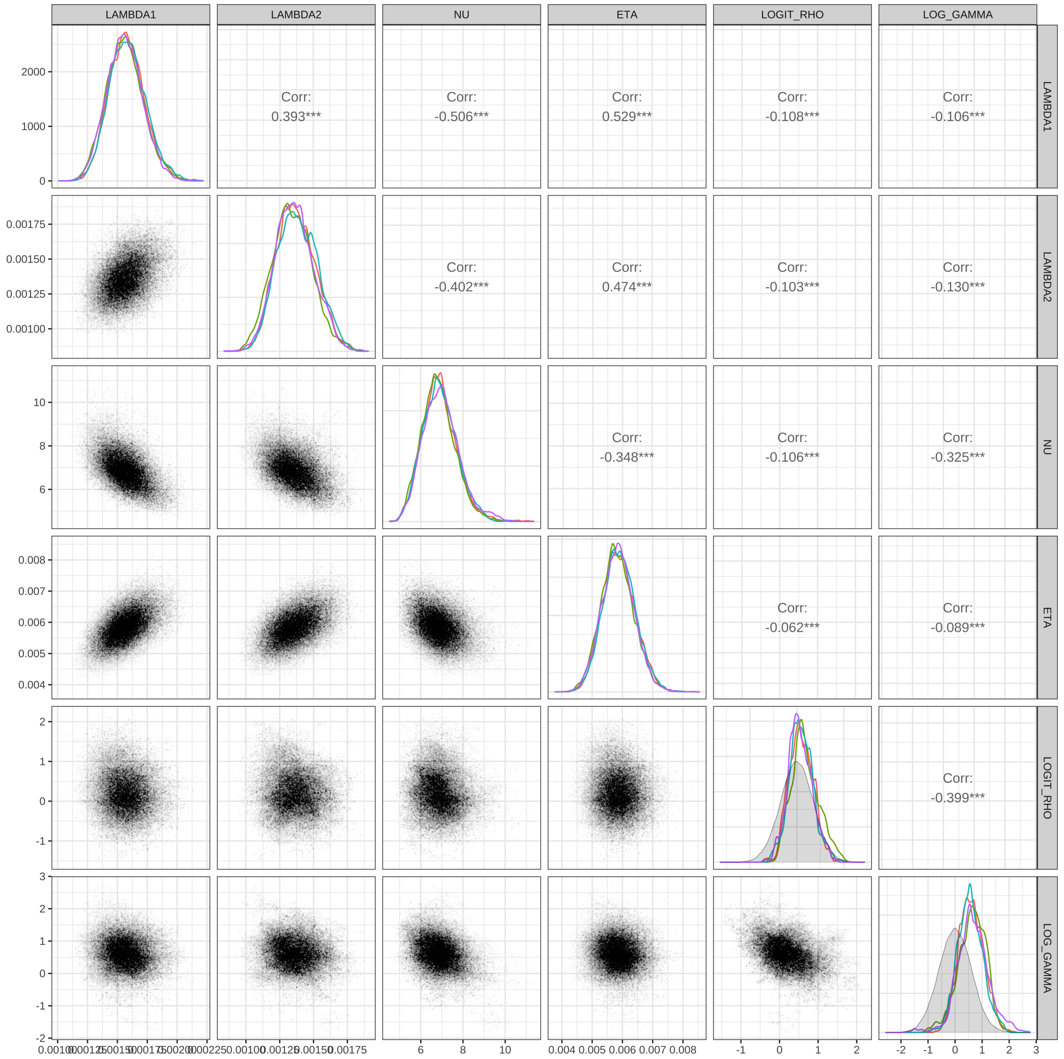

**Figure C.2:** Pairwise joint posterior distributions for each parameter pair (aggregated across chains after discarding the burn-in period). Marginal posteriors for each parameter, stratified by chain, are shown across the diagonal; prior distributions for $\{\text{logit}(\rho), \log(\gamma)\}$ are shown in grey (the other parameters have flat priors). Averaging over seasonal fluctuations, $\Lambda_1$ represents the mean force of inoculation up to day 200 of the study period, while $\Lambda_2$ represents the mean force of inoculation thereafter.

##### C.1.4 Summary of posterior estimates

A decline in malaria transmission was apparent in the second half of the study. Early detection and prompt effective antimalarial treatment throughout the camp was likely a contributing factor. Falciparum malaria declined more than vivax malaria. The ratio of the vivax to falciparum inoculation rates was estimated to rise from 0.61 (95% CrI 0.50 to 0.73) to 1.10 (95% CrI 0.89 to 1.34) after April 1994 following the widespread adoption of mefloquine-artesunate treatment for falciparum malaria throughout the camp.

| Quantity | Interpretation | Median [95% CrI] |
|---|---|---|
| $1/\eta$ | Average duration of hypnozoite carriage | 171 [144, 206] days |
| $\log(2)/\eta$ | Half-life of a hypnozoite in the liver | 118 [100, 143] days |
| $1 - e^{-14\eta}$ | Prob of hypnozoite activation in a 2 week window | 0.08 [0.07, 0.09] |
| $1 - e^{-28\eta}$ | Prob of hypnozoite activation in a 4 week window | 0.15 [0.13, 0.18] |
| $1 - e^{-84\eta}$ | Prob of hypnozoite activation in a 12 week window | 0.39 [0.33, 0.44] |
| $1 - e^{-168\eta}$ | Prob of hypnozoite activation in a 24 week window | 0.63 [0.56, 0.69] |
| $\nu$ | Average sporozoite batch size | 6.9 [5.4, 8.7] |
| $\nu p_{\text{rel}}$ | Average hypnozoite batch size | 2.7 [2.2, 3.5] |
| $1 - 1/(1 + \nu(1 - p_{\text{rel}}))$ | Probability of primary infection per bite | 0.80 [0.77, 0.84] |
| $\nu p_{\text{rel}} + p_{\text{rel}}/(1 - p_{\text{rel}})$ | Ratio of expected relapse vs primary per bite | 3.4 [2.8, 4.2] |
| $\Lambda_1$ | Average force of inoculation before day 200 | 0.57 [0.47, 0.69] year$^{-1}$ |
| $\Lambda_2$ | Average force of inoculation after day 200 | 0.50 [0.40, 0.60] year$^{-1}$ |
| $\Lambda_1(1 - 1/(1 + \nu(1 - p_{\text{rel}})))$ | Average force of primary infection before day 200 | 0.46 [0.38, 0.54] year$^{-1}$ |
| $\Lambda_2(1 - 1/(1 + \nu(1 - p_{\text{rel}})))$ | Average force of primary infection after day 200 | 0.40 [0.33, 0.48] year$^{-1}$ |

**Table C.1:** Summary of posterior median estimates for quantities of epidemiological interest. Posterior median estimates and 95% credible intervals are provided for each quantity.

#### C.2 Posterior predictive checks

##### C.2.1 Simulating symptomatic vivax episodes

To assess the model fit, we simulate symptomatic vivax episodes under the calibrated model; falciparum episodes are not simulated. In generating posterior predictive data, we retain the age distribution and clinical follow-up pattern of the SPf66 cohort. For each child $\ell$ in the SPf66 cohort, we record the age at enrolment $n_{\text{age}}^{(\ell)}$ (in units of $T$ day windows) and extract a masking vector $\mathbf{m}^{(\ell)} \in \{0,1\}^{n_{\text{obs}}}$, where $m_i^{(\ell)} = 0$ if at least 50% of window $i$ was masked for child $\ell$ due to left/right-censoring or a documented camp absence, and $m_i^{(\ell)} = 1$ otherwise.

We sample 2000 parameter combinations $(\lambda, \eta, \nu, \rho, \gamma)$ uniformly at random from the posterior (without replacement). For each parameter combination, we simulate clinical recurrences across $n_{\text{obs}} = 65$ windows, each of length $T = 10$, for each child $\ell = 1, \dots, n_{\text{cohort}}$ as follows.

Denote by $\lambda_{\max} = \max \lambda_i S_i$. We simulate the timing of infectious mosquito bites from birth until the end of the study period by thinning a homogeneous Poisson process with rate $\lambda_{\max}$. We first sample

$$M_{\max}^{(\ell)} \sim \text{Poisson}\big(\lambda_{\max}(n_{\text{age}}^{(\ell)} + n_{\text{obs}})T\big).$$

Conditional on $M_{\max}^{(\ell)}$, we sample a sequence of prospective bite times $\tau_1^{(\ell)}, \dots, \tau_{M_{\max}^{(\ell)}}^{(\ell)}$ independently from the uniform distribution

$$\tau_1^{(\ell)}, \dots, \tau_{M_{\max}^{(\ell)}}^{(\ell)} \overset{\text{i.i.d.}}{\sim} \text{Uniform}\big[0, (n_{\text{age}}^{(\ell)} + n_{\text{obs}})T\big].$$

Each prospective bite time $\tau_q^{(\ell)}$ is placed into a window $\lceil \tau_q^{(\ell)}/T \rceil$, and retained with probability $\lambda_{\lceil \tau_q^{(\ell)}/T \rceil} S_{\lceil \tau_q^{(\ell)}/T \rceil} / \lambda_{\max}$ to yield a thinned set of bite times, denoted $T_1^{(\ell)}, \dots, T_{M^\ell}^{(\ell)}$ hereafter.

The sequence of recurrences associated with each bite $j = 1, \dots, M^{(\ell)}$ is then simulated as per Appendix D.1.2.1, but with a geometrically-distributed sporozoite batch size. These data are collated to recover a binary infection state $\mathbf{I}^{(\ell)} \in \{0, 1\}^{n_{\text{obs}}}$, where $I_k^{(\ell)} = 1$ if at least one recurrence was simulated in the interval $[(n_{\text{age}} + k - 1)T, (n_{\text{age}} + k)T)$, and $I_k^{(\ell)} = 0$ otherwise.

From the binarised sequence of infection states $\mathbf{I}^{(\ell)}$ and masking vector $\mathbf{m}^{(\ell)}$, we construct a ternary infection sequence $\mathbf{C}^{(\ell)} \in \{M, H, C\}^{n_{\text{obs}}}$ corrected for post-treatment prophylaxis and anti-disease masking (with $C_k^{(\ell)} = M$ indicating masking due to left/right-censoring, documented camp absences or prophylaxis; $C_k^{(\ell)} = C$ indicating a detected and treated clinical recurrence in window $k$ and $C_k^{(\ell)} = H$ otherwise). We model post-treatment prophylaxis as follows: if antimalarial treatment is administered in window $k_{\text{treat}}$, we mask clinical recurrences in window $k_{\text{treat}} + 1$ (i.e. account for a period of complete prophylactic protection spanning $T$ days), and delay the detection of clinical recurrences in window $k_{\text{treat}} + 2$ to window $k_{\text{treat}} + 3$ (i.e. account for a prophylactic bunching period spanning $2T$ days). For all $k$ such that $m_k^{(\ell)} = 0$, we set $C_k^{(\ell)} = M$. We then iterate across infection windows $k = 1, \dots, n_{\text{obs}}$ and perform the following steps:

- If $C_k^{(\ell)} \in \{H, M\}$ has already been assigned, we leave as is.
- If $C_k^{(\ell)} = C$ has already been assigned or $C_k^{(\ell)}$ has not yet been assigned and $I_k^{(\ell)} = 1$, we simulate a Bernoulli random variable $A_k^{(\ell)}$ with success parameter $p_{\text{clin}}(n_{\text{age}}^{(\ell)})$ (computed using the anti-disease masking parameters $\rho$ and $\gamma$).

– If $A_k^{(\ell)} = 1$, we set $C_k^{(\ell)} = C$ corresponding to a clinical recurrence that has prompted antimalarial treatment. We additionally set $C_{k+1}^{(\ell)} = M$ (due to complete prophylactic protection). If $m_{k+2} = 1$, then we set $C_{k+2}^{(\ell)} = H$; if additionally $I_{k+2} = 1$ and $m_{k+3} = 1$, then we set $C_{k+3}^{(\ell)} = C$ (to adjust for prophylactic bunching).

– If $A_k^{(\ell)} = 0$, we set $C_k^{(\ell)} = H$ corresponding to an undetected asymptomatic recurrence.

- If $C_k^{(\ell)}$ has not yet been assigned and $I_k^{(\ell)} = 0$, we set $C_k^{(\ell)} = H$.

##### C.2.2 Seasonal fluctuations in the incidence of symptomatic vivax malaria

The aggregated incidence in window $k$ is computed as

$$W_k = \frac{\sum_\ell \mathbb{1}_{\{C_k^{(\ell)}=C\}}}{\sum_\ell \mathbb{1}_{\{C_k^{(\ell)}\neq M\}}},$$

and is shown in Figure C.3. We find that the model is unable to recapitulate seasonal fluctuations in the incidence of symptomatic vivax malaria, particularly in the first 7 months of the study.

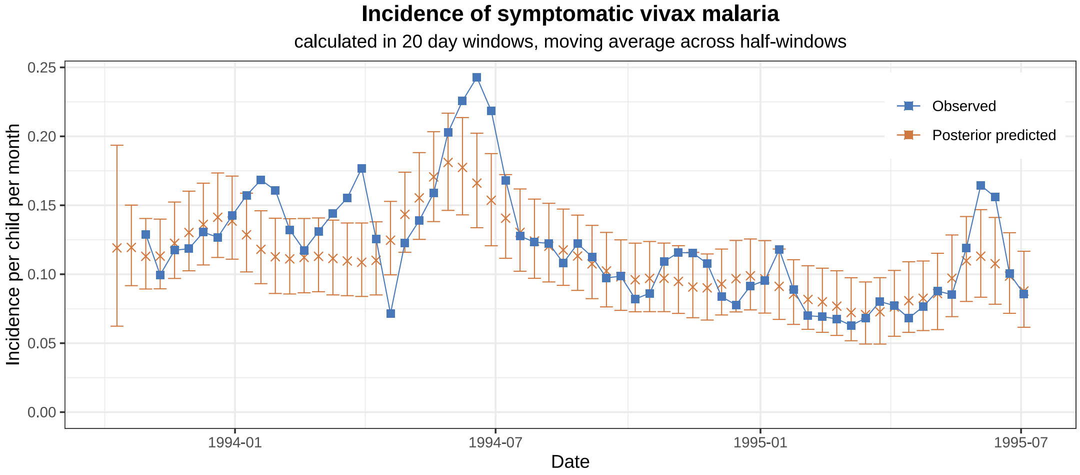

**Figure C.3:** Observed vs posterior predicted incidence by window, aggregated over age groups. Points indicate medians, while error bars show 95% credible intervals for posterior predictive data.

We use the incidence of symptomatic falciparum malaria as a proxy for the force of inoculation. It is likely that observed fluctuations in falciparum incidence were determined by variation in recrudescence rates across the camp more generally. In individuals not enrolled in the SPf66 trial, uncomplicated falciparum malaria in the camp was largely treated with mefloquine monotherapy until early 1994, with high rates of treatment failure [3, 25]. The persistence of gametocytes following mefloquine monotherapy of resistant infections [17] likely had implications for *P. falciparum* transmission across the camp. This unaccounted variation in transmission due to falciparum treatment failure biases our estimates of seasonality in the force of inoculation for vivax malaria.

##### C.2.3 Age structure in the incidence of symptomatic vivax

The simulated incidence rate for age group $a$ is calculated as the quotient

$$R(a) = \frac{\sum_\ell \mathbb{1}_{\{n_{\text{age}}^{(\ell)}=a\}} \sum_{k=1}^{n_{\text{obs}}} \mathbb{1}_{\{C_k^{(\ell)}=C\}}}{\sum_\ell \mathbb{1}_{\{n_{\text{age}}^{(\ell)}=a\}} \sum_{k=1}^{n_{\text{obs}}} \mathbb{1}_{\{C_k^{(\ell)}\neq M\}}}.$$

Under the assumption of a homogeneous force of inoculation across age groups, posterior predictive data yield monotonic age structure in the incidence of symptomatic vivax malaria (see main text). This is at odds with non-monotonicity in the observed age-stratified incidence rate of symptomatic vivax malaria (Appendix A.4).

#### C.3 Rates of vivax recurrence in fixed follow-up windows

##### C.3.1 Posterior predictive distributions

Given a parameter vector $\theta := (\Lambda_1, \Lambda_2, \nu, \eta, \rho, \gamma)$, we compute the likelihood of child $\ell$ experiencing a vivax recurrence in windows $w_0$ to $w_1$ inclusive. In doing so, we condition on the history of vivax recurrence $\mathbf{C}^{(\ell)}_{(w_0)} = (C^{(\ell)}_1, \dots, C^{(\ell)}_{w_0-1})$ prior to window $w_0$. During windows $w_0$ to $w_1$ inclusive, we account for masking due to lapses in clinical follow-up (left- or right-censoring, or a documented absence from the camp), or post-treatment prophylaxis due to the treatment of falciparum monoinfection only. We thus define an additional vector $\mathbf{C}^{(\ell)}_{(w_0,w_1)} = (W^{(\ell)}_1, \dots, W^{(\ell)}_{w_1})$ such that $W^{(\ell)}_i = C^{(\ell)}_i$ for $i < w_0$, while for each window $i \in \{w_0, \dots, w_1\}$, we set $W^{(\ell)}_i = M$ if window $i$ is masked due to censoring or falciparum monoinfection prophylaxis, and $W^{(\ell)}_i = H$ (corresponding to no hypnozoite and/or immediate sporozoite development events) otherwise. Denote by $V^{(\ell)}_{(w_0,w_1)}$ the indicator function that child $\ell$ experiences at least one vivax recurrence in windows $w_0$ to $w_1$ inclusive, conditional on the history of vivax recurrence prior to window $w_0$. We compute the conditional probability

$$L\Big[V^{(\ell)}_{(w_0,w_1)} = 1 \,\Big|\, \theta\Big] = 1 - \frac{L\big(\mathbf{C}_{(w_0,w_1)} \,\big|\, \theta\big)}{L\big(\mathbf{C}_{(w_0)} \,\big|\, \theta\big)}.$$

For a fixed follow-up period spanning $Q$ windows, we identify a set of individuals and baseline time points of interest $S \subset \{1, \dots, n_{\text{cohort}}\} \times \{1, \dots, n_{\text{obs}} - Q\}$, such that for each $(\ell, w) \in S$, child $\ell$ is subject to at least partial clinical follow-up (adjusting for falciparum prophylaxis and censoring) in windows $(w+1)$ to $(w+Q)$ inclusive. Given a parameter vector $\theta$, the proportion of windows $\psi_{\text{vivax}}(S)$ accompanied by a vivax recurrence is modelled to be the scaled sum of a set of independent, but non-identical Bernoulli random variables (i.e. a Poisson binomial distribution), that is,

$$\psi_{\text{vivax}}(S) \,|\, \theta \sim \frac{1}{|S|} \sum_{(\ell,w)\in S} V^{(\ell)}_{(w+1,w+Q)}(\theta)$$

where

$$V^{(\ell)}_{(w+1,w+Q)}(\theta) \overset{\text{independent}}{\sim} \text{Bernoulli}\Big(L\Big[V^{(\ell)}_{(w_0,w_1)} = 1 \,|\, \theta\Big]\Big).$$

As a caveat, we note that the assumption of independence between the random variables $V^{(\ell)}_{(w+1,w+Q)}(\theta)$ is misspecified if multiple baseline windows $w$ are considered for the same child $\ell$; the construction of a joint analysis across multiple follow-up windows, however, is unclear.

To generate posterior predictive distributions for $\psi_{\text{vivax}}(S)$, we sample $n_{\text{posterior}} = 2000$ parameter combinations $\theta$ uniformly at random from the posterior $\pi(\theta)$. We then recover the posterior predictive distribution function

$$P(\psi_{\text{vivax}}(S) \leq x) = \frac{1}{n_{\text{posterior}}} \sum_{i=1}^{n_{\text{posterior}}} P(\psi_{\text{vivax}}(S) \leq x \,|\, \theta_i)$$

using the the Poisson-binomial distribution function implemented in the R package `poisbinom` [26].

##### C.3.2 Confounding due to seasonality

To gauge confounding due to seasonality, we compare the observed vs posterior predicted rates of vivax recurrence across the cohort in fixed follow-up windows (of length 20 to 80 days) at various time points in the SPf66 trial; that is, for $Q \in \{2, \ldots, 8\}$ and $w$ in the range 0 to 55 in increments of 5, we consider $S(w) = \{(\ell, w) : \ell \in \{1, \ldots, n_{\text{cohort}}\}\}$. The Poisson-binomial independence assumption is justified for this analysis, because each child is considered at most once for each baseline time point $w$ and follow-up window $Q$. While seasonality leads to systematic biases, observed rates (closed circles, Figure C.4) generally lie within 99% CrI for posterior predictive distributions (error bars, Figure C.4).

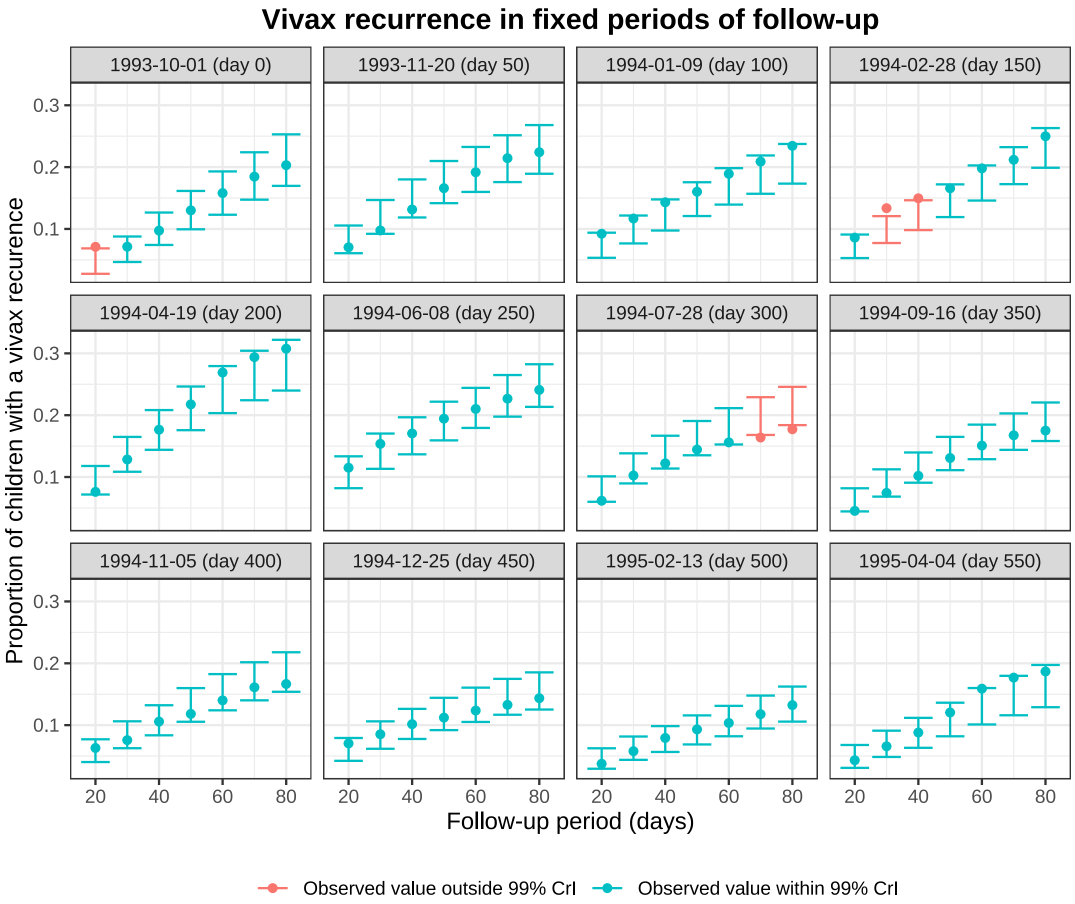

**Figure C.4:** Rates of vivax recurrence in fixed follow-up windows at specified time points in the SPf66 trial. Observed rates are shown with closed circles, while error bars show 99% CrI for posterior predictive distributions.

### Appendix D

### Sensitivity to model misspecification

#### D.1 On the assumption of geometric sporozoite batches

Our theoretical framework is predicated on the assumption of geometric batch sizes (originally proposed by White et al. [18] based on a visual examination of sporozoite inoculum data collected by Beier et al. [27]). This parametric form introduces constraints on the variance-to-mean ratio of sporozoite batch sizes, that may not hold in practice. Here, we characterise the sensitivity of our statistical framework to parametric misspecification of the sporozoite batch size.

##### D.1.1 An extention: negative binomial sporozoite batches

A natural generalisation — which can be motivated under a model of heterogeneity in mosquito infectivity[1] — is a negative binomial distribution governing the sporozoite batch size

$$S \sim \text{NegativeBinomial}\Big(\frac{r}{\nu + r}, r\Big)$$

with PGF

$$\mathbb{E}\left[z^S\right] = \Big(1 + \frac{\nu}{r}(1-z)\Big)^{-r},$$

parametrised by the mean incolum size $\nu$ and the "success" parameter $r$.

The parameter $r$ modulates overdispersion in sporozoite inocula. For $r \leq 1$, we obtain a mode of zero sporozoites with increasing zero inflation as $r \to 0$; for $r > 1$, the mode is instead given by $\lfloor \nu(1 - \frac{1}{r}) \rfloor$, approaching the mean inoculum size $\nu$ in the limit $r \to \infty$ whereby we recover the Poisson distribution with mean $\nu$. In the case $r = 1$, we recover the geometric distribution.

[1] Conditional on a mosquito infectivity coefficient $\zeta$, we claim that sporozoite inocula are Poisson-distributed with mean $\zeta$, but allow $\zeta$ to follow a Gamma distribution with mean $\nu$ and variance $\nu^2/r$.

| Parameter | Interpretation | Values | Units |
|---|---|---|---|
| $\lambda$ | Force of inoculation | $\{0.5/365, 1/365\}$ | $\text{day}^{-1}$ |
| $\eta$ | Hypnozoite activation rate | $\{1/600, 1/400, 1/200, 1/100\}$ | $\text{day}^{-1}$ |
| $\nu$ | Mean sporozoite batch size | 4 | — |
| $r$ | Negative binomial 'size' parameter | $\{0.25, 0.5, 1, 2\}$ | — |
| $p_{\text{rel}}$ | Hypnozoite fating probability | 0.6 | — |

**Table D.1:** Simulated parameter sets

Keeping the mean inoculum size $\nu$ fixed, but reducing overdispersion (i.e. increasing $r$) augments the probability of each infective bite giving rise to a primary infection

$$b_{\text{prim}} := 1 - \Big(1 + \frac{\nu}{r}(1 - p_{\text{rel}})\Big)^{-r}, \tag{D.1}$$

in addition to reducing the expected disparity in the relapse burden conditional on the absence/presence of primary infection.

##### D.1.2 Simulation study details

###### D.1.2.1 A simulation framework assuming negative binomial sporozoite batches

We simulate vivax recurrences under the theoretical framework, allowing for negative binomial sporozoite batches. For clarity, we ignore the effects of anti-disease masking (that is, we set $p_{\text{clin}}(a) = 1$ for all age groups $a$) and assume a constant force of inoculation, setting the seasonality vector $\mathbf{S} = \mathbf{1}$. We assume each observed recurrences is treated with a long-lived antimalarial, adopting a simple model of prophylactic masking/bunching [5].

For each parameter combination $\{\lambda, \eta, \nu, r, p_{\text{rel}}\}$ detailed in Table D.1, we simulate a hypothetical cohort of $n_{\text{cohort}} = 1120$ individuals, with 80 children within each age group $a = 2, 3, \dots, 15$ years. We assume complete follow up over a study period discretised into $n_{\text{obs}} = 65$ windows, each of length $T = 10$ days.

For an individual of age $a$, the number of infective bites $M(a)$ experienced from birth until the end of the study period is sampled from a Poisson distribution

$$M(a) \sim \text{Poisson}\Big(\lambda \cdot (365 \cdot a + n_{\text{obs}} T)\Big).$$

Conditional on $M(a)$, the respective bite times $T_1, \dots, T_{M(a)}$ are sampled independently from the uniform distribution

$$T_1, \dots, T_{M(a)} \overset{\text{i.i.d.}}{\sim} \text{Uniform}\big[0, 365 \cdot a + n_{\text{obs}} T\big].$$

For each bite $j$, we simulate a sporozoite batch

$$S_j \sim \text{NegativeBinomial}\Big(\frac{r}{\nu + r}, r\Big).$$

Conditional on the sporozoite batch size $S_j$, the number of hypnozoites $H_j$ that are destined to activate is sampled from a binomial distribution

$$H_j \big| S_j \sim \text{Binomial}\big(S_j, p_{\text{rel}}\big).$$

Granted $H_j < S_j$, that is, at least one sporozoite established by bite $j$ undergoes immediate development, we record a primary infection at time $T_j$. We additionally record $H_j$ relapses, with relapse $i$ of bite $j$ occurring at time

$$R_j^{(i)} = T_j + A_j^{(i)} \text{ where } A_j^{(i)} \overset{\text{i.i.d.}}{\sim} \text{Exponential}(\eta)$$

with the rate parametrisation of the exponential distribution.

For each window $k = 1, \ldots, n_{\text{obs}}$, we recover a binary infection state $I_k$ with $I_k = 1$ if at least one recurrence was recorded in the interval $[365 \cdot a + (k-1)T, 365 \cdot a + kT)$ and $I_k = 0$ otherwise. To generate a quaternary infection sequence $\mathbf{C} \in \{M, H, B, C\}^{n_{\text{obs}}}$ corrected for post-treatment prophylaxis (with $C_k = M$ indicating masking due to propyhlaxis in window $k$; $C_k = C$ indicating a recurrence manifesting in window $k$; $C_k = B$ indicating a hypnozoite activation or immediate sporozoite development event with delayed manifestation due to prophylactic bunching; and $C_k = H$ otherwise), we iterate across infection windows $k = 1, \ldots, n_{\text{obs}}$ and perform the following steps:

- If $C_k \in \{H, M, B\}$ has already been assigned, we leave as is.
- If $C_k = C$ or $C_k$ is yet to be assigned and $I_k = 1$, we set $C_k = C$ and $C_{k+1} = M$ i.e. we model a period of complete prophylactic protection spanning 10 days. Additionally, if $I_{k+1} \neq 0$ or $I_{k+2} \neq 0$, we set $C_{k+2} = B$ and $C_{k+3} = C$, corresponding to a prophylactic bunching period spanning 20 days.
- If $C_k$ is yet to be assigned and $I_k = 0$, we set $C_k = H$.

By mapping $C$ to $B$, we recover a ternary infection sequence $\mathbf{C}' \in \{M, H, B\}^{n_{\text{obs}}}$.

###### D.1.2.2 An inferential framework assuming geometric sporozoite batches

We estimate the parameters $\{\lambda, \eta, \nu\}$ using the Metropolis-Hastings algorithm. Inference is performed under the assumption of geometrically-distributed batch sizes, with the likelihood of observing a given ternary sequence $\mathbf{C}' \in \{H, M, B\}^{n_{\text{obs}}}$ for an individual of age $a$ computed using the formulae derived in Appendix B.2. In light of computational constraints, we restrict inference to simulated individuals with at most 12 unique recurrences. We take flat improper priors on $(0, \infty)$ for each parameter $\{\lambda, \nu, \eta\}$, with (symmetric) rectified normal proposals

$$\lambda^* \sim \mathcal{N}^{\mathcal{R}}(\lambda', 0.02/365) \qquad \nu^* \sim \mathcal{N}^{\mathcal{R}}(\nu', 0.2) \qquad \eta^* \sim \mathcal{N}^{\mathcal{R}}(\eta', 1/2000),$$

and initial values are sampled from uniform distributions

$$\lambda' \sim U[0.1/365, 1/365] \qquad \nu' \sim [0.5, 8] \qquad \eta' \sim [1/500, 1/50].$$

We run 4 chains over 32000 iterations, discarding the initial 1000 iterations of each chain as the burn-in period.

###### D.1.2.3 Recapitulating the burden of primary infection and relapse

Notwithstanding parametric misspecification of the sporozoite batch size, we seek to recapitulate the respective burden of relapse and primary infection. As such, we introduce:

- The force of primary bloodstream infection, defined to be the product of the force of inoculation $\lambda$ and the probability $b_{\text{prim}}$ of a primary infection associated with each bite
$$\lambda_{\text{prim}}(r) := \lambda \Big[ 1 - \Big( 1 + \frac{\nu}{r}(1 - p_{\text{rel}}) \Big)^{-r} \Big]. \tag{D.2}$$
- The relative burden of relapse vs primary infection, defined to be the ratio of the expected number of activating hypnozoites $\nu p_{\text{rel}}$ and primary infections $b_{\text{prim}}$ per bite
$$Y_{\text{relapse:prim}}(r) := \frac{\nu p_{\text{rel}}}{1 - \Big( 1 + \frac{\nu}{r}(1 - p_{\text{rel}}) \Big)^{-r}}. \tag{D.3}$$

In particular, we seek to see whether the respective truth values $\lambda_{\text{prim}}(r)$ and $Y_{\text{relapse:prim}}(r)$ for negative binomial sporozoite inocula (which are over-dispersed relative to the geometric distribution in the case $r < 1$, and under-dispersed if $r > 1$) can be recapitulated by the quantities $\lambda_{\text{prim}}(1)$ and $Y_{\text{relapse:prim}}(1)$ estimated under the inferential framework predicated on geometric batch sizes.

##### D.1.3 Simulation study results

Marginal posteriors for a series of epidemiologically-plausible parameter sets are shown in Figure D.1. In short, estimates for the force of inoculation $\lambda$ and the mean batch size $\nu$ are sensitive to parameteric misspecification of sporozoite inocula. For over-dispersed distributions ($r < 1$), $\lambda$ is systematically under-estimated to compensate for unaccounted zero-inflation in sporozoite inoc-

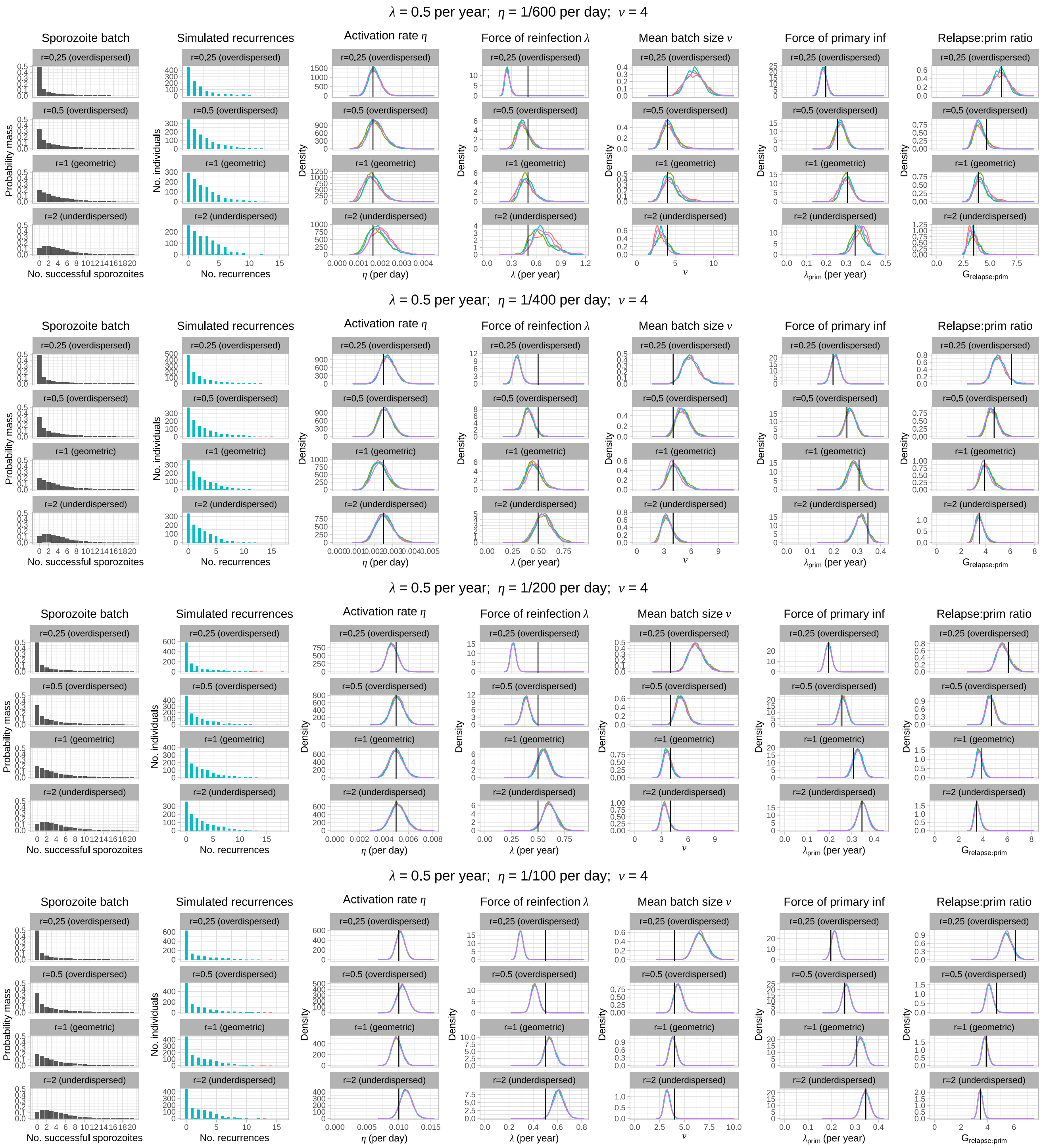

**(a)** $\lambda = 0.5/365$ day$^{-1}$, $\eta \in \{1/600, 1/400, 1/200, 1/100\}$ day$^{-1}$, $\nu = 4$, $r \in \{0.25, 0.5, 1, 2\}$, $p_{\mathrm{rel}} = 0.6$

**Figure D.1:** Simulation study results. Marginal posteriors are stratified by chain.

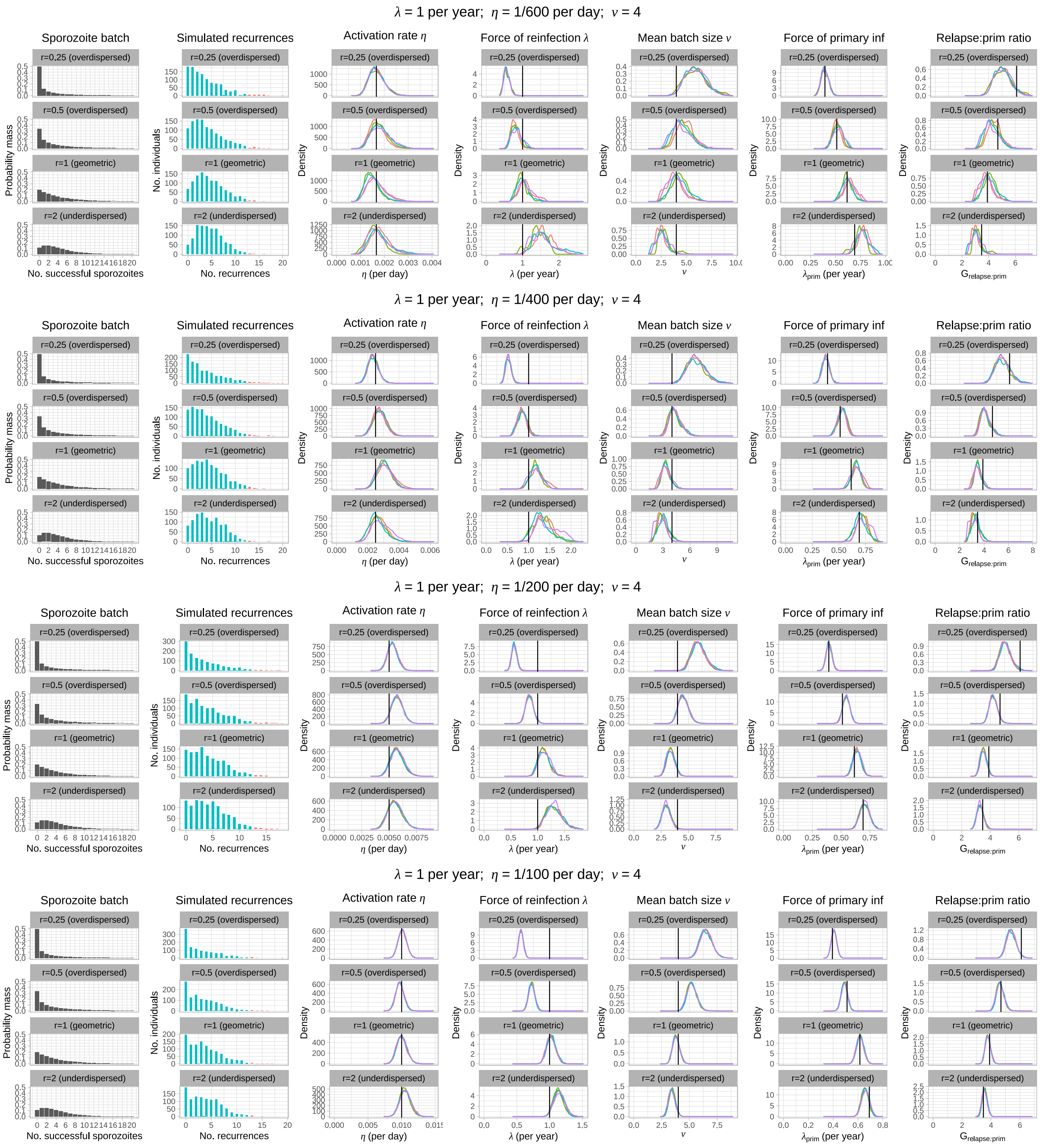

**(b)** $\lambda = 1/365 \text{ day}^{-1}$, $\eta \in \{1/600, 1/400, 1/200, 1/100\} \text{ day}^{-1}$, $\nu = 4$, $r \in \{0.25, 0.5, 1, 2\}$, $p_{\text{rel}} = 0.6$

**Figure D.1:** Simulation study results. Marginal posteriors are stratified by chain.

ula; while $\nu$ is systematically over-estimated to better capture over-dispersion in the simulated risk of recurrence. The converse applies for under-dispersed distributions ($r > 1$). Estimates for the hypnozoite activation rate $\eta$, however, appear to be more robust with proportionally little systematic bias apparent for the simulated set of non-geometric sporozoite distributions.

Geometric batch sizes offer reasonable flexibility to recapitulate the burden of relapse vs primary infection in spite of parametric misspecification of the sporozoite batch size. Estimates for the force of *primary* infection $\lambda_{\text{prim}}$ appear to exhibit little systematic bias. For over-dispersed sporozoite distributions with long right tails (i.e. small $r$), estimates for the ratio of relapse vs primary infection per bite $G_{\text{relapse:prim}}$ tend to be downward-biased (but less severely than estimates for the mean sporozoite batch size $\nu$): we postulate that this occurs due to redundancy in hypnozoite activation events following large sporozoite inoculations.

#### D.2 Hypnozoite fating probability $p_{\text{rel}}$

The probability $p_{\text{rel}} = 0.4$ that a "successful" sporozoite will form a hypnozoite that is destined to activate vs undergo immediate activation is informed by *in vivo* and *in vitro* experiments for the Chesson strain of *P. vivax* [28], with origins in the island of New Guinea [29]. For completeness, we perform a sensitivity analysis for the hypnozoite fating probability $p_{\text{rel}} \in \{0.1, 0.25, 0.4, 0.6, 0.75, 0.9\}$, using the Metropolis-Hastings regime detailed in Appendix C.1.1 for the SPf66 vaccine trial data. Marginal posterior densities, stratified by chain, are shown in Figure D.2. In short, we attain consistent estimates for the hypnozoite activation rate $\eta$ and the mean *hypnozoite* batch size $\nu p_{\text{rel}}$ as the hypnozoite fating probability $p_{\text{rel}}$ is varied. While estimates for the force of inoculation (particularly in the first 200 days of the study) appear to be somewhat sensitive to the hypnozoite fating probability $p_{\text{rel}}$, estimates for the expected cumulative number of hypnozoites and primary infections acquired over the course of a year

$$B_{\text{total}} = \lambda\Big(1 - \frac{1}{1 + \nu(1 - p_{\text{rel}}} + \nu p_{\text{rel}}\Big)$$

(i.e. the product of the average yearly force of inoculation. and the expected number of primary infections and hypnozoites established by each bite) are generally consistent as $p_{\text{rel}}$ is varied.

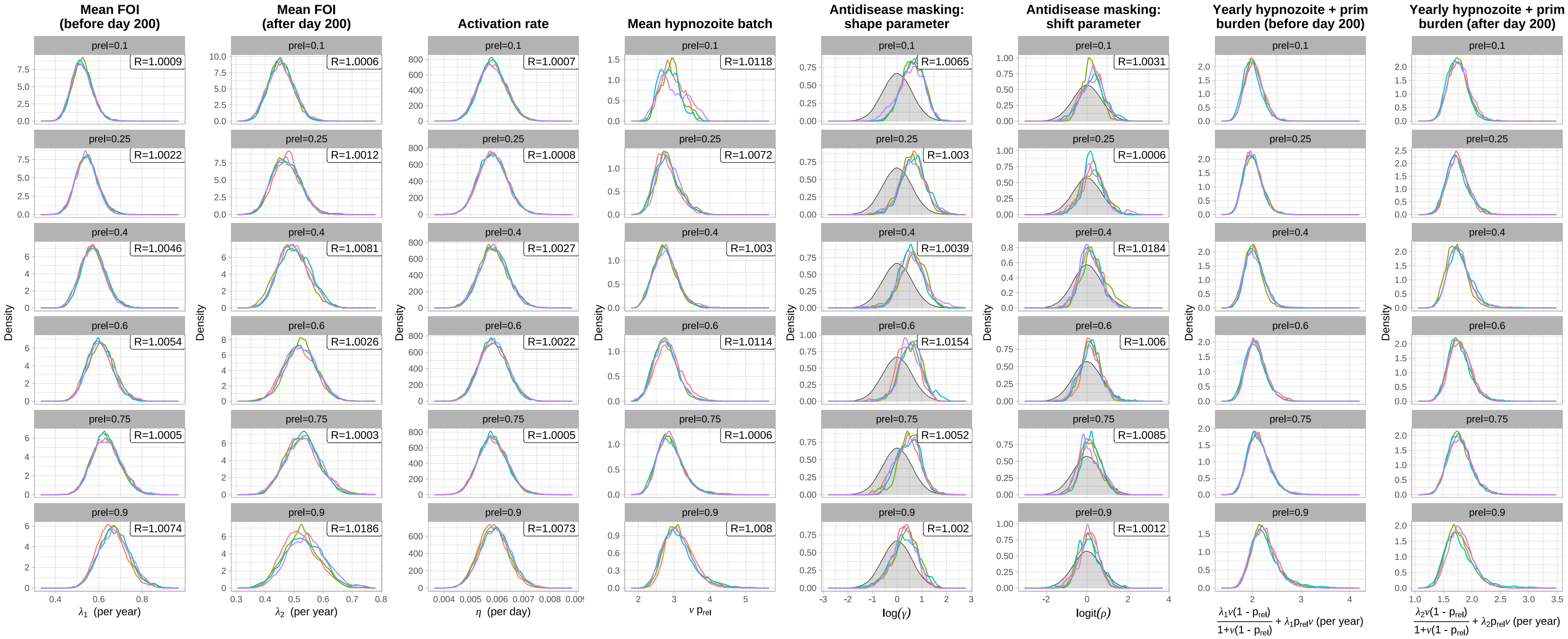

**Figure D.2:** Marginal posteriors for each estimated parameter, stratified by chain, for different hypnozoite fating probabilities $p_{\text{rel}} \in \{0.1, 0.25, 0.4, 0.6, 0.75, 0.9\}$; prior distributions for $\{\text{logit}(\rho), \log(\gamma)\}$ are shown in grey. Averaging over seasonal fluctuations, $\lambda_1$ represents the mean force of inoculation up to day 200 of the study period, while $\lambda_2$ represents the mean force of inoculation thereafter. $R$ denotes the Gelman-Rubin diagnostic, calculated using Equation (1.1) of [24] after discarding the burn-in period (i.e. the initial 20,000 iterations of each chain).

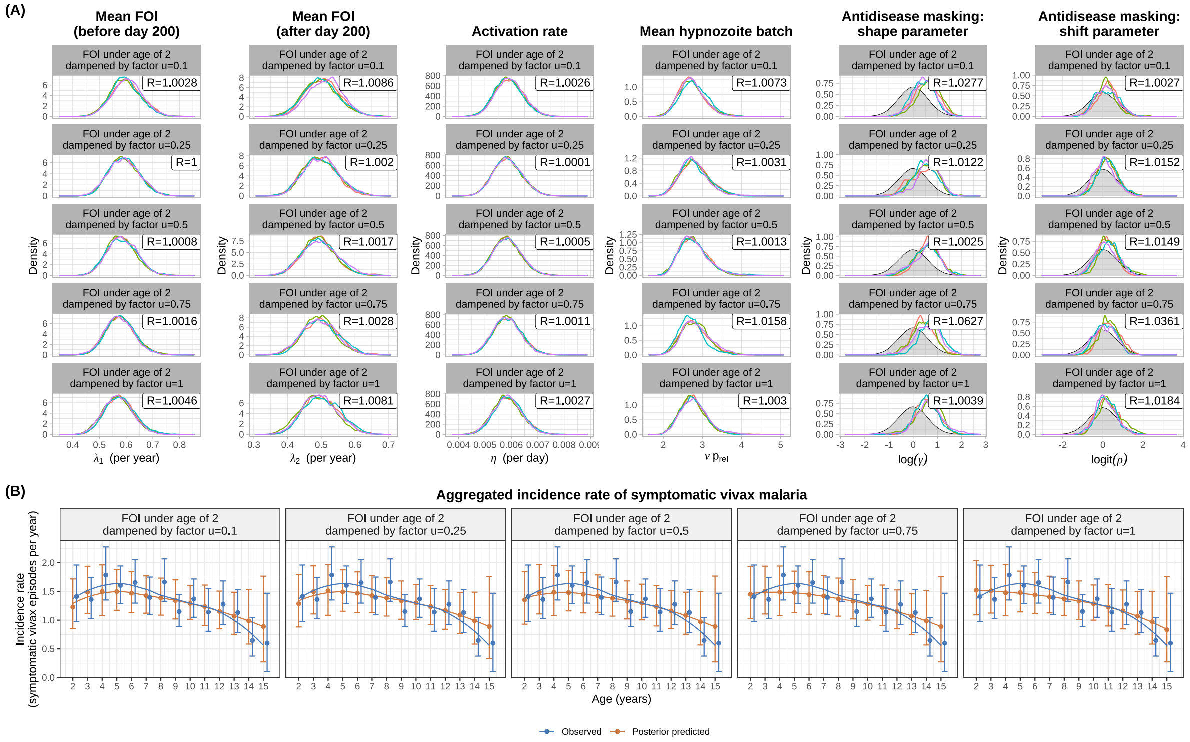

**Figure D.3:** Estimates accounting for a dampened force of inoculation (parametrised by a fixed factor $u$) between birth and age 2.

(A) Marginal posterior densities for each parameter, stratified by chain. $R$ denotes the Gelman-Rubin diagnostic.

(B) Observed vs posterior predicted incidence rate, stratified by age group. Error bars indicate 95% confidence intervals for empirical data (generated by bootstrapping with 2000 replicates), and 95% CIs for data simulated under the posterior. Smoothing splines based on median incidence rates (indicated with points) have been generated with `ggplot::geom_smooth` [30] using the method `loess` with default parameters.

#### D.3 Age stratification in the force of inoculation

It is plausible that non-monotonic age structure in the incidence of symptomatic vivax malaria is a consequence of age-stratification in mosquito inoculation rates. We therefore perform additional model fits under the assumption that each individual experiences a dampened force of inoculation from birth until age 2, adjusted by a fixed factor $u \in \{0.1, 0.25, 0.5, 0.75, 1\}$. In the extreme case $u = 0$, the hypnozoite reservoir accrues only from age 2 onwards, while in the case $u = 1$ there is no age-stratification in the force of inoculation. Model calibration is performed using the Metropolis-Hasting algorithm outlined in Appendix C.1.1 for the SPf66 vaccine trial data, with appropriate age-dependent adjustments to the seasonality vector $\mathbf{S}$ for each child. Posterior predictive symptomatic vivax recurrence data (discretised across $n_{\text{obs}} = 65$ windows, each of length $T = 10$ days) are simulated for 2000 parameter combinations $(\Lambda_1, \Lambda_2, \eta, \nu, \rho, \gamma)$ sampled uniformly at random from the posterior (without replacement) — retaining the age distribution and clinical follow-up pattern of the SPf66 cohort, and adjusting for post-treatment prophylaxis — using the procedure detailed in Appendix C.2.1, likewise with appropriate age-dependent adjustments to the seasonality vector $\mathbf{S}$.

Marginal posterior densities for the force of inoculation $\Lambda$; hypnozoite activation rate $\eta$; mean sporozoite batch size $\nu$; the shape parameter $\gamma$ for the age-dependent anti-disease masking probability; and the shift parameter $\rho$ (i.e. the proportion reduction in the probability of symptomatic malaria for 2 vs 15 year olds) appear to be insensitive to the dampening factor $u$ (Figure D.3A). However, dampening the force of inoculation from birth to age 2 ($u < 1$) better recapitulates non-monotonicity in the age-stratified incidence rate of symptomatic vivax malaria (Figure D.3B). Given the estimated time-scale of hypnozoite carriage (approximately 5 months on average), we note that this non-montonicity is governed most strongly by the relative reduction in the force of inoculation between the ages of 1 and 2: assuming a dampened force of inoculation from birth to age 1, for instance, would not necessarily give rise to non-monotonic age structure in the symptomatic incidence rate.

### Appendix E

### Quantities of epidemiological interest

#### E.1 Recurrences following a single infective bite

Conditional on a hypnozoite batch of size $H = h \geq m$, the $m^{\text{th}}$ inter-relapse interval

$$T_m|_{H=h\geq m} \sim \text{Exponential}(\eta(h-m+1))$$

(where we have adopted the rate parametrisation of the exponential distribution). Using the law of total expectation, we evaluate the moments

$$\mathbb{E}[T_m|H \geq m] = \frac{\sum_{h=m}^{\infty} \frac{1}{\eta(h-m+1)} \left(\frac{\nu p_{\text{rel}}}{1+\nu p_{\text{rel}}}\right)^h}{\sum_{h=m}^{\infty} \left(\frac{\nu p_{\text{rel}}}{1+\nu p_{\text{rel}}}\right)^h} = \frac{\log(1+\nu p_{\text{rel}})}{\eta \nu p_{\text{rel}}}$$

$$\mathbb{E}[T_m^2|H \geq m] = \frac{\sum_{h=m}^{\infty} \frac{2}{\eta^2(h-m+1)^2} \left(\frac{\nu p_{\text{rel}}}{1+\nu p_{\text{rel}}}\right)^h}{\sum_{h=m}^{\infty} \left(\frac{\nu p_{\text{rel}}}{1+\nu p_{\text{rel}}}\right)^h} = \frac{2\text{Li}_2\left(\frac{\nu p_{\text{rel}}}{1+\nu p_{\text{rel}}}\right)}{\eta^2 \nu p_{\text{rel}}},$$

where $\text{Li}_2(\cdot)$ denotes the polylogarithm function of order 2. The coefficient of variation for the $m^{\text{th}}$ inter-relapse, given it occurs, can therefore be written

$$\text{CV}(T_m|H \geq m) = \frac{\text{SD}(T_m|H \geq m)}{\mathbb{E}[T_m|H \geq m]} = \frac{\sqrt{2\nu p_{\text{rel}} \cdot \text{Li}_2\left(\frac{\nu p_{\text{rel}}}{1+\nu p_{\text{rel}}}\right) - \log(1+\nu p_{\text{rel}})^2}}{\log(1+\nu p_{\text{rel}})}.$$

Treating the time to first recurrence $T_1$ as a baseline, we also consider the difference between the $m^{\text{th}}$ and first inter-relapse intervals $T_m - T_1$. Conditional on the hypnozoite batch size $H = h \geq m$, we note that $T_m$ and $T_1$ are conditionally independent. A similar application of the law of total expectation thus yields

$$\mathbb{E}[T_m - T_1|H \geq m] = \frac{\sum_{h=m}^{\infty} \left[\frac{1}{\eta(h-m+1)} - \frac{1}{h}\right] \left(\frac{\nu p_{\text{rel}}}{1+\nu p_{\text{rel}}}\right)^h}{\sum_{h=m}^{\infty} \left(\frac{\nu p_{\text{rel}}}{1+\nu p_{\text{rel}}}\right)^h}$$

$$= \frac{\log(1+\nu p_{\text{rel}})}{\nu p_{\text{rel}}} - \frac{\Phi\left(\frac{\nu p_{\text{rel}}}{1+\nu p_{\text{rel}}}, 1, m\right)}{1+\nu p_{\text{rel}}}$$

$$\text{Var}[(T_m - T_1)^2 | H \geq m] = \frac{\sum_{h=m}^{\infty} \frac{2}{\eta^2}\left[\frac{1}{(h-m+1)^2} + \frac{1}{h^2}\right]\left(\frac{\nu p_{\text{rel}}}{1+\nu p_{\text{rel}}}\right)^h}{\sum_{h=m}^{\infty}\left(\frac{\nu p_{\text{rel}}}{1+\nu p_{\text{rel}}}\right)^h}$$

$$= \frac{\text{Li}_2\left(\frac{\nu p_{\text{rel}}}{1+\nu p_{\text{rel}}}\right)}{\nu p_{\text{rel}}} + \frac{\Phi\left(\frac{\nu p_{\text{rel}}}{1+\nu p_{\text{rel}}}, 2, m\right)}{1+\nu p_{\text{rel}}}$$

where $\Phi(z, s, a)$ denotes the Hurwitz–Lerch transcendent.

Here, all infinite sums have been evaluated symbolically using Mathematica [31].

#### E.2 Metrics related to the hypnozoite burden

##### E.2.1 Size of the hypnozoite reservoir

From Equation (34) of [2], the size of the hypnozoite reservoir $H(t)$ after a period of accrual $t$ has PGF

$$\mathbb{E}\left[z^{H(t)}\right] = \exp\left\{\int_0^t \lambda(\tau)\left[-1 + \frac{1}{1+\nu p_{\text{rel}}(1-z)e^{-\eta(t-\tau)}}\right]d\tau\right\}. \tag{E.1}$$

Under a piecewise constant FORI $\lambda_i$ over uniform windows of length $T$, we can evaluate Equation (E.1) analytically to yield the size of the hypnozoite reservoir $H(n)$ at the end of the $n^{\text{th}}$ window, or equivalently, after period $nT$ of accrual:

$$\mathbb{E}\left[z^{H(n)}\right] = \prod_{i=1}^{n}\left[\frac{1+\nu p_{\text{rel}}(1-z)e^{-\eta(n-i+1)T}}{1+\nu p_{\text{rel}}(1-z)e^{-\eta(n-i)T}}\right]^{\frac{\lambda_i}{\eta}}.$$

Using Equations (74) and (75) of [2], we can write the PMF for $H(n)$ using complete exponential Bell polynomials $B_h$:

$$P\big(H(n) = h\big) = \frac{1}{n!}\left(\prod_{i=1}^{n}\left[\frac{1+\nu p_{\text{rel}}e^{-\eta(n-i+1)T}}{1+\nu p_{\text{rel}}e^{-\eta(n-i)T}}\right]^{\frac{\lambda_i}{\eta}}\right)\cdot B_h\big(x_1(n), \dots, x_h(n)\big) \tag{E.2}$$

where

$$x_k(n) = \frac{1}{\eta}(k-1)!\sum_{i=1}^{n}\lambda_i\left[\nu p_{\text{rel}}e^{-\eta(n-i+1)T}\right]^k \cdot \left[\frac{1}{(e^{-\eta T}+\nu p_{\text{rel}}e^{-\eta(n-i+1)T})^k} - \frac{1}{(1+\nu p_{\text{rel}}e^{-\eta(n-i+1)T})^k}\right]. \tag{E.3}$$

At stationarity (that is, in the limit $t \to \infty$) under a constant FORI $\lambda$, we show in [2] (Equation (37)) that the size of the hypnozoite reservoir $H^*$ follows a negative binomial distribution with

PGF

$$\mathbb{E}\left[z^{H*}\right] = \big(1 + \nu p_{\text{rel}}(1-z)\big)^{-\frac{\lambda}{\eta}} \tag{E.4}$$

and PMF

$$P(H^* = n) = \frac{\Gamma\big(\frac{\lambda}{\eta} + n\big)}{n! \cdot \Gamma\big(\frac{\lambda}{\eta}\big)} \frac{(p_{\text{rel}}\nu)^n}{(1 + p_{\text{rel}}\nu)^{n + \frac{\lambda}{\eta}}}$$

where $\Gamma(\cdot)$ denotes the Gamma function.

##### E.2.2 Time to hypnozoite clearance following the cessation of mosquito transmission

Suppose mosquito transmission is curbed completely from time $t = nT$ onwards. Given an individual harbours precisely $h$ hypnozoites, the total time to hypnozoite clearance $T_{\text{clear}}$ for an individual has cdf

$$P\big[T_{\text{clear}} \leq u \,|\, H(n) = h\big] = \big(1 - e^{-\eta u}\big)^h.$$

Using the law of total expectation, it thus follows that

$$P\big[T_{\text{clear}} \leq u\big] = \sum_{h=0}^{\infty} P(H(n) = h) \cdot \big(1 - e^{-\eta u}\big)^h = \mathbb{E}\left[(1 - e^{-\eta u})^{H(n)}\right] \tag{E.5}$$

$$= \prod_{i=1}^{n} \left[\frac{1 + \nu p_{\text{rel}} e^{-\eta[(n-i+1)T+u]}}{1 + \nu p_{\text{rel}} e^{-\eta[(n-i)T+u]}}\right]^{\frac{\lambda_i}{\eta}}. \tag{E.6}$$

For a stationary hypnozoite reservoir under a constant force of inoculation $\lambda$, we obtain

$$P\big[T^*_{\text{clear}} \leq u\big] = \big(1 + \nu p_{\text{rel}} e^{-\eta u}\big)^{-\frac{\lambda}{\eta}} \tag{E.7}$$

by substituting Equation (E.4) into (E.5).

To achieve spontaneous hypnozoite clearance with probability $c$ in each individual, mosquito-to-human transmission would therefore need to be interrupted for time

$$T_{\text{interrupt}}(c) = \max\left\{0,\, \frac{1}{\eta} \log\left(\frac{\nu p_{\text{rel}}}{c^{-\frac{\eta}{\lambda}} - 1}\right)\right\}. \tag{E.8}$$

#### E.3 Recent recurrence as a predictor of hypnozoite carriage

##### E.3.1 Joint distribution of the recurrence and hypnozoite burden

Here, we derive the joint distribution of number of recurrences $N_i$ in discretised windows $i$, in addition to the hypnozoite burden $R_n$ at a desired endpoint $n$. We can derive the multivariate PGF for $N_i$, $R_n$ through a slight modification (highlighted in red) of the argument detailed in Appendix B.2. Specifically, given a successful sporozoite is inoculated at time $t_{j-1} < \tau < t_j$, the multivariate PGF for the number of hypnozoite activation $H_i$ and immediate sporozoite development $F_i$ events in each interval $(t_{i-1}, t_i]$, in addition to the hypnozoite burden in window $n$ can be written

$$\begin{aligned}\mathbb{E}\left[ w^{R_n} \prod_{i=1}^{n} x_i^{H_i} y_i^{F_i} \,\middle|\, \text{sporozoite established at time } t_{j-1} < \tau < t_j \right] \\ = \underbrace{(1-p_{\text{rel}})y_j}_{\text{immediate development}} + \underbrace{p_{\text{rel}}[1 - B(t_n - \tau)]w}_{\text{hypnozoite activates after } t_n} + \underbrace{p_{\text{rel}} B(t_j - \tau) x_j}_{\text{hypnozoite activates in } (\tau, t_j]} \\ + \sum_{k=j+1}^{n} \underbrace{p_{\text{rel}}\big[B(t_k - \tau) - B(t_{k-1} - \tau)\big] x_k}_{\text{hypnozoite activates in } (t_{k-1}, t_k]}.\end{aligned}$$

An identical argument then allows us to recover the joint PGF of the hypnozoite burden $R_{n_{\text{obs}}}$ at the end of an observation period spanning $n_{\text{obs}}$ windows, in addition to the number of hypnozoite activation and sporozoite developments $U_i$ with the potential to cause clinical symptoms in window $i$

$$U_i | N_{i+n_{\text{age}}} \sim \text{Binomial}\big(N_{i+n_{\text{age}}}, p_{\text{clin}}\big).$$

For uniformly separated windows $t_j = jT$, we obtain

$$\begin{aligned}\mathbb{E}\left[ w^{R_{n_{\text{obs}}}} \prod_{i=1}^{n_{\text{obs}}} x_i^{U_i} \right] = \prod_{j=1}^{n_{\text{age}}} e^{-\lambda_j T} \left( 1 - \frac{1 - e^{-\eta T}}{1 + \nu p_{\text{rel}} p_{\text{clin}} e^{-\eta T (n_{\text{age}} - j)} \left[ e^{-\eta T} - w e^{-\eta T (n_{\text{obs}} + 1)} - (1 - e^{-\eta T}) \sum_{k=1}^{n_{\text{obs}}} x_k \cdot e^{-\eta T k} \right]} \right)^{-\frac{\lambda_j}{\eta}} \\ \prod_{j=1}^{n_{\text{obs}}} e^{-\lambda_j T} \left( 1 - \frac{\big(1 + \nu p_{\text{rel}} p_{\text{clin}} (1 - x_j)\big)(1 - e^{-\eta T})}{1 + \nu p_{\text{rel}} p_{\text{clin}} - \nu p_{\text{rel}} p_{\text{clin}} \left[ w e^{-\eta (n_{\text{obs}} - j + 1) T} + (1 - e^{-\eta T}) \sum_{k=j}^{n_{\text{obs}}} x_k e^{-\eta (k - j) T} \right]} \right)^{-\frac{\lambda_j (1 - p_{\text{clin}} + p_{\text{clin}} x_j)}{\eta (1 + \nu p_{\text{rel}} p_{\text{clin}} (1 - x_j))}} \\ \left( 1 - \frac{\big(1 + \nu(1 - p_{\text{rel}}(1 - p_{\text{clin}}) - p_{\text{rel}} p_{\text{clin}} x_j)\big)(1 - e^{-\eta T})}{1 + \nu\big(1 - p_{\text{rel}}(1 - p_{\text{clin}})\big) - \nu p_{\text{rel}} p_{\text{clin}} \left[ w e^{-\eta (n_{\text{obs}} - j + 1) T} + (1 - e^{-\eta T}) \sum_{k=j}^{n_{\text{obs}}} x_k e^{-\eta (k - j) T} \right]} \right)^{-\frac{\lambda_j p_{\text{clin}} (1 - x_j)}{\eta (1 + \nu (1 - p_{\text{rel}} (1 - p_{\text{clin}}) - p_{\text{rel}} p_{\text{cli}} \text{[illegible]}}}\end{aligned} \tag{E.9}$$

Differences to the joint PGF for $U_i$ (Equation (B.12)) are highlighted in red.

If we allow the hypnozoite reservoir to reach stationarity prior to the study period, that is, we take the limit $n_{\text{age}} \to \infty$ under a constant historical force of inoculation $\lambda$ then we can write

$$
\begin{aligned}
\mathbb{E}\left[w^{R_{n_{\text{obs}}}} \prod_{i=1}^{n_{\text{obs}}} x_i^{U_i}\right] = &\left(1 + \nu p_{\text{rel}} p_{\text{clin}} e^{-\eta n_{\text{obs}} T}(1 - w) + \nu p_{\text{rel}} p_{\text{clin}} \sum_{k=1}^{n_{\text{obs}}} e^{-\eta(k-1)T}(1 - e^{-\eta T})(1 - z_k)\right)^{-\frac{\lambda}{\eta}} \\
&\prod_{j=1}^{n_{\text{obs}}} e^{-\lambda_j T}\left(1 - \frac{(1 + \nu p_{\text{rel}} p_{\text{clin}}(1 - x_j))(1 - e^{-\eta T})}{1 + \nu p_{\text{rel}} p_{\text{clin}} - \nu p_{\text{rel}} p_{\text{clin}}\left[w e^{-\eta(n_{\text{obs}} - j + 1)T} + (1 - e^{-\eta T}) \sum_{k=j}^{n_{\text{obs}}} x_k e^{-\eta(k-j)T}\right]}\right)^{-\frac{\lambda_j(1 - p_{\text{clin}} + p_{\text{clin}} x_j)}{\eta(1 + \nu p_{\text{rel}} p_{\text{clin}}(1 - x_j))}} \\
&\left(1 - \frac{(1 + \nu(1 - p_{\text{rel}}(1 - p_{\text{clin}}) - p_{\text{rel}} p_{\text{clin}} x_j))(1 - e^{-\eta T})}{1 + \nu(1 - p_{\text{rel}}(1 - p_{\text{clin}})) - \nu p_{\text{rel}} p_{\text{clin}}\left[w e^{-\eta(n_{\text{obs}} - j + 1)T} + (1 - e^{-\eta T}) \sum_{k=j}^{n_{\text{obs}}} x_k e^{-\eta(k-j)T}\right]}\right)^{-\frac{\lambda_j p_{\text{clin}}(1 - x_j)}{\eta(1 + \nu(1 - p_{\text{rel}}(1 - p_{\text{clin}}) - p_{\text{rel}} p_{\text{clin}} x_j))}}.
\end{aligned}
\tag{E.10}
$$

##### E.3.2 Accuracy of recent recurrence as a predictor of hypnozoite carriage

Under a constant force of inoculation $\lambda$, where the hypnozoite reservoir is likewise modelled to reach stationarity, the joint PGF for the number of hypnozoite activation and/or immediate sporozoite development events $U$ in a window of length $T$, and the size of the hypnozoite resevoir $R$ at the end of that window, can be written

$$
\begin{aligned}
f(x, w) := \mathbb{E}\left[x^U w^R\right] = &\left(1 + \nu p_{\text{rel}} - \nu p_{\text{rel}}\left[w e^{-\eta T} + x(1 - e^{-\eta T})\right]\right)^{-\frac{\lambda}{\eta}} \\
&e^{-\lambda T}\left(1 - \frac{(1 + \nu p_{\text{rel}}(1 - x))(1 - e^{-\eta T})}{1 + \nu p_{\text{rel}} - \nu p_{\text{rel}}\left[w e^{-\eta T} + x(1 - e^{-\eta T})\right]}\right)^{-\frac{\lambda x}{\eta(1 + \nu p_{\text{rel}}(1 - x))}} \left(1 - \frac{(1 + \nu - \nu p_{\text{rel}} x)(1 - e^{-\eta T})}{1 + \nu - \nu p_{\text{rel}}\left[w e^{-\eta T} + x(1 - e^{-\eta T})\right]}\right)^{-\frac{\lambda(1 - x)}{\eta(1 + \nu - \nu p_{\text{rel}} x)}}
\end{aligned}
\tag{E.11}
$$

where we have set $n_{\text{obs}} = 1$, $p_{\text{clin}} = 1$ in Equation (E.9).

Suppose a serological test can, with perfect accuracy, detect whether an individual experienced a bloodstream infection in the preceding $T$ day window. Under the above conditions, the specificity of this test in predicting hypnozoite carriage, or equivalently, the conditional probability that a non-hypnozoite carrier has not experienced a recurrence in the preceding $T$ day window, can be computed using Equation (E.11):

$$
h_{\text{spec}}(\lambda) := \frac{P(U = 0, R = 0)}{P(R = 0)} = \frac{f(0, 0)}{f(1, 0)} = e^{-\frac{\lambda T \nu}{1 + \nu}}. \tag{E.12}
$$

Likewise, the sensitivity of the test, or equivalently, the conditional probability that a hypnozoite carrier has experienced a recurrence in the preceding $T$ day window, can be written

$$
\begin{aligned}
h_{\text{sens}}(\lambda) &:= \frac{P(U > 0, R > 0)}{P(R > 0)} = \frac{1 - f(0, 1) - f(1, 0) + f(0, 0)}{1 - f(1, 0)} \\
&= \frac{1 - (1 + \nu p_{\text{rel}})^{-\frac{\lambda}{\eta}}\left[1 - e^{-\frac{\lambda T \nu}{1 + \nu}}\right] - e^{-\lambda T}\left(1 + \nu p_{\text{rel}}(1 - e^{-\eta T})\right)^{-\frac{\lambda}{\eta}}\left(1 - \frac{(1 + \nu)(1 - e^{-\eta T})}{1 + \nu - \nu p_{\text{rel}} e^{-\eta T}}\right)^{-\frac{\lambda}{\eta(1 + \nu)}}}{1 - (1 + \nu p_{\text{rel}})^{-\frac{\lambda}{\eta}}}.
\end{aligned}
\tag{E.13}
$$

The positive predictive value, or the conditional probability of hypnozite carriage given a recurrence within the preceding $T$ day window is given by

$$\frac{P(U>0,R>0)}{P(U>0)} = \frac{1-f(0,1)-f(1,0)+f(0,0)}{1-f(0,1)}$$

$$= \frac{1-(1+\nu p_{\text{rel}})^{-\frac{\lambda}{\eta}}\left[1-e^{-\frac{\lambda T\nu}{1+\nu}}\right]-e^{-\lambda T}\big(1+\nu p_{\text{rel}}(1-e^{-\eta T})\big)^{-\frac{\lambda}{\eta}}\left(1-\frac{(1+\nu)(1-e^{-\eta T})}{1+\nu-\nu p_{\text{rel}}e^{-\eta T}}\right)^{-\frac{\lambda}{\eta(1+\nu)}}}{1-e^{-\lambda T}\big(1+\nu p_{\text{rel}}(1-e^{-\eta T})\big)^{-\frac{\lambda}{\eta}}\left(1-\frac{(1+\nu)(1-e^{-\eta T})}{1+\nu-\nu p_{\text{rel}}e^{-\eta T}}\right)^{-\frac{\lambda}{\eta(1+\nu)}}}, \tag{E.14}$$

while the false omission rate takes the form

$$\frac{P(U=0,R>0)}{P(U=0)} = 1-\frac{f(0,0)}{f(0,1)}$$

$$= 1-e^{\frac{\lambda T}{1+\nu}}(1+\nu p_{\text{rel}})^{-\frac{\lambda}{\eta}}\big(1+\nu p_{\text{rel}}(1-e^{-\eta T})\big)^{\frac{\lambda}{\eta}}\left(1-\frac{(1+\nu)(1-e^{-\eta T})}{1+\nu-\nu p_{\text{rel}}e^{-\eta T}}\right)^{\frac{\lambda}{\eta(1+\nu)}}. \tag{E.15}$$

##### E.3.3 Allowing for population hetereogeniety in the force of inoculation

To accommodate population heterogeneity, we model the force of inoculation $\lambda$ to follow a Gamma distribution [19]

$$\lambda \sim \text{Gamma}(\kappa,\theta)$$

where we have adopted the shape-scale parametrisation. The mean force of inoculation in the population takes the form $k\theta$, while scale parameter $\theta$ can be interpreted as the variance-to-mean ratio for population heterogeneity in the force of inoculation.

For each individual in the population, we sample a force of inoculation $\lambda^*$ from this Gamma distribution, and model the hypnozoite reservoir to reach stationarity under the inoculation rate $\lambda^*$. Then by the law of total probability, the specificity of recent recurrence (within a preceding $T$ window) as a predictor of hypnozoite carriage — which we compute as the conditional probability that a randomly-sampled individual from the population does not carry hypnozoites, given they have not experienced any recurrences in the preceding $T$ days — follows from the law of total probability

$$h_{\text{neg}}^{(\text{het})}(\kappa,\theta) = \int_0^\infty h_{\text{spec}}(\lambda)\cdot p(\lambda|\kappa,\theta)d\lambda = \left(1+\frac{\theta T\nu}{1+\nu}\right)^{-\kappa} \tag{E.16}$$

where we have used Equation (E.12) and recognised the moment generating function for the Gamma distribution. Similarly, the sensitivity of recent recurrence (within a preceding $T$ day window) as a predictor of hypnozoite carriage — which we compute as the conditional probability that a randomly-sampled individual from the population carries hypnozoites, given they have experienced at least one recurrence in the preceding $T$ days — can be computed

$$\begin{aligned} h_{\text{pos}}^{(\text{het})}(\kappa,\theta) &= \int_0^\infty h_{\text{sens}}(\lambda)\cdot p(\lambda|\kappa,\theta)d\lambda \\ &= 1 + \left(\frac{\eta}{\theta\log(1+\nu p_{\text{rel}})}\right)^{\kappa}\Bigg[\zeta\left(\kappa, 1+\frac{\eta}{\log(1+\nu p_{\text{rel}})}\left(\frac{1}{\theta}+\frac{T\nu}{1+\nu}\right)\right) \\ &\qquad - \zeta\left(\kappa, \frac{\frac{\eta}{\theta}+\eta T+\log\left(1+\nu p_{\text{rel}}(1-e^{-\eta T})\right)+\frac{1}{1+\nu}\log\left(1-\frac{(1+\nu)(1-e^{-\eta T})}{1+\nu-\nu p_{\text{rel}}e^{-\eta T}}\right)}{\log(1+\nu p_{\text{rel}})}\right)\Bigg] \end{aligned} \tag{E.17}$$

using Equation (E.13) and integral 3.411.7 of [23], where $\zeta(\cdot,\cdot)$ denotes the Hurwitz zeta function.

###### E.3.3.1 A more intuitive formulation of population heterogeneity

To yield a more intuitive measure of population heterogeneity, we can reparameterise the Gamma distribution by the force of inoculation averaged over the population $\Lambda = k\theta$ and the proportion of bites $P_{0.8}$ that are experienced by the 20% of individuals subject to the highest transmission intensity. Denote by $X_{0.8}$ the 0.8 quantile of the Gamma distribution with shape $k$ and scale $\theta$, such that

$$\frac{\Gamma\big(k, \frac{X_{0.8}}{\theta}\big)}{\Gamma(k)} = 0.2$$

where $\Gamma(\cdot)$ denotes the Gamma function, and $\Gamma(\cdot,\cdot)$ denotes the upper incomplete Gamma function. The proportion of bites experienced by the 20% of individuals subject to the highest transmission intensity takes the form

$$P_{0.8} := \frac{1}{k\theta}\int_{X_{0.8}}^\infty \frac{1}{\Gamma(k)\theta^k}x^k e^{-\frac{x}{\theta}}dx = \frac{\Gamma\big(k+1, \frac{X_{0.8}}{\theta}\big)}{\Gamma(k+1)}.$$

Using a standard recurrence relation for the upper incomplete Gamma function (identity 8.356.2 of [23]), we can write

$$P_{0.8} = \frac{k\Gamma\big(k, \frac{X_{0.8}}{\theta}\big) + \big(\frac{X_{0.8}}{\theta}\big)^k e^{-\frac{X_{0.8}}{\theta}}}{\Gamma(k+1)} = 0.2 + \frac{1}{\Gamma(k+1)\theta^k}X_{0.8}^k e^{-\frac{X_{0.8}}{\theta}}. \tag{E.18}$$

##### E.3.4 Accuracy of an imperfect serological test for recent recurrence as a predictor of hypnozoite carriage

Now, consider an imperfect serological test with specificity $r_{\text{spec}}$ and sensitiviy $r_{\text{sens}}$ as a predictor for recent recurrence within a preceding $T$ day window. Denote by $R' = \mathbb{1}\{R \geq 0\}$ the indicator that an individual harbours hypnozoites at the time of testing, and $U' = \mathbb{1}\{U \geq 0\}$ the indicator that an individual experiences at least one recurrence in the preceding $T$ day window. We make the assumption that

$$\begin{aligned} r_{\text{sens}} &= P(\text{positive test} \,|\, U' = 1) = P(\text{positive test} \,|\, U' = 1, R'), \\ r_{\text{spec}} &= P(\text{negative test} \,|\, U' = 0) = P(\text{positive test} \,|\, U' = 0, R'). \end{aligned}$$

Then by the law of total probability, granted the hypnozoite reservoir has reached stationarity under a constant force of inoculation $\lambda$, the specificity of the test as a predictor of hypnozoite carriage takes the form

$$s_{\text{spec}}(\lambda) = (1 - r_{\text{sens}}) + (r_{\text{sens}} + r_{\text{spec}} - 1)h_{\text{spec}}(\lambda)$$

where $h_{\text{spec}}(\lambda)$ is given by Equation (E.12). Similarly, the sensitivity of the serological test as a predictor of hypnozoite carriage can be written

$$s_{\text{sens}}(\lambda) = (1 - r_{\text{spec}}) + (r_{\text{sens}} + r_{\text{spec}} - 1)h_{\text{sens}}(\lambda)$$

where we $h_{\text{sens}}(\lambda)$ is given by Equation (E.13). An analogous functional form holds in the presence of population heterogeneity in the force of inoculation.

#### E.4 On the classification of relapse vs primary infection

The classification of an observed vivax recurrence as a primary infection vs relapse is of epidemiological interest, with possible implications for treatment and control [32]. Relapses and primary infections, however, are typically indistinguishable in natural transmission settings.

Given the sequence of clinical recurrences recorded for a child, we can derive the probability that a given recurrence is a relapse i.e. can be attributed to a hypnozoite activation event only. In doing so, we leverage each inter-recurrence intervals recorded for a child (with recurrences in quick succession characteristic of a recently-established hypnozoite batch); in addition to seasonality. Probabilistic classifications for 7 year olds with at least two recorded clinical recurrences ($n = 56$ children) are shown in Figure E.1.

##### E.4.1 Derivation: probabilistic classification under the within-host framework

Probabilistic classification of observed recurrences necessitates the derivation of the multivariate PGF for the number of relapses $H_i$ and the number of primary infections $P_i$ experienced in each window $i$. We can derive this PGF using much the same reasoning as Appendix B.2 (differences

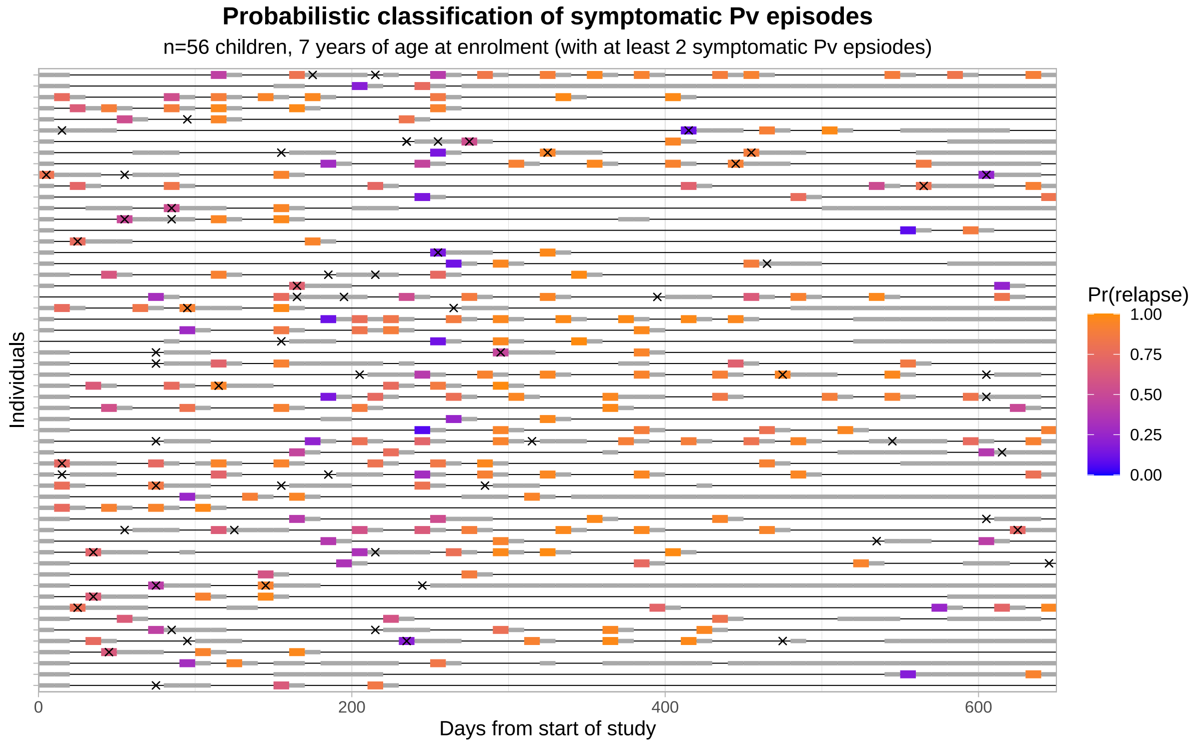

**Figure E.1:** Probabilistic classification of symptomatic vivax episodes for 7 year old children ($n = 56$) with at least two vivax episodes. Predictions are predicated on posterior median estimates for each parameter $\eta$, $\nu$, $\Lambda_1$, $\Lambda_2$, as well as the relative probability of symptomatic infection $p_{\text{clin}}(a)$ at age 7 compared to age 2. Symptomatic vivax episodes are shown with boxes, shaded according to the computed probability that they are attributable to hypnozoite activation event(s) only; dark grey boxes indicate recurrences which could not be classified due to numerical errors. Light grey bars indicate masking, either due to left/right-censoring, a documented absence from the camp or complete prophylactic protection following a previous bout of antimalarial treatment. Symptomatic falciparum episodes are marked with crosses.

are highlighted in red for clarity). Given a single inoculation at time $t_{j-1} < \tau < t_j$, we can show that

$$\begin{aligned}
&\mathbb{E}\left[ \prod_{i=1}^{n} y_i^{P_i} z_i^{H_i} \,\middle|\, \text{bite at time } t_{j-1} < \tau < t_j \right] \\
&\quad = {\color{red} y_j} \left( 1 + \nu p_{\text{rel}} \Big[ B(t_n - \tau) - z_j B(t_j - \tau) - \sum_{k=j+1}^{n} z_k \big[ B(t_k - \tau) - B(t_{k-1} - \tau) \big] \Big] \right)^{-1} \\
&\qquad + (1 - {\color{red} y_j}) \left( 1 + \nu(1 - p_{\text{rel}}) + \nu p_{\text{rel}} \Big[ B(t_n - \tau) - z_j B(t_j - \tau) - \sum_{k=j+1}^{n} z_k \big[ B(t_k - \tau) - B(t_{k-1} - \tau) \big] \Big] \Big\} \right)^{-1}.
\end{aligned} \tag{E.19}$$

As before, we impose the assumption of uniformly separated intervals $t_j = jT$ with a piecewise constant force of inoculation $\lambda_j$. Setting

$$W_i \big| P_{i+n_{\text{age}}} \sim \text{Binomial}\big(P_{i+n_{\text{age}}}, p_{\text{clin}}\big)$$
$$R_i \big| H_{i+n_{\text{age}}} \sim \text{Binomial}\big(H_{i+n_{\text{age}}}, p_{\text{clin}}\big)$$

we follow an identical argument to Appendix B.2 to obtain

$$\begin{aligned}
&G(\mathbf{w}, \mathbf{x}) := \mathbb{E}\left[ \prod_{i=1}^{n_{\text{obs}}} w_i^{W_i} x_i^{R_i} \right] \\
&= \prod_{j=1}^{n_{\text{age}}} e^{-\lambda_j T} \left( 1 - \frac{1 - e^{-\eta T}}{1 + \nu p_{\text{rel}} p_{\text{clin}} e^{-\eta T (n_{\text{age}} - j)} \left[ e^{-\eta T} - e^{-\eta T (n_{\text{obs}} + 1)} - (1 - e^{-\eta T}) \sum_{k=1}^{n_{\text{obs}}} x_k \cdot e^{-\eta T k} \right]} \right)^{-\frac{\lambda_j}{\eta}} \\
&\quad \prod_{j=1}^{n_{\text{obs}}} e^{-\lambda_j T} \left( 1 - \frac{(1 + \nu p_{\text{rel}} p_{\text{clin}} (1 - x_j))(1 - e^{-\eta T})}{1 + \nu p_{\text{rel}} p_{\text{clin}} - \nu p_{\text{rel}} p_{\text{clin}} [e^{-\eta (n_{\text{obs}} - j + 1) T} + (1 - e^{-\eta T}) \sum_{k=j}^{n_{\text{obs}}} x_k e^{-\eta (k - j) T}]} \right)^{-\frac{\lambda_j (1 - p_{\text{clin}} + p_{\text{clin}} {\color{red} w_j})}{\eta (1 + \nu p_{\text{rel}} p_{\text{clin}} (1 - x_j))}} \\
&\qquad \left( 1 - \frac{(1 + \nu(1 - p_{\text{rel}}(1 - p_{\text{clin}}) - p_{\text{rel}} p_{\text{clin}} x_j))(1 - e^{-\eta T})}{1 + \nu(1 - p_{\text{rel}}(1 - p_{\text{clin}})) - \nu p_{\text{rel}} p_{\text{clin}} [e^{-\eta (n_{\text{obs}} - j + 1) T} + (1 - e^{-\eta T}) \sum_{k=j}^{n_{\text{obs}}} x_k e^{-\eta (k - j) T}]} \right)^{-\frac{\lambda_j p_{\text{clin}} (1 - {\color{red} w_j})}{\eta (1 + \nu (1 - p_{\text{rel}} (1 - p_{\text{clin}}) - p_{\text{rel}} p_{\text{clin}} x_j))}}.
\end{aligned} \tag{E.20}$$

Application of the inclusion-exclusion principle allows us to recover the likelihood that each recurrence is a relapse or primary infection. Under our framework, we do not constrain the number of hypnozoite activation and/or immediate sporozoite development events in each window. As such, a recurrence in window $i$ can be attributed to both hypnozoite activation (relapse) and immediate sporozoite development (primary infection). In particular, we note that

$$\begin{aligned}
&\mathbb{E}\left[ \prod_{i=1}^{n_{\text{obs}}-1} w_i^{W_i} x_i^{R_i} \,\middle|\, W_{n_{\text{obs}}} = 0,\ R_{n_{\text{obs}}} > 0 \text{ i.e. relapse only at window } n_{\text{obs}} \right] \\
&\quad = G(w_1, \ldots, w_{n_{\text{obs}}-1}, 0, x_1, \ldots x_{n_{\text{obs}}-1}, 1) - G(w_1, \ldots, w_{n_{\text{obs}}-1}, 0, x_1, \ldots x_{n_{\text{obs}}-1}, 0) \\
&\mathbb{E}\left[ \prod_{i=1}^{n_{\text{obs}}-1} w_i^{W_i} x_i^{R_i} \,\middle|\, W_{n_{\text{obs}}} > 0,\ R_{n_{\text{obs}}} = 0 \text{ i.e. primary infection only at window } n_{\text{obs}} \right] \\
&\quad = G(w_1, \ldots, w_{n_{\text{obs}}-1}, 1, x_1, \ldots x_{n_{\text{obs}}-1}, 0) - G(w_1, \ldots, w_{n_{\text{obs}}-1}, 0, x_1, \ldots x_{n_{\text{obs}}-1}, 0)
\end{aligned}$$

$$\mathbb{E}\left[\prod_{i=1}^{n_{\text{obs}}-1} w_i^{W_i} x_i^{R_i} \,\middle|\, W_{n_{\text{obs}}} > 0,\, R_{n_{\text{obs}}} = 0 \text{ i.e. primary infection and relapse at window } n_{\text{obs}}\right]$$
$$= G(w_1, \dots, w_{n_{\text{obs}}-1}, 1, x_1, \dots x_{n_{\text{obs}}-1}, 1) - G(w_1, \dots, w_{n_{\text{obs}}-1}, 0, x_1, \dots x_{n_{\text{obs}}-1}, 1)$$
$$- G(w_1, \dots, w_{n_{\text{obs}}-1}, 1, x_1, \dots x_{n_{\text{obs}}-1}, 0) + G(w_1, \dots, w_{n_{\text{obs}}-1}, 0, x_1, \dots x_{n_{\text{obs}}-1}, 0).$$

#### E.5 Recurrences attributable to a single batch

Here, we examine the probability that successive recurrences are derived from different sporozoite batches. We make the simplifying assumption of a constant force of inoculation $\lambda$. We allow for the accrual of hypnozoites over the interval $(0, Y]$ before the study period $(Y, Y + nT]$, discretised into $n$ uniform windows of length $T$. We say that a batch "contributes" to the burden of bloodstream infection in window $j$ if either an inoculated sporozoite develops immediately, or a hypnozoite activates in window $j$.

Suppose a baseline recurrence is observed in window 1, and that the next recurrence is recorded in window $n$. Here, we seek to derive the probability that the same batch contributes to both windows 1 and $n$. We denote the arrival time of batch $i$ by $\tau_i$, and set:

- $W_i = 1$ if batch $i$ does not contribute to the burden of bloodstream infection in windows $2, \dots, (n-1)$; and $W_i = 0$ otherwise.
- $V_i = 1$ if batch $i$ contributes to the burden of bloodstream infection in windows 1 and $n$, but not $2, \dots, (n-1)$; and $V_i = 0$ otherwise.

The number of sporozoite batches $M$ inoculated in the interval $(0, Y + Tn]$

$$M \sim \text{Poisson}\big(\lambda(Y + nT)\big).$$

Given $M = m$, the batch arrival times

$$\tau_1, \dots, \tau_m \overset{\text{i.i.d.}}{\sim} \text{Uniform}[0, Y + nT].$$

Suppose batch $i$ is inoculated at time $x$, that is, $\tau_i = x$. The probability that sporozoite $i_k$ of batch $i$ activates in windows $2, \dots, (n-1)$ is given by

$$P(\text{sporozoite } i_k \text{ contributes in windows } 2, \dots, n-1 \,|\, \tau_i = x)$$

$$= \begin{cases} p_{\text{rel}} e^{-\eta(Y+T-x)}(1 - e^{-\eta T(n-2)}) & \text{if } x \leq Y + T \\ (1 - p_{\text{rel}}) + p_{\text{rel}}\big(1 - e^{-\eta(Y+(n-1)T-x)}\big) & \text{if } Y + T < x \leq Y + (n-1)T \\ 0 & \text{if } x > Y + (n-1)T. \end{cases}$$

Since each sporozoite is modelled to be governed by an independent stochastic process, accounting for a geometrically-distributed sporozoite batch size using the law of total probability yields

$$P(W_i = 1 \,|\, \tau_i = x) = \begin{cases} \frac{1}{1+\nu p_{\text{rel}} e^{-\eta(Y-x+T)}(1-e^{-\eta T(n-2)})} & \text{if } x \leq Y + T \\ \frac{1}{1+\nu-\nu p_{\text{rel}} e^{-\eta(Y+(n-1)T-x)}} & \text{if } Y + T < x \leq Y + (n-1)T \\ 1 & \text{if } x > Y + (n-1)T. \end{cases}$$

A further application of the law of total probability over the batch arrival time $\tau_i$ then yields

$$\begin{aligned} P(W_i = 1) &= \frac{1}{Y + nT} \int_0^{Y+nT} P(W_i = 1 \,|\, \tau_i = x) dx \\ &= \frac{1}{Y + nT} \Bigg[ T - \frac{1}{\eta} \log \left( 1 - \frac{1 - e^{-\eta(Y+T)}}{1 + \nu p_{\text{rel}} e^{-\eta(Y+T)} \big[1 - e^{-\eta T(n-2)}\big]} \right) \\ &\qquad - \frac{1}{\eta(1+\nu)} \log \left( 1 - \frac{(1+\nu)(1 - e^{-\eta T(n-2)})}{1 + \nu - \nu p_{\text{rel}} e^{-\eta T(n-2)}} \right) \Bigg] \end{aligned}$$

where we have used standard integral 2.313.1 of [23].

Likewise, given $\tau_i = x$, we adopt similar reasoning — and the inclusion-exclusion principle — to compute

$$P(V_i = 1) = \begin{cases} \frac{1}{1+\nu p_{\text{rel}} e^{-\eta(Y-x+T)}(1-e^{-\eta T(n-2)})} - \frac{1}{1+\nu p_{\text{rel}} e^{-\eta(Y-x+T)}(1-e^{-\eta T(n-1)})} \\ \quad - \frac{1}{1+\nu p_{\text{rel}} e^{-\eta(Y-x)}(1-e^{-\eta T(n-1)})} + \frac{1}{1+\nu p_{\text{rel}} e^{-\eta(Y-x)}(1-e^{-\eta T n})} & \text{if } x < Y \\ \frac{1}{1+\nu p_{\text{rel}}-\nu p_{\text{rel}} e^{-\eta(Y-x+T)}(1-e^{-\eta T(n-2)})} - \frac{1}{1+\nu p_{\text{rel}}-\nu p_{\text{rel}} e^{-\eta(Y-x+T)}(1-e^{-\eta T(n-1)})} \\ \quad - \frac{1}{1+\nu-\nu p_{\text{rel}} e^{-\eta(Y+(n-1)T-x)}} + \frac{1}{1+\nu-\nu p_{\text{rel}} e^{-\eta(Y+nT-x)}} & \text{if } Y < x \leq Y + T \\ 0 & \text{if } x > Y + T. \end{cases}$$

Using the law of total probability to account for stochasticity in the batch arrival time $\tau_i$, it follows that

$$\begin{aligned} P(V_i = 1) =& \frac{1}{Y + nT} \int_0^{Y+nT} P(V_i = 1 \,|\, \tau_i = x) dx \\ =& -\frac{1}{\eta(Y + nT)} \Bigg[ \log \left( 1 - \frac{1 - e^{-\eta(Y+T)}}{1 + \nu p_{\text{rel}} e^{-\eta(Y+T)} \big[1 - e^{-\eta T(n-2)}\big]} \right) \end{aligned}$$

$$
\begin{aligned}
&- 2\log\left(1 - \frac{1-e^{-\eta(Y+T)}}{1+\nu p_{\text{rel}} e^{-\eta(Y+T)}\left[1-e^{-\eta T(n-1)}\right]}\right) \\
&+ \log\left(1 - \frac{1-e^{-\eta(Y+T)}}{1+\nu p_{\text{rel}} e^{-\eta(Y+T)}\left[1-e^{-\eta T n)}\right]}\right)\Bigg] \\
&+ \frac{1}{1+\nu}\log\Big(1+\nu-\nu p_{\text{rel}} e^{-\eta n T}\Big) \\
&- \frac{2}{1+\nu}\log\Big(1+\nu-\nu p_{\text{rel}} e^{-\eta(n-1)T}\Big) \\
&+ \frac{1}{1+\nu}\log\Big(1+\nu-\nu p_{\text{rel}} e^{-\eta(n-2)T}\Big)\Bigg]
\end{aligned}
$$

where we have again used standard integral 2.313.1 of [23].

Given the arrival of precisely $M = m$ batches in the interval $[0, Y + nT]$, we denote by $C(m)$ the probability that the same batch contributes to both windows 1 and $n$, while no recurrences occur in windows $2, \ldots, (n-1)$. By homogeneity, since the batch arrival times $\tau_1, \ldots, \tau_m$ are i.i.d.

$$C(m) = m \cdot P(V_i = 1) \cdot P(W_i = 1)^{m-1}.$$

Denote by $B(Y, n)$ the indicator for the event that, following a baseline recurrence in window 1, the next recurrence occurs at window $n$ *and* the same batch contributes to both recurrences in windows 1 and $n$. Applying the law of total probability over the number of sporozoite batches $M$ inoculated in the interval $[0, Y + nT]$, we obtain

$$
\begin{aligned}
P(B(Y,n)=1) &= \sum_{m=1}^{\infty} C(m) \cdot \frac{1}{m!} e^{-\lambda(Y+nT)} \lambda^m (Y+nT)^m \\
&= \lambda(Y+nT) P(V_i=1) \cdot e^{-\lambda(Y+nT)[1-P(W_i=1)]}
\end{aligned}
$$

recognising the Taylor expansion of the exponential function.

##### E.5.1 Metrics under a stationary hypnozoite burden

In the limit $Y \to \infty$ — that is, allowing the hypnozoite reservoir to reach stationarity prior to the manifestation of the baseline recurrence — we obtain

$$
\begin{aligned}
S(n) :=& \lim_{Y\to\infty} P(B(Y,n)=1) \\
=& \frac{1}{\eta}\Bigg[\log\left(\frac{\left[1+\nu p_{\text{rel}}(1-e^{-\eta(n-1)T})\right]^2}{\left[1+\nu p_{\text{rel}}(1-e^{-\eta(n-2)T})\right]\left[1+\nu p_{\text{rel}}(1-e^{-\eta n T})\right]}\right)
\end{aligned}
$$

$$\frac{1}{1+\nu}\log\left(\frac{\left[1+\nu-\nu p_{\text{rel}}e^{-\eta(n-1)T}\right)\right]^2}{\left[1+\nu-\nu p_{\text{rel}}e^{-\eta(n-2)T}\right)\right]\left[1+\nu-\nu p_{\text{rel}}e^{-\eta nT}\right)\right]}\right)\Bigg]$$

$$\times e^{-\lambda(n-2)T}\Big(1+\nu p_{\text{rel}}\big(1-e^{-\eta T(n-2)}\big)\Big)^{-\frac{\lambda}{\eta}}\left(1-\frac{(1+\nu)(1-e^{-\eta(n-2)T})}{1+\nu-\nu p_{\text{rel}}e^{-\eta(n-2)T}}\right)^{-\frac{\lambda}{\eta(1+\nu)}}. \tag{E.21}$$

From the multivariate PGF (B.10), granted the hypnozoite reservoir has reached stationarity, the probability of a baseline recurrence in window 1 is

$$R_1(1)=1-e^{-\lambda T}\Big(1+\nu p_{\text{rel}}\big(1-e^{-\eta T}\big)\Big)^{-\frac{\lambda}{\eta}}\left(1-\frac{(1+\nu)(1-e^{-\eta T})}{1+\nu-\nu p_{\text{rel}}e^{-\eta T}}\right)^{-\frac{\lambda}{\eta(1+\nu)}}, \tag{E.22}$$

while the joint probability of a baseline recurrence in window 1 and a subsequent recurrence in window $n$ is given by

$$\begin{aligned}R_2(1,n):=&e^{-\lambda(n-2)T}\Big(1+\nu p_{\text{rel}}\big(1-e^{-\eta T(n-2)}\big)\Big)^{-\frac{\lambda}{\eta}}\left(1-\frac{(1+\nu)(1-e^{-\eta(n-2)T})}{1+\nu-\nu p_{\text{rel}}e^{-\eta(n-2)T}}\right)^{-\frac{\lambda}{\eta(1+\nu)}}\\&-2e^{-\lambda(n-1)T}\Big(1+\nu p_{\text{rel}}\big(1-e^{-\eta T(n-1)}\big)\Big)^{-\frac{\lambda}{\eta}}\left(1-\frac{(1+\nu)(1-e^{-\eta(n-1)T})}{1+\nu-\nu p_{\text{rel}}e^{-\eta(n-1)T}}\right)^{-\frac{\lambda}{\eta(1+\nu)}}\\&+e^{-\lambda nT}\Big(1+\nu p_{\text{rel}}\big(1-e^{-\eta Tn}\big)\Big)^{-\frac{\lambda}{\eta}}\left(1-\frac{(1+\nu)(1-e^{-\eta nT})}{1+\nu-\nu p_{\text{rel}}e^{-\eta nT}}\right)^{-\frac{\lambda}{\eta(1+\nu)}}\end{aligned} \tag{E.23}$$

where we have used the inclusion-exclusion principle.

The conditional probability that, given a baseline recurrence in window 1, the next recurrence will occur in window $n$, is given by the quotient $R_2(1,n)/R_1(1)$. Given a set of consecutive recurrences in windows 1 and $n$, the conditional probability that the same sporozoite batch contributes to both windows is given by the quotient $S(n)/R_2(1,n)$.

#### E.6 Detectable relapses

A critical feature of *P. vivax* infection is the "remarkable periodicity" of early inter-relapse intervals [1, 33]. White et al. [18] have shown through simulation that periodicity in detected relapses can emerge through a masking process, rendering activated hypnozoites undetectable if they arise within a fixed window of $T = 14$ days of the most recent detected relapse. Here, we present a more thorough probabilistic characterisation of this process, characterising both the number and timing of detectable relapses in detail.

Denote by $H_i$ the size of the hypnozoite reservoir just before the onset of the $i$th detectable relapse. If less than $i$ relapses occur, we set $H_k = 0$ for all $k \geq i$. The initial size of the hypnozoite reservoir $H_1$ is assumed to have PGF

$$G_1(z_1) := \mathbb{E}\left[z_1^{H_1}\right].$$

We note that $H_{n+1}$ is dependent only on $H_n$, that is, the depletion of the hypnozoite reservoir over successive relapses is Markovian. Each hypnozoite persists through the masking period (of fixed length $T$) with probability $e^{-\eta T}$. Under the assumption of hypnozoite independence, the hypnozoite burden upon the onset of the $(n+1)$th relapse is binomially-distributed conditional on $H_n = N \geq 1$, that is,

$$H_{n+1} \stackrel{d}{=} \begin{cases} \text{Binomial}(N-1, e^{-\eta T}) & \text{if } H_n = N \geq 1 \\ 0 & \text{if } H_n = 0. \end{cases}$$

Here, we propose an iterative procedure to recover the multivariate PGF of $H_1, \dots, H_n$, that is,

$$G_n(z_1, \dots, z_n) = \mathbb{E}\left[\prod_{i=1}^{n} z_i^{H_i}\right] = \sum_{j_1=1}^{\infty} \cdots \sum_{j_n=1}^{\infty} P(H_1 = j_1, \dots, H_n = j_n) \prod_{i=1}^{n} z_i^{j_i}.$$

The PGF $G_{n+1}$ follows readily from $G_n$ through a direct application of the law of total expectation:

$$\begin{aligned} G_{n+1}(z_1, \dots, z_n, z_{n+1}) &= \mathbb{E}\left[\prod_{i=1}^{n+1} z_i^{H_i}\right] \\ &= \sum_{j_1=0}^{\infty} \cdots \sum_{j_n=0}^{\infty} P(H_1 = j_1, \dots, H_n = j_n) \cdot \left(\prod_{i=1}^{n} z_i^{j_i}\right) \cdot \underbrace{\left(1 - e^{-\eta T} + z_{n+1} e^{-\eta T}\right)^{\max\{0, j_n - 1\}}}_{\text{binomial PGF for } H_{n+1} \text{ given } H_n = j_n} \\ &= \frac{G_n\big(z_1, \dots, z_{n-1}, z_n \cdot \left[1 - e^{-\eta T} + z_{n+1} e^{-\eta T}\right]\big) - G_n(z_1, \dots, z_{n-1}, 0)}{1 - e^{-\eta T} + z_{n+1} e^{-\eta T}} + G_n(z_1, \dots, z_{n-1}, 0). \end{aligned}$$

We can use the joint PGF $G_n$ to recover quantities of epidemiological interest. The likelihood that a single mosquito inoculation event gives rise to precisely $\ell$ detectable relapses is given by

$$\begin{aligned} &P(\ell \text{ detectable relapses}) = P(H_{\ell+1} = 0,\, H_\ell > 0) \\ &\quad = G_{\ell+1}(z_1 = 1, \dots, z_{\ell-1} = 1, z_\ell = 1, z_{\ell_1} = 0) - G_{\ell+1}(z_1 = 1, \dots, z_{\ell-1} = 1, z_\ell = 0, z_{\ell_1} = 0). \end{aligned}$$

We can also recover precise distributions of inter-relapse times. For notational convenience, we consider the marginalised PGF

$$G_\ell^{(i)}(z_i, z_\ell, z_{\ell+1}) := \mathbb{E}\left[z_i^{H_i} z_\ell^{H_\ell} z_{\ell+1}^{H_{\ell+1}}\right],$$

obtained by setting $z_j = 1$ for all $j \notin \{i, \ell, \ell+1\}$ in $G_{\ell+1}$. Given precisely $\ell$ relapses occur, the size of the hypnozoite $H_i$ reservoir just before the onset of the $i$th relapse, $i \leq \ell$ has PGF

$$\mathbb{E}\left[z^{H_i} \middle| H_{\ell+1} = 0,\, H_\ell > 0\right] = \frac{G_\ell^{(i)}(z,1,0) - G_\ell^{(i)}(z,0,0)}{G_\ell^{(i)}(1,1,0) - G_\ell^{(i)}(1,0,0)}.$$

Let $T_\ell^{(i)}$ denote the $i$th inter-relapse time, given precisely $\ell$ relapses are detected. By the law of total expectation,

$$\begin{aligned} P\big(T_\ell^{(i)} \leq x\big) &= \mathbb{1}_{\{x \geq T\}} \cdot \sum_{n=0}^{\infty} \underbrace{\left[1 - e^{-\eta n(x-T)}\right]}_{\substack{\text{time to first activation} \\ \text{for } n \text{ hypnozoites}}} \cdot P\big(H_i = n | H_{\ell+1} = 0, H_\ell > 0\big) \\ &= 1 - \mathbb{1}_{\{x \geq T\}} \cdot \frac{G_\ell^{(i)}(e^{-\eta(x-T)},1,0) - G_\ell^{(i)}(e^{-\eta(x-T)},0,0)}{G_\ell^{(i)}(1,1,0) - G_\ell^{(i)}(1,0,0)}. \end{aligned}$$

### Appendix F

### Theses of vivax relapse biology

Here, we interrogate the calibrated model to address the 8 theses of vivax relapse biology posited by White [1] (reproduced below). Metrics of biological interest are predicated on posterior median estimates for the hypnozoite activation rate $\eta$ and mean sporozoite batch size $\nu$ (Table C.1), in addition to the hypnozoite fating probability $p_{\text{rel}} = 0.4$ informed by *in vivo* and *in vitro* experiments on the Chesson strain of *P. vivax* [28].

##### Thesis 1

*Relapses show remarkable periodicity.*

Post-treatment prophylaxis conferred by the commonly used antimalarial drugs chloroquine and mefloquine, as given to children in the SPf66 vaccine trial, can result in apparent periodicity under the exponential clock model [18]. This is because the drugs are eliminated slowly and suppress asexual stage parasite multiplication (commonly termed post-treatment prophylaxis). However, here, we consider inter-relapse intervals for a single infective bite, whilst ignoring the effects of post-treatment prophylaxis (i.e. intervals between successive hypnozoite activation events).

Given a hypnozoite batch of size $H = h \geq m$, the $m^{\text{th}}$ inter-relapse interval $T_m$ is exponentially-distributed with expectation and standard deviation

$$\mathbb{E}[T_m|H = h] = \text{SD}(T_m|H = h) = \frac{1}{\eta(h - m + 1)}$$

scaling inversely with the size of the remaining hypnozoite reservoir. As such, the progressive lengthening of successive inter-relapse intervals as the hypnozoite reservoir is depleted is accompanied by the increasing variability of inter-relapse intervals. Accounting for a geometrically-distributed hypnozoite batch $H$ of mean size $\nu p_{\text{rel}}$, the coefficient of variation (CV) for the $m^{\text{th}}$

inter-relapse interval $T_m$

$$\mathrm{CV}(T_m|H \geq m) = \frac{\mathrm{SD}(T_m|H \geq m)}{\mathbb{E}[T_m|H \geq m]}$$

can be shown to be a function of the average hypnozoite batch size $\nu p_{\mathrm{rel}}$ only, independent of both $m$ and $\eta$; that is, the standard deviation for the $m^{\mathrm{th}}$ inter-relapse interval scales linearly with the mean for the $m^{\mathrm{th}}$ inter-relapse interval. We estimate the CV for inter-relapse intervals arising from a single infective bite to be 1.41 (95% CrI 1.35 to 1.48). As a comparator, the CV for the exponential distribution is 1; a CV exceeding 1 can be interpreted to signify a relatively high degree of variability in the $m^{\mathrm{th}}$ inter-relapse interval following an infective bite across a hypothetical population of individuals. In light of this variability, echoing [18], we suggest that the temporal sequence of hypnozoite activation events is not intrinsically periodic, so under the exponential clock hypothesis relapse periodicity must be attributed to an external process e.g. post-treatment propyhlaxis.

**Thesis 2**

*Early relapses reach patency around three weeks after starting treatment which suggests emergence from the liver at least one week earlier.*

This thesis refers to rapidly eliminated treatments. In any given 2 week interval, the probability of activation for each hypnozoite is $p_{\mathrm{act}} = 0.08$ (95% CrI 0.07 to 0.09). One hypnozoite's progeny can cause a relapse so this can be interpreted as the expected proportion of relapses attributable to a single inoculum that will reach patency within 3 weeks. The probability of a spontaneous hypnozoite activation event within 2 weeks of treatment for a previous episode is a direct function of the hypnozoite burden $H$:

$$P(\text{relapse within 3 weeks of treatment}) = 1 - (1 - p_{\mathrm{act}})^H.$$

Following a single infective bite, the probability that at least one hypnozoite activates within the initial 2 week period is estimated to be 0.18 (95% CrI 0.15 to 0.22).

**Thesis 3**

*Not all P. vivax primary infections are followed by a relapse. In Thailand approximately 50% of infections are followed by a subsequent relapse within 28 days if a rapidly eliminated antimalarial drug (artesunate) is given for treatment of the primary infection and primaquine is not given. Elsewhere the probability of relapse generally varies between 20% and 80%. Animal experiments, the malaria therapy experience, and volunteer studies all suggest this proportion is a function of sporozoite inoculum.*

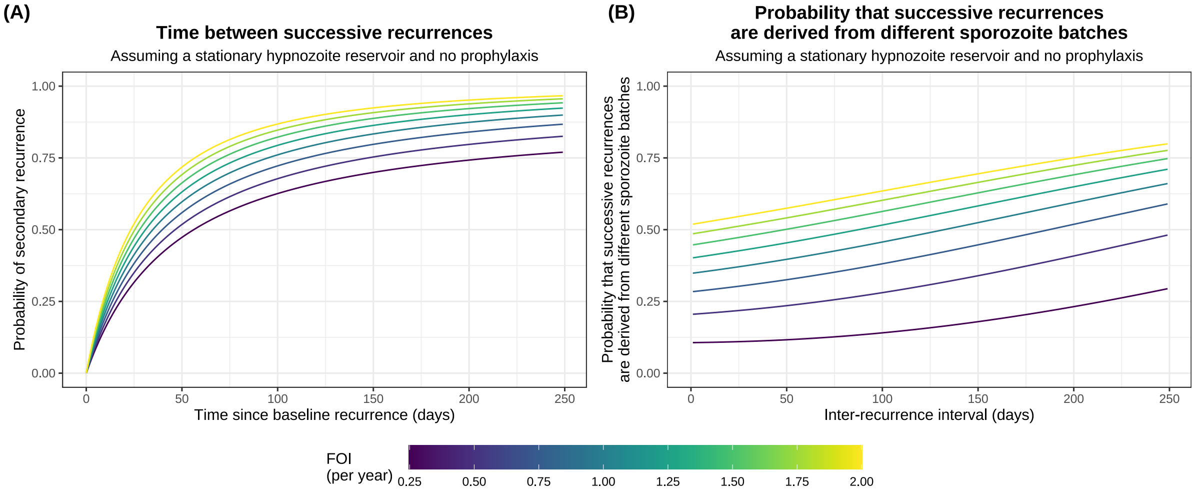

**Figure F.1:** Based on the model and derived parameters, successive recurrences in an endemic setting are shown as a function of the force of inoculation (FOI). Panel A shows the cumulative distribution function for the time between successive recurrences (i.e. the conditional probability that a secondary recurrence has occurred by day $n$, given a baseline recurrence on day 1). Panel B shows the conditional probability that a pair of successive recurrences with inter-recurrence interval $n$ days are derived from different sporozoite batches. Relevant formulae, derived under the assumption that the hypnozoite reservoir has reached stationarity under a constant FOI, are provided in Appendix E.5.

We estimate that 80% (95% CrI 77% to 84%) of infective bites give rise to a primary infection, while 6.8% (95% CrI 5.8% to 8.0%) of bites cause at least one relapse without a preceding primary infection (although in this case, the initial relapse would manifest as a primary infection — albeit with a longer incubation period). Conditional on the inoculation of at least one successful sporozoite, each infective bite gives rise to two or more recurrences with probability 79% (95% CrI 75% to 83%). We can interpret this as the estimated proportion of ostensibly primary infections that would eventually be followed by a second relapse, given a single inoculation.

In the interpretation of recurrent infections in an endemic setting, it is necessary to account for both the pre-existing hypnozoite reservoir and the possibility of further mosquito inoculation. The cumulative distribution function for the time between successive recurrences (in the absence of post-treatment prophylaxis) is shown in Figure F.1A as a function of the the force of inoculation. Under a force of inoculation of 0.5 bites per year, we predict 38% of baseline recurrences to be followed by a second recurrence within 28 days of follow-up, with this figure rising to 55% under a more intense force of inoculation of 2 bites per year. We note that these estimates do not account for immunity, which may lead to asymptomatic or low-density recurrences remaining undetected.

**Thesis 4**

*Multiple relapses are common, particularly in young children, even though sporozoite inocula are thought to be relatively small (median 6-10 sporozoites). It is not uncommon in tropical areas for children to have four to six relapses at 4-6 week intervals and sometimes more following an incident infection. Even larger numbers of relapses were observed in soldiers following intense exposure and in Rhesus monkeys receiving very large sporozoite inocula. Importantly the fraction of people experiencing a relapse after each illness episode in a particular location appears constant.*

Under the assumption of geometrically-distributed sporozite batch sizes, we estimate that each bite establishes a mean of $\nu p_{\text{rel}} = 2.7$ (95% CrI 2.2 to 3.5) hypnozoites that are destined to activate eventually (this estimate appears to be insensitive to the hypnozoite fating probability $p_{\text{rel}}$, see Appendix D.2). Accounting for the masking of hypnozoite activation events within a 10 day interval of a previous relapse (i.e. the possibility that a relapse would not be detected because it coincides with an earlier infection), we estimate that approximately 26% of children will experience 4 or more "detectable" (but not necessarily symptomatic) relapses following a single inoculation, while 12% of children will experience 6 or more detectable relapses (Figure F.2A). If masking is longer then the proportion is correspondingly lower. Inter-relapse intervals are expected to lengthen progressively as the hypnozoite reservoir is depleted (Figure F.2B). The manifestation of four to six relapses, each in successive 4 to 6 week intervals, might be plausible for a bite giving rise to 8 or more detectable relapses (Figure F.2B). This is predicted to occur for 5.2% of infective bites. Tightly clustered bouts of successive relapses following a single infective bite are therefore not inherently incompatible with the exponential clock model, but they are predicted to be relatively unlikely under our estimates for the mean hypnozoite batch size.

By construction, a geometrically-distributed sporozoite inoculum is compatible with a constant fractional reduction in the proportion of individuals experiencing successive relapses following a single inoculation (e.g. in the case of malaria therapy or "volunteer" studies), with the fractional reduction dependent on the mean inoculum size.

**Thesis 5**

*In long-latency phenotypes there is commonly a period of 8-9 months either before the first symptomatic infection, or between the first symptomatic infection and the first relapse. This long-latency interval appears to be normally distributed (mode 28 weeks for the Madagascar strain. Sometimes there are several short interval relapses followed by a long interval. Conversely long*

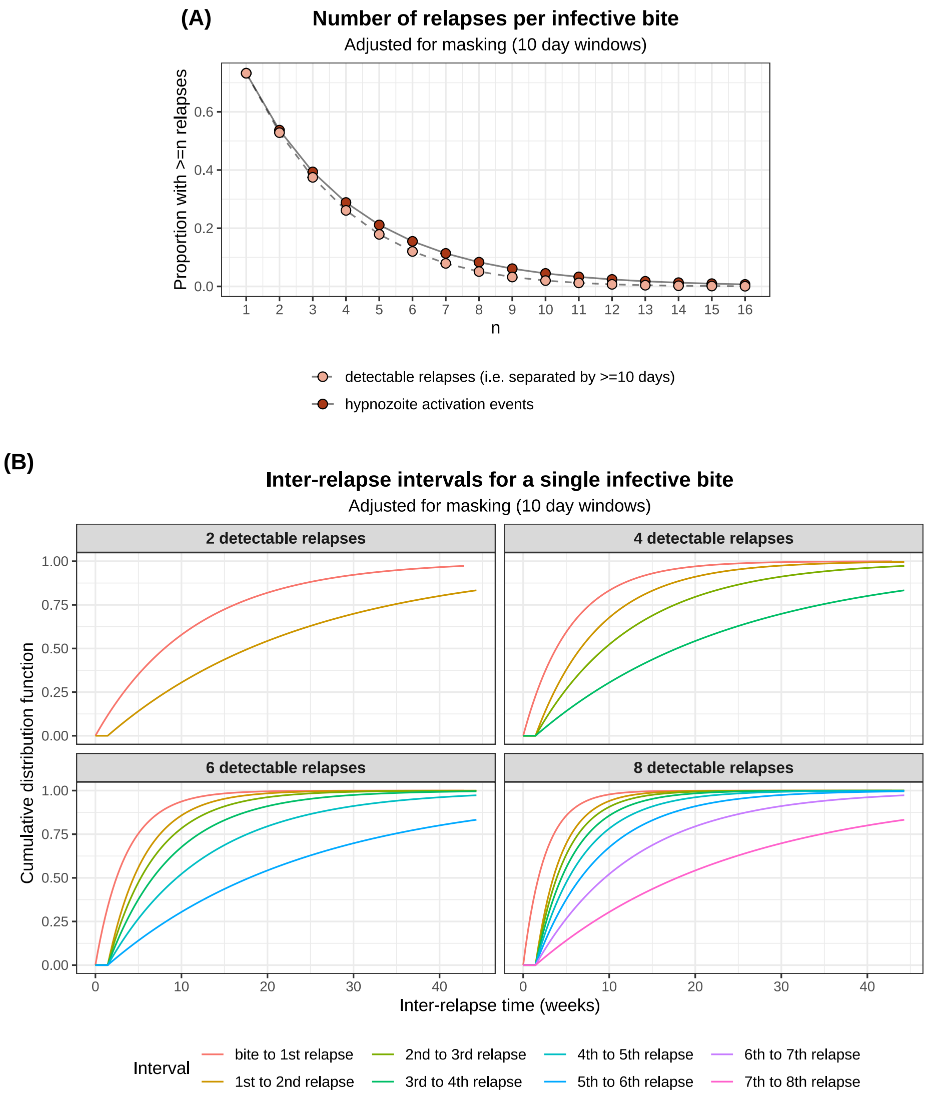

**Figure F.2:** Under the model, the distribution of relapses attributable to a single, geometrically-distributed sporozoite inoculum. The first hypnozoite activation event (if it occurs) is assumed to cause to a "detectable" relapse, but subsequent hypnozoite activation events are detectable only if they occur more than 10 days after the most recent detectable relapse. Panel A shows the tail distribution for the number of hypnozoite activation events vs detectable relapses. Panel B shows the cumulative distributions functions for inter-relapse intervals, conditional on the total number of detected relapses per bite. Distributions have been calculated using the formulae derived in Appendix E.6.

*latencies may also occur after multiple relapses in the tropical frequent relapse phenotype.*

To explain long-latency phenotypes, a biological clock mechanism has been hypothesised to give rise to a pre-programmed, genetically-determined dormancy period, before which hypnozoite activation is prohibited [1, 34, 35]. This can be modelled by coupling the exponential clock model to an enforced dormancy period [18, 36].

Under the exponential clock model, the expected time to relapse scales inversely with the size of the hypnozoite reservoir: halving the hypnozoite reservoir doubles the expected time to relapse. Accordingly, we predict inter-relapse intervals to increase progressively as the hypnozoite reservoir is depleted. As such, the exponential clock mechanism is compatible with the observation that for tropical phenotypes, multiple relapses in rapid succession may be followed by a long inter-relapse interval (Figure F.2B). When coupled to an enforced dormancy period, the exponential clock model is also compatible with the observation that the initial relapse for long-latency phenotypes may be followed by several short interval relapses, and then a longer relapse interval. Due to the inherent variability of inter-relapse intervals, long and short intervals may also be interspersed.

**Thesis 6**

*If there are further relapses after the long latent period then they occur frequently with short intervals which are very similar to those observed in the tropical "strains".*

The long-latent period has been modelled with an Erlang-distributed dormancy period, in association with the biological clock mechanism, that is either collective for each batch of hypnozoites [18] or independent for each hypnozoite [36]. Upon emergence from dormancy, an identical activation process has been assumed for long-latency hypnozoites [18]. If the exponential clock mechanism can recapitulate inter-relapse intervals for tropical strains, then the within-host models of [18, 36] may explain relapse dynamics for long-latency phenotypes.

**Thesis 7**

*The relapses in clinical studies conducted in endemic areas are commonly with a genotype which is different to that identified in the primary infection (48% in Columbian isolates, 55% in Indian isolates, 61% in Thai and Burmese isolates, and 71% in East Timor isolates).*

The probability that successive recurrences are derived from different sporozoite batches is predicted to be an increasing function of both the force of inoculation, and the inter-recurrence interval (Figure F.1B). Given a secondary recurrence occurs precisely 28 days after a baseline recurrence, we predict that it is derived from a different sporozoite batch with probability 0.22

under a force of inoculation of 0.5 bites per year, but a substantially higher probability of 0.55 under a more intense force of inoculation of 2 bites per year. To characteristic genetic relationships between successive recurrences, we would need to couple our within-host framework of recurrence to a comparative model of genetic similarity of sporozoites within vs between batches — which, in turn, would exhibit dependence on transmission intensity.

**Thesis 8**

*A remarkably high proportion of acute infections with Plasmodium falciparum are followed by an episode of P. vivax infection. The proportion is currently 30% in Thailand and 50% in Myanmar. The intervals between the acute P. falciparum malaria illness and the subsequent P. vivax malaria are similar to those between acute P. vivax malaria and the subsequent P. vivax relapse. The epidemiological characteristics suggest that these are all relapses.*

Survival curves for the time to first vivax recurrence following falciparum monoinfection, with treatment arms matched for the history of vivax malaria and seasonality (Figure A.2B) suggest that mefloquine eliminates bloodstream infections which emerge in the first month after treatment. After adjusting for post-treatment prophylaxis, the exponential clock model is generally able to explain observed rates of vivax malaria following falciparum monoinfections treated with artesunate-mefloquine combination therapy. In contrast, the model is unable to capture the substantially higher observed rates of vivax malaria following falciparum monoinfections treated with artesunate monotherapy. This could be explained by an external triggering mechanism, but on a time scale that is overwhelmed by the extended duration of prophylactic protection provided by mefloquine treatment i.e. we hypothesise that hypnozoites that undergo activation following the febrile falciparum stimulus malaria are often unable to establish bloodstream infections because they are eliminated by mefloquine post-treatment prophylaxis.
